# Auricular Vagus Neuromodulation – A Systematic Review on Quality of Evidence and Clinical Effects

**DOI:** 10.1101/2020.11.26.20239509

**Authors:** Nishant Verma, Jonah D Mudge, Maïsha Kasole, Rex C Chen, Stephan L Blanz, James K Trevathan, Eric Lovett, Justin C Williams, Kip A Ludwig

## Abstract

This review is intended to identify key gaps in the mechanistic knowledge and execution of aVNS studies, to be addressed in future works, and aid the successful translation of neuromodulation therapies.

**Background:** The auricular branch of the vagus nerve runs superficial to the surface of the skin, which makes it a favorable target for non-invasive stimulation techniques to modulate vagal activity. For this reason, there have been many early-stage clinical trials on a diverse range of conditions. These trials often report conflicting results for the same indication.

**Methods:** Using the Cochrane Risk of Bias tool we conducted a systematic review of auricular vagus nerve stimulation (aVNS) randomized controlled trials (RCTs) to identify the factors that led to these conflicting results. As is common for early-stage studies, the majority of aVNS studies were assessed as having ‘some’ or ‘high’ risk of bias, which makes it difficult to interpret their results in a broader context.

**Results:** There is evidence of a modest decrease in heart rate during higher stimulation dosages, sometimes at above the level of sensory discomfort. Findings on heart rate variability conflict between studies and are hindered by trial design, including inappropriate washout periods, and multiple methods used to quantify heart rate variability. There is early-stage evidence to suggest aVNS may reduce circulating levels and endotoxin-induced levels of inflammatory markers. Studies on epilepsy reached primary endpoints similar to previous RCTs testing implantable vagus nerve stimulation (VNS) therapy, albeit with concerns over quality of blinding. Preliminary evidence shows that aVNS ameliorated pathological pain but not evoked pain.

**Discussion:** Based on results of the Cochrane analysis we list common improvements for the reporting of results, which can be implemented immediately to improve the quality of evidence. In the long term, existing data from aVNS studies and salient lessons from drug development highlight the need for direct measures of local neural target engagement. Direct measures of neural activity around the electrode will provide data for the optimization of electrode design, placement, and stimulation waveform parameters to improve on-target engagement and minimize off-target activation. Furthermore, direct measures of target engagement, along with consistent evaluation of blinding success, must be used to improve the design of controls in the long term – a major source of concern identified in the Cochrane analysis.

**Conclusion:** The need for direct measures of neural target engagement and consistent evaluation of blinding success is applicable to the development of other paresthesia-inducing neuromodulation therapies and their control designs.

## 1. INTRODUCTION

Electrical stimulation of the nervous system, commonly known as neuromodulation, manipulates nervous system activity for therapeutic benefits. The wandering path of the vagus nerve, the tenth cranial nerve, and its communication with several visceral organs and brain structures makes it an attractive target to address many diseases. Vagus nerve stimulation (VNS) to treat epilepsy has been approved by the United States Food and Drug Administration (FDA) since 1997 (Wellmark, 2018). An implantable pulse generator (IPG) is implanted below the clavicle and delivers controlled doses of electrical stimulation through electrodes wrapped around the cervical vagus. Due to the safety versus efficacy profile of the therapy, implantable VNS is currently a last line therapy after patients have been shown refractory to at least two appropriately dosed anti-epileptic drugs (American Association of Neurological Surgeons, 2021). Implantable VNS for epilepsy is purported to work through vagal afferents terminating in the nucleus of the solitary tract (NTS). NTS in turn has direct or indirect projections to the nuclei providing noradrenergic, endorphinergic, and serotonergic fibers to different parts of the brain (Kaniusas et al., 2019).

In a similar fashion, the auricular branch of the vagus also projects to the NTS, carrying somatosensory signals from the ear (Kaniusas et al., 2019). The superficial path of the nerve (Bermejo et al., 2017) in the ear means a low amplitude electrical stimulation applied at the surface of the skin can, in theory, generate electric field gradients at the depth of the nerve sufficient to alter its activity. Auricular vagus nerve stimulation (aVNS) delivered percutaneously or transcutaneously offers a method to modulate neural activity on the vagus nerve with the potential for a more favorable safety profile. Fig. 1 shows innervation of the auricle by four major nerve branches, overlapping regions of innervation in the auricle, and several electrode designs to deliver electrical stimulation at the ear.

**Figure 1:**
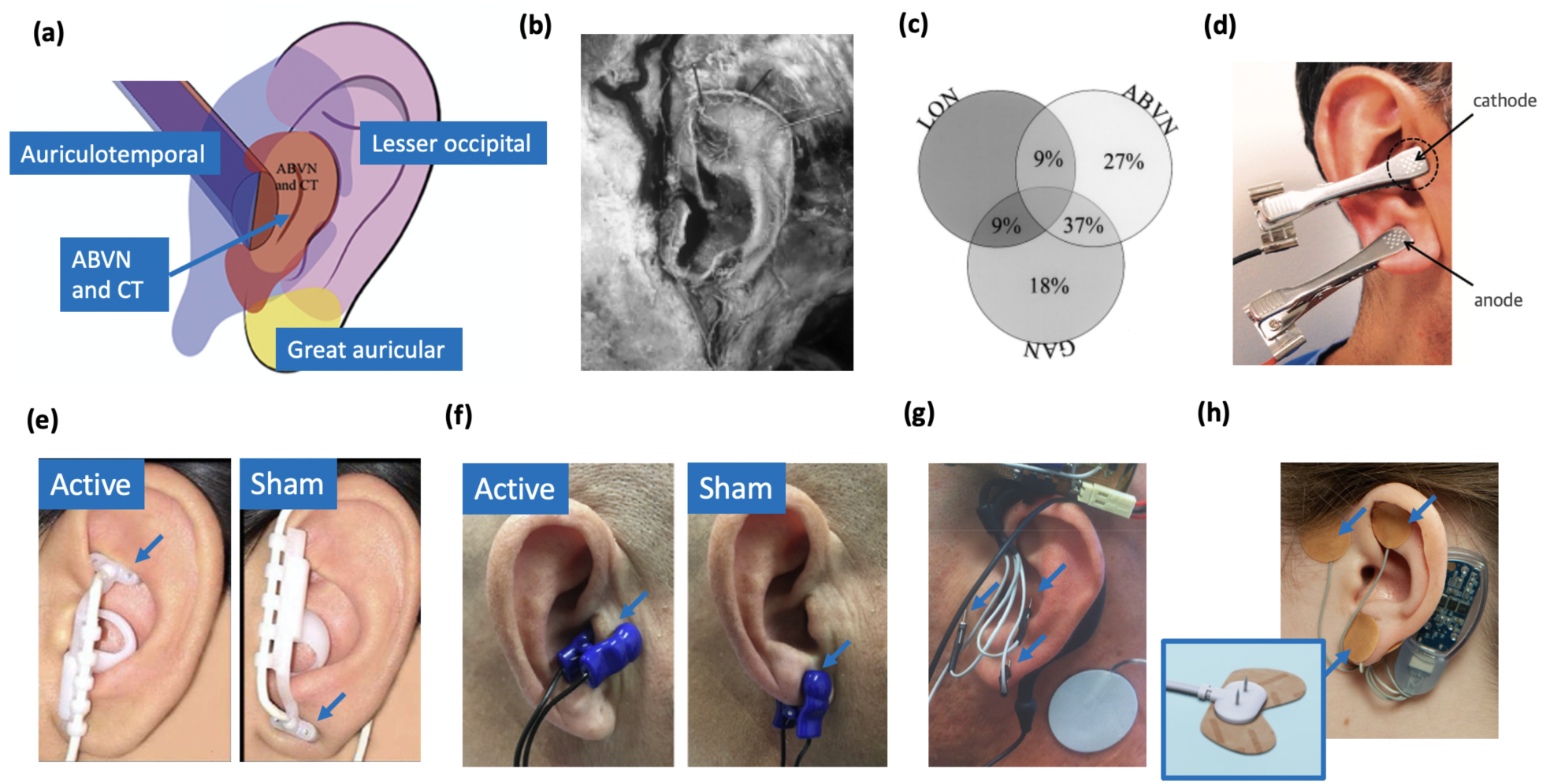
**(a)** Innervation of the auricle by five nerves (Watanabe et al., 2016): auricular branch of the vagus nerve (ABVN), chorda tympani (CT) from the facial nerve, auriculotemporal nerve originating from the mandibular branch of the trigeminal nerve, great auricular nerve, and lesser occipital nerve. **(b)** The only primary data study (Peuker and Filler, 2002) mapping the innervation of the human auricle in 7 cadavers by dissection. Note, microdissection cannot trace the finest of nerve branches. **(c)** Overlapping regions of innervation reported between the auricular branch of the vagus nerve (ABVN), great auricular nerve (GAN), and lesser occipital nerve (LON) (Peuker and Filler, 2002). Here the ABVN and GAN overlap for 37% of the area on the medial dorsal middle third of the ear setting the precedent for large overlaps in regions innervated by different nerves. Commonly used electrodes: **(d)** Clips transcutaneously targeting the tragus and earlobe simultaneously in the intervention group (Stavrakis et al., 2015). **(e)** NEMOS electrodes by Cerbomed (Erlangen, Germany) transcutaneously targeting cymba concha in the intervention group and ear lobe in the sham group. **(f)** Parasym (London, UK) transcutaneously targeting tragus in the intervention group and ear lobe in the sham group. **(g)** Percutaneously targeting intrinsic auricular muscles zones in the intervention group (Cakmak et al., 2017). **(h)** Innovative Health Solutions (Versailles, IN, USA) percutaneously targeting several cranial nerves in the auricular and periauricular region in the intervention group.

Given aVNS can be implemented with minimally invasive approaches and has the potential to modulate vagal activity, there have been many early-stage clinical trials investigating a diverse range of potential therapeutic indications, including heart failure, epilepsy, depression, pre-diabetes, Parkinson’s, and rheumatoid arthritis. Several companies are already developing aVNS devices, such as Parasym (London, UK), Cerbomed (Erlangen, Germany), Spark Biomedical (Dallas, Texas, USA), SzeleSTIM (Vienna, Austria), Ducest Medical (Ducest, Mattersburg, Germany), Innovative Health Solutions (Versailles, IN, USA) and Hwato (Suzhou, Jiangsu Province, China). Despite the large number of aVNS clinical studies, clinical evidence to support a specific therapeutic outcome is often mixed, with conflicting trial results for the same physiological outcome measure (Burger et al., 2020; Keute et al., 2020).

According to the Oxford Centre for Evidence Based Medicine’s (CEBM) Levels of Clinical Evidence Scale, the highest level of clinical evidence is a systematic review of multiple high-quality double-blinded, randomized, and controlled clinical trials (RCTs) with narrow confidence intervals, each homogeneously supporting the efficacy and safety of a therapy for a specific clinical outcome (Centre for Evidence-Based Medicine, 2009). However, reaching this level of evidence is costly and time-consuming. Years of precursor clinical studies with fewer number of subjects are needed to identify the most efficacious embodiment of the therapy that can be safely delivered. Data from these precursor studies are required to design more definitive clinical studies. The field of aVNS, being relatively new clinically, is understandably still in these early phases of clinical development.

We performed a systematic review of aVNS RCTs with two primary goals: 1) to provide an accessible framework for the aVNS community to review current studies for specific outcome measures as a resource to inform future study design and 2) to perform a qualitative assessment of the current level of clinical evidence to support aVNS efficacy for the most common outcome measures reported. To this end, the Cochrane Risk of Bias Tool (Sterne et al., 2019) – a framework previously used to identify risk of bias in RCT studies of epidural spinal cord stimulation (Duarte et al., 2020) and dorsal root ganglion stimulation (Deer et al., 2020) to treat pain – was first used to assess the quality of evidence in individual aVNS RCTs. These data were aggregated to broadly assess the current level of clinical evidence, according to the Oxford CEBM scale, to support aVNS efficacy across the common physiological outcomes. Our efforts were not intended to provide a precise assessment of the current level of clinical evidence but to identify the most common gaps in clinical study design and reporting. These gaps were analyzed to identify systematic next steps that should be addressed before aVNS can move to a higher level of evidence for any specific clinical outcome.

## 2. METHODS

### 2.1 Search method

Our literature search was designed to identify reports of clinical RCTs testing aVNS as an intervention. Two databases were searched systematically: PubMed and Scopus (includes MEDLINE and Embase databases). Additionally, two search strategies were used. The first strategy combined search terms related to aVNS and RCT. The second search strategy focused on search terms related to commercial aVNS devices and their manufacturers. Complete search strings for both strategies are available in supplementary material. The search was last updated in July 2020. In addition, citations of all selected studies were searched to identify additional studies that met the inclusion criteria. The citations of relevant reviews (Murray et al., 2016; Yap et al., 2020) were also searched.

As our primary goal was to assess the effects of auricular stimulation, studies using any stimulation modality from any field, including acupuncture and electroacupuncture were initially included as long as the intervention was at the auricle. When it became evident that a meta-analysis would not be possible due to incomplete reporting of information, we decided to exclude traditional Chinese medicine (TCM) studies, which typically used acupuncture, electroacupuncture, or acupuncture beads. Studies were considered TCM studies if acupoints were used to justify location of stimulation or if they were published in a TCM journal. This is captured in Fig. 2 adapted from PRISMA (Moher et al., 2009). All studies excluded at the end of the search after full-text review are listed in supplementary material.

**Figure 2:**
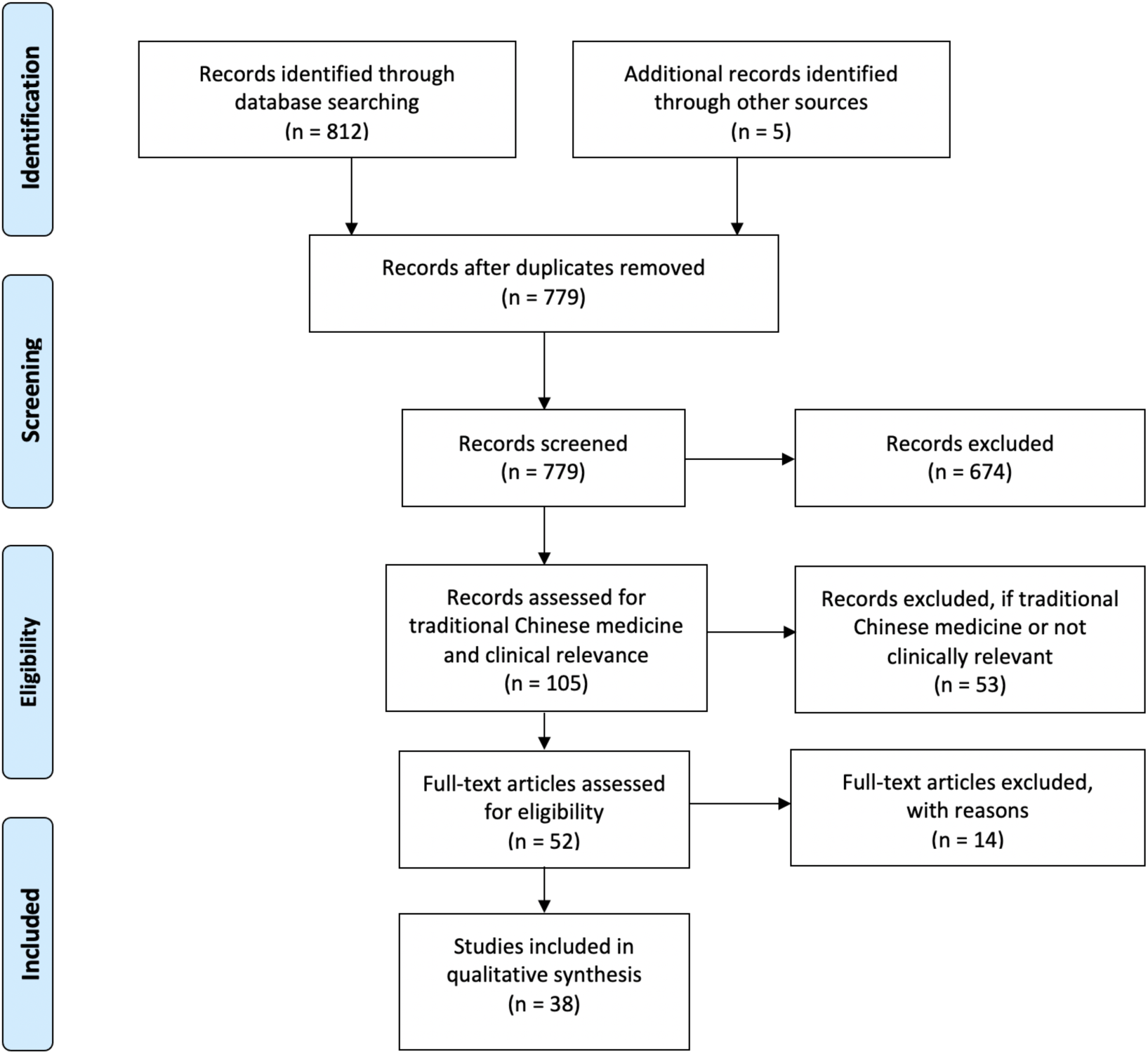
Adapted PRISMA Flow Chart.

### 2.2 Inclusion and exclusion criteria

In papers where more than one clinical trial was reported, each trial that was randomized and controlled was included in the systematic review; non-RCT portions of included publications were not analyzed. Only publications 1991 and after were included, with the cutoff marking the first time autonomic activity biomarkers were reportedly measured during auricular stimulation (Johnson et al., 1991).

Included studies had to report measurements of direct clinical significance. This exclusion partially relied on whether the study claimed direct clinical implications of their findings. Additionally, studies were excluded if the measurements did not have a well-established link to clinically significant outcomes. For instance, pupil size, functional magnetic resonance imaging (fMRI), electroencephalography (EEG), and somatosensory evoked potentials (SSEPs) are secondary physiological measures of target engagement (Burger et al., 2020). Although they may be useful to study the mechanisms of aVNS, they do not have well-established links to clinically significant outcomes. In comparison, heart rate variability (HRV), a measure of sympathovagal tone, is considered a measurement of direct clinical significance as sympathovagal imbalance is related to several disease states (Bootsma et al., 2003). Similarly, studies focused on cognitive neuroscience topics, such as behavior, learning, fear extinction, or executive functions were excluded. In contrast, psychological studies addressing addiction, depression, pain, and stress were included in the final qualitative review as they have had direct clinical significance.

### 2.3 Cochrane risk of bias 2.0 tool to assess quality of evidence

We used the Cochrane risk of bias 2.0 tool (RoB), an established tool to assess bias in clinical RCTs (Sterne et al., 2019), which has been cited over 40,000 times in Google Scholar, to evaluate the quality of evidence in each study. The RoB tool assesses bias in five subsections intended to capture the most common sources of possible bias in clinical studies. It is important to note a rating of ‘some’ or ‘high’ risk of bias does not mean that researchers conducting the study were themselves biased, or that the results they found are inaccurate. Deviations from ideal practice frequently occur due to a variety of potentially uncontrollable reasons. These deviations from the ideal just increase the chance that any stated result is a ‘false negative’ or a ‘false positive’ beyond the stated statistical convention used in the study.

For each study, the tool provides a suggested algorithm to rate bias through a series of guiding questions across the following five sections. Each section ends with a bias assignment of ‘low’, ‘some concerns’, or ‘high’. Template rubrics provided by Cochrane with the answers to these guiding questions have been included for every study evaluated in supplementary material. At several instances, the suggested algorithm was overridden by the reviewer with justification annotated on the individual rubric found in supplementary material. Below is an explanation of how each subsection was evaluated with respect to aVNS, see (Higgins et al., 2019) for more information on the recommended implementation of the Cochrane assessment tool.

#### Bias arising from the randomization process

randomization is important in a clinical study to ensure that differences in the outcome measure between the treatment and control groups were related to the intervention as opposed to an unintended difference between the two groups at baseline. To obtain a rating of ‘low’ risk of bias, the study had to 1) randomize the allocation sequence, 2) conceal the randomized sequence from investigators and subjects till the point of assignment, and 3) test for baseline differences even after randomization. The latter is essential as even in a truly randomized design, it is conceivable that randomization yields an unequal distribution of a nuisance variable across the two groups. This is more likely to occur in studies with a smaller number of participants (Kang et al., 2008), which is the sample size found in many aVNS studies. Even in studies with a crossover design – meaning participants may receive a treatment and then, after an appropriate wash-out period, receive a sham therapy – it is important to test for baseline differences and that an equal number of subjects be presented with sham or therapy first (Nair, 2019).

#### Bias due to deviations from intended interventions

During the implementation of a clinical trial it is foreseeable for several subjects who were randomized to a given group to not receive the intended intervention or for blinding to be compromised. Compromised blinding is especially pertinent in neuromodulation studies, including aVNS studies, where there may be a difference in paresthesia or electrode location between the intervention and control group (Robbins and Lipton, 2017). Marked visual or perceptual differences between intervention groups can clue investigators and subjects to become aware of the treatment or control arm assignments, thereby violating the principle of blinding and deviating from the intended intervention. In order to receive a ‘low’ risk of bias score, the study must have minimized and accounted for deviations from intended intervention due to unblinding, lack of adherence, or other failures in implementation of the intervention.

#### Bias due to missing outcome data

In conducting a clinical trial, being unable to record all intended outcome measures on all subjects is common. This can happen for a variety of reasons, including participant withdrawal from the study, difficulties in making a measurement on a given day, or records being lost or unavailable for other reasons (Higgins et al., 2019). In assessing how missing data may lead to bias it is important to consider the reasons for missing outcome data as well as the proportions of missing data. In general, if data was available for all, or nearly all participants, this measure was given ‘low’ risk of bias. If there was notable missing data that was disproportionate between the treatment and control group, or the root cause for missing data suggested there may be a systemic issue, this measure was rated ‘some concerns’ or ‘high’ risk of bias depending on severity.

#### Bias in measurement of the outcome

How an outcome was measured can introduce several potential biases into subsequent analyses. Studies in which the assessor was blinded, the outcome measure was deemed appropriate, and the measurement of the outcome was performed consistently between intervention and control groups were generally considered ‘low’ risk of bias. If the outcome assessor was not blinded but the outcome measure was justified as unlikely to be influenced by knowledge of intervention the study was also generally considered ‘low’ risk of bias.

#### Bias in selection of the reported result

An important aspect in reporting of clinical trial results is to differentiate if the data is exploratory or confirmatory (Hewitt et al., 2017). Exploratory research is used to generate hypotheses and models for testing and often includes analyses that are done at least in part retrospectively and are therefore not conclusive. Exploratory research is intended to minimize false negatives but is more prone to false positives. Confirmatory research is intended to rigorously test the hypothesis and is designed to minimize false positives. An important aspect of confirmatory research is pre-registration of the clinical trial before execution, including outlining the hypothesis to be studied, the data to be collected, and the analysis methods to be used. This is necessary to ensure that the investigators did not 1) collect data at multiple timepoints but report only some of the data, 2) use several analysis methods on the raw data in search for statistical significance, or 3) evaluate multiple endpoints without appropriate correction for multiple comparisons. Each of these common analysis errors violates the framework by which certain statistical methods are intended to be conducted – introducing an additional chance of yielding a false positive result. Studies that pre-registered their primary outcomes and used the measurements and analyses outlined in pre-registration generally scored ‘low’ risk of bias in this category.

### 2.4 Information extraction

Each paper was read in its entirety and a summary table was completed capturing study motivation, study design, study results, and critical review. Study motivation outlined the study hypothesis and hypothesized therapeutic mechanism of action if mentioned in the paper. We also noted whether implantable VNS had achieved the hypothesized effect in humans. Study design encapsulated subject enrollment information (diseased or healthy, the power of the study, and the inclusion and exclusion criteria), type of control and blinding, group design (crossover or parallel), stimulation parameters, randomization, baseline comparison, and washout periods. Lastly, study results included primary and secondary endpoints, adverse effects, excluded and missing data, and statistical analysis details (pre-registered, handling of missing and incomplete data, multiple group comparison, etc.). Study results also analyzed if the effect was due to a few responders or improvements across the broad group, worsening of any subjects, control group effect size, and clinical relevance and significance of findings. Where sufficient information was reported, standardized effect size was calculated using Hedges’ g (Turner and Bernard, 2006).

Each publication had a primary reviewer, and an additional secondary reviewer went through all papers. Any concerns raised by either reviewer were discussed as a group. If crucial basic information (e.g., which ear was stimulated, electrode used, etc.) was not reported (NR), an attempt was made to reach out to the author and if unsuccessful, to infer the information from similar studies by the group. Inferred or requested information is annotated as such. This effort helped highlight incomplete reporting of work while maximizing available information for the review to conduct an informed analysis.

## 3. RESULTS

A total of 38 articles were reviewed totaling 41 RCTs – two each in the publications by Hein et al. (2013), Cakmak et al. (2017), and Badran et al. (2018). In an initial review of the RCTs, it was apparent that a wide variety of electrode designs, stimulation parameters, study methodologies, and clinical indications were tested. As a framework by which to organize this multifaceted problem in the results below, we first discuss the electrode designs and stimulation parameters used across aVNS studies with the goal of identifying the most common aVNS implementation strategies and rationale for selection. Next, we discuss the study design features across all studies, again with the goal of identifying the most common practices. We then provide an assessment of all studies, regardless of clinical indication, using the Cochrane risk of bias (RoB) tool. Finally, we discuss the commonly measured outcomes based on treatment indication to identify which findings were most consistent across studies.

A sortable table summarizing the design and result features of every reviewed study has been included as an excel file in supplementary material to allow viewing based on specific features of interest. Design and result features have been reduced to common keywords in this spreadsheet to facilitate sorting; however, this means specific details of outcome measures have been reduced to general categories in some cases.

### 3.1 aVNS electrode Designs, configurations, and stimulation parameters across studies

Upon initial review, it was immediately evident that implementation of both active and sham varied greatly across studies. Table 1 details the electrode design, configuration (monopolar or bipolar), target location, and stimulation parameters for the active and control arms of the study. Fig. 3 shows a box plot presenting the distribution of pulse widths, stimulation current amplitudes, and frequencies used across studies. In the active arm, the interquartile range (IQR) of stimulation current amplitudes was 0.2-5 mA, pulse width was 200-500 μs, and frequency of stimulation was 10-26 Hz. It is notable that the commonly used aVNS waveform parameters are similar to the parameters typically used for stimulation of the cervical vagus at a pulse width of 250 µs and frequency of 20 Hz (LivaNova, 2017), which uses surgically implanted epineural cuff electrodes. Outside of the IQR, the spread of the parameters is wide.

**Table 1:** Electrode design, configuration, target location, and stimulation parameters (sorted by indication type, primary endpoints, and color coded RoB score)

| Author (y)<br>Indication* | Primary Endpoints | Active waveform** &<br>location | Active amplitude & electrode<br>type*** | Control |
| --- | --- | --- | --- | --- |
| Cardiac |  |  |  |  |
| Andreas (2019)<br>postop atrial<br>fibrillation | ECG post atrial<br>fibrillation | 1 Hz, PW not reported (NR),<br>40 min on 20 min off<br>Side NR Triangular fossa | Sub-sensory (1 mA), Ducrest<br>Neurostimulator V | Sham: same location,<br>low level stimulation<br>(mA NR) |
| Stavrakis (2015)<br>Atrial fibrillation | Atrial fibrillation<br>cycle length and<br>duration, TNF-α,<br>CRP | 20 Hz, 1 ms pulse width<br>(PW), on/off cycle NR<br>Right tragus | 50% of heart sinus rate slowing<br>current threshold (mA NR),<br>Grass S88 stimulator clip<br>electrodes | Placebo: no current,<br>same location |
| Stavrakis (2020)<br>Atrial Fibrillation | Atrial fibrillation<br>burden | 20 Hz, 200 μs PW, on/off<br>cycle NR<br>Right tragus | Strong sensory (1 mA below<br>mild pain threshold, mean 16.8<br>mA), Parasym clip electrodes | Sham: diff amplitude<br>(mean 19.9 mA), diff<br>location<br>(right earlobe) |
| Badran (2018)<br>Heart rate | ΔHR during<br>stimulation | 9 waveforms (1, 10, 25 Hz) x<br>(100, 200, 500 μs PW), on/off<br>cycle NR<br>Left tragus | 2x sensory threshold (at 100 μs<br>PW: $9.28 \pm 2.56$ mA, at 200 μs<br>PW: $5.32 \pm 1.60$ mA, at 500 μs<br>PW: $3 \pm 0.93$ mA), custom clip<br>electrodes | Sham: At 100 μs PW:<br>$6.57 \pm 1.83$ mA. At 200<br>μs PW: $3.64 \pm 1.26$ mA.<br>At 500 μs PW: $1.97 \pm$<br>$0.71$ mA. diff location<br>(earlobe) |
| Afanasiev (2016)<br>Coronary<br>Insufficiency and<br>left ventricular<br>dysfunction | Heart rate and 6<br>minute walk<br>distance | Frequency NR, PW NR, on/off<br>cycle NR<br>Side NR concha | Titration method NR (mean<br>$0.05$-$0.15$ mA), electrode NR | Placebo: same location<br>(concha) |
| Tobaldini (2019)<br>Orthostatic<br>stress | ΔHR, LF/HF, systolic<br>arterial BP<br>variance, R-R<br>interval pattern,<br>respiratory rate | 25 Hz, 200 ms PW, on/off<br>cycle NR, phase NR<br>Left cymba concha | First sensory (1-6 mA), NEMOS<br>ball contact electrodes | Placebo: No<br>stimulation, same<br>location. |
| Fisher (2018)<br>Hypertension | Percentage<br>decrease in median<br>systolic blood<br>pressure (SBP) | 25 Hz, 15 ms PW, 1 s<br>duration (gated to<br>exhalation), biphasic<br>Left cymba concha and<br>beneath antihelix | Strong sensory (mA NR),<br>stimulator NR, surface<br>electrodes | Placebo: no current,<br>same location |
| Stowell (2019)<br>Hypertension | Arterial blood<br>pressure | 2, 10, 25 or 100 Hz. 300 μs<br>PW, 1 s on/off<br>Left cymba concha | Strong sensory (mA NR)<br>Urostim device, custom-built<br>ergonomic electrodes | Placebo: no current,<br>same location |
| Zamotrinsky (2001) coronary artery disease | HR, BP, left ventricular (LV) diastolic function, LV filling | 3 Hz, 1500 μs PW, on/off cycle NR<br>Bilateral cavum concha | Titration method NR (0.2-1.25 mA), acupuncture needles | No intervention |
| Antonino (2017) Spontaneous cardiac baroreflex sensitivity (cBRS) | cBRS from systolic blood pressure and RR interval<br>HRV (LF/HF) | 30 Hz, 200 μs PW, on/off cycle NR<br>Bilateral tragus | Sensory threshold (10-50 mA device range), ear clip electrodes | 1 Placebo<br>1 Sham: same waveform (mA NR), diff location (bilateral earlobe) |
| Bretherton (2019) HRV and baroreflex sensitivity | cBRS and HRV | 30 Hz, 200 μs PW, on/off cycle NR<br>Side NR inner & outer tragus | Sensory threshold (2-4 mA), custom TENS electrodes | Placebo: same location (inner & outer tragus) |
| Clancy (2014) HRV & Sympathetic Activity | HRV (LF/HF) | 30 Hz, 200 μs PW, continuous,<br>Side NR inner & outer tragus | Sensory threshold (10-50 mA device range), V-TENS Plus with modified surface electrodes | Placebo: no current, same location |
| De Couck (2017) HRV | ECG with HRV | 25Hz, 250 μs PW, on/off cycle NR<br>Bilateral cymba concha | Strong sensory (mean ~0.7 mA), NEMOS ball contact electrodes | Placebo: no current, same location |
| Borges (2019) cardiac vagal activity | HRV | 25 Hz, PW of 200-300 μs, 30 s on/off<br>Left cymba concha | Set stimulation (1 mA) and strong sensory stimulation (2.5 ± 0.93 mA), NEMOS ball contact electrodes | Sham: same set stimulation, different strong sensory stimulation (2.76 ± 1.01 mA), diff location (earlobe) |
| Tran (2019) Left Ventricular Strain and Autonomic Tone | LV global longitudinal strain (GLS) | 20 Hz, 200 μs PW, on/off cycle NR<br>Right tragus | Strong sensory (1mA below pain threshold, mean 22.6 mA active), Parasym earclip electrodes | Sham: diff current (mean 21.8 mA), diff location (right earlobe) |
| Yu (2017) Myocardial Ischemia-Reperfusion Injury | Ventricular premature beat (VPB) incidence | 20 Hz, 1 ms PW, 5 s on/off<br>Right tragus | 50% of heart sinus rate slowing threshold, clip electrodes (S20 stimulator, Jinjiang, Chengdu City, China) | Placebo: no current, same location (right tragus) |
| Epilepsy |  |  |  |  |
| Bauer (2016) Epilepsy | Reduction in seizure frequency (per 28 days) | 25 Hz, 250 μs PW, on/off cycle NR<br>Left concha | Sensory threshold (0.50 ± 0.47 mA), NEMOS ball contact electrode | Sham: diff waveform 1 Hz and 1.02 ± 0.83 mA, same location (left concha) |
| Aihua (2014) Epilepsy | Reduction in seizure frequency (per month) | 20 Hz, 200 ms PW, on/off cycle NR<br>Bilateral concha and external ear canal | Pain threshold (mA NR), electrode NR | Sham: same waveform (mA NR), diff location (bilateral earlobe) |
| Rong (2014)<br>Epilepsy | Seizure frequency<br>(per 4 weeks) | 20-30 Hz, $\leq 1$ ms PW<br>Ear side NR cymba concha<br>and cavum concha | Set stim (1 mA), electrode with<br>3 carbon-impregnated silicone<br>tips (Suzhou Medical Appliance<br>Co. Ltd.) | Sham: same waveform,<br>diff location; contacts<br>at scapha and<br>antihelical fold |
| Pain |  |  |  |  |
| Straube (2015)<br>Chronic Migraine | Decrease in<br>headache per 28<br>days | 25 Hz, 250 μs PW, 30 s on/off<br>Left concha | Strong sensory (mA NR),<br>NEMOS ball contact electrodes | Sham: 1 Hz frequency,<br>same location |
| Janner (2018)<br>Pain | Perceived pain<br>intensity and<br>temporal<br>summation of pain | 100 Hz, 200 μs PW, 0.01 s<br>on/0.49 s off<br>Bilateral cymba concha | Strong sensory (mA NR),<br>custom earplug electrodes<br>wrapped in NaCl-soaked wool | Sham: same waveform,<br>diff location (earlobe).<br>Placebo: same<br>location, no current |
| Kovacic (2017)<br>GI pain | Change in max<br>abdominal pain<br>intensity and<br>composite of Pain-Frequency-Severity-Duration<br>scale | Alternating 1 Hz and 10 Hz<br>every 2 s, 1 ms PW, 2 hrs<br>on/off<br>Side NR Earlobe, triangular<br>fossa, ventral periauricular<br>tragus | Sub-sensory (mA NR), 2 mm<br>titanium percutaneous<br>electrodes (monopolar) | Sham: no current (sub<br>sensory like active),<br>same location |
| Kutlu (2020)<br>Fibromyalgia | Visual Analog Scale,<br>Beck Depression<br>Scale, Beck Anxiety<br>Scale, Fibromyalgia<br>Impact<br>Questionnaire,<br>Short Form-36 for<br>life quality | 10 Hz, $<500$ μs PW, biphasic<br>asymmetrical<br>Bilateral tragus and concha | First sensory (mA NR), custom<br>designed surface electrodes | Only exercise (active<br>group is exercise and<br>aVNS) |
| Busch (2013)<br>Pain | Thermal,<br>mechanical, and<br>pressure pain<br>thresholds | 25 Hz, 250 μs PW, on/off<br>cycle NR<br>Left concha | First sensory ($1.6 \pm 1.5$ mA),<br>STV02 (Cerbomed) electrodes | Placebo: no current,<br>same location |
| Juel (2017) Pain<br>&<br>Gastrointestinal<br>Motility | ECG and PPG,<br>mechanical pain<br>threshold, cold<br>pressor test, and<br>drink test<br>(ultrasound<br>imaging) | 30 Hz, 250 μs PW,<br>continuous<br>Left concha | Pain/uncomfortable (intensity<br>increased to counteract<br>habituation, device rated<br>between 0.1 and 10 mA),<br>NEMOS ball contact electrodes | Sham: same waveform,<br>diff location (left<br>earlobe) |
| Frøkjær (2016)<br>Gastroduodenal<br>motility and pain<br>threshold | ECG and PPG,<br>mechanical pain<br>threshold, cold<br>pressor test, and<br>drink test<br>(ultrasound<br>imaging) | 30 Hz, 250 μs PW,<br>continuous<br>Left concha | Strong sensory (1.07 mA),<br>NEMOS ball contact electrode | Sham: diff amplitude<br>(mean 1.57 mA), diff<br>location (left earlobe) |
| Napadow (2012)<br>Pain | Mechanical deep-tissue pain intensity rating, and temporal pain summation | 30 Hz, 450 μs PW, 0.5 s on/off, gated to exhalation phase of respiration<br>Left cymba concha and antihelix/cavum concha slope | Strong sensory (mA NR), modified press-tack electrodes (0.20 x 1.5 mm) | Sham: same waveform, diff location (left earlobe) |
| Johnson (1991)<br>Pain threshold and autonomic function | Electrical pain threshold and autonomic function | 100 Hz pulses every 2.4 Hz, 10 ms PW, 10 ms on/off, Right concha | Strong sensory (mA NR), carbon rubber electrodes | Sham: same waveform, diff location (antitragus) |
| Laqua (2014)<br>Pain | Electrical pain threshold | Alternating between 2 Hz and 10 Hz, 200 μs PW, on/off cycle NR<br>Bilateral concha (anode) and mastoid (cathode) | Strong sensory (mA NR), silver EEG electrode at anode and PEGG electrode at cathode | Placebo: subsensory stim, same location |
| Psychological |  |  |  |  |
| Hein (2013)<br>Depression | Hamilton Depression Rating Scale, Beck's Depression Inventory | 1.5 Hz, PW NR, on/off cycle NR<br>Bilateral concha | Sub-sensory (0-600 μA device range in study 1 and 130 μA in study 2), TENS-2000 (study 1) and TENS-1000 (study 2) Auri-Stim Medical, Inc. with corresponding electrodes (4 contacts) | Placebo: no current, same location |
| Hasan (2015)<br>Schizophrenia | Positive and negative schizophrenia symptom scale | 25 Hz, 30 s on/180 s off, 250 μs PW, phase NR<br>Left outer ear canal | Strong sensory (mA NR), CM02 (Cerbomed) titan electrodes | Placebo: no current, same location |
| Burger (2019)<br>Negative Thought Occurrence | Number of negative thought intrusions | 25 Hz, 250 μs PW, 30 s on/off<br>Left cymba concha | Set (0.5 mA), NEMOS ball contact electrodes | Sham: same waveform, diff location (left earlobe) |
| Others |  |  |  |  |
| Addorisio (2019)<br>Rheumatoid Arthritis | Endotoxin-induced interleukin (IL)-6, IL-1β, and TNF | ~160 Hz vibrations, Right cymba concha | NA (vibratory device) | Sham: same waveform diff location (right gastrocnemius) |
| Salama (2020)<br>Acute inflammatory response after lung lobectomy | CRP, IL6, IL10, IL-1B, IL-18, TNF-a | 1 Hz, 200 μs PW, 40 min on 20 min off<br>Side NR Triangular Fossa | First sensory (230 nA), ~2 mm needles | No intervention |
| Huang (2014)<br>Impaired glucose tolerance | 2-hr plasma glucose levels | 20 Hz, <= 1 ms PW, phase NR<br>Side NR concha | Set (1 mA, intensity adjusted based on tolerance of subjects), electrodes similar to Rong (2014) | Sham: same waveform, diff location (superior scapha) |
| Cakmak (2017)<br>Parkinson's | Motor examination (part III of the unified Parkinson's disease rating scale (UPDRS)) | 130 Hz, 100 μs PW, continuous, biphasic<br>Ipsilateral ear to dominant motor symptoms in tragus, antitragus, and helix minor muscles | Strong sensory (100-130 μA), percutaneous electrodes | Placebo: no current, 2 percutaneous electrodes in upper helix |
| Maharjan (2018)<br>olfactory function | Odor threshold test and supra-threshold test | 80 Hz or 10 Hz, 180 μs PW, on/off cycle NR<br>Left internal (concha) & external ear | Pain threshold (0.1 - 10 mA), electrode NR | Sham: same waveform, diff location (left earlobe) |
| Tutar (2020)<br>Tinnitus | Tinnitus Handicap Inventory and Depression Anxiety Stress Scales | 200 Hz, 1 ms PW, on/off cycle NR<br>Unilateral and bilateral cyma concha | First sensory (10-30 mA), silver electrodes (Provile TENS stimulator) (monopolar) | Placebo: same location |
| *Risk of Bias score indicated by box color: red for 'high', orange for 'some concerns', and green for 'low' risk of bias.<br>**Monophasic if phase not otherwise specified.<br>***Bipolar if electrode polarity not otherwise specified. Configuration considered monopolar only if the return electrode is distant enough not to activate the target region.<br>A list of abbreviations is found in supplementary material. |  |  |  |  |

**Figure 3:**
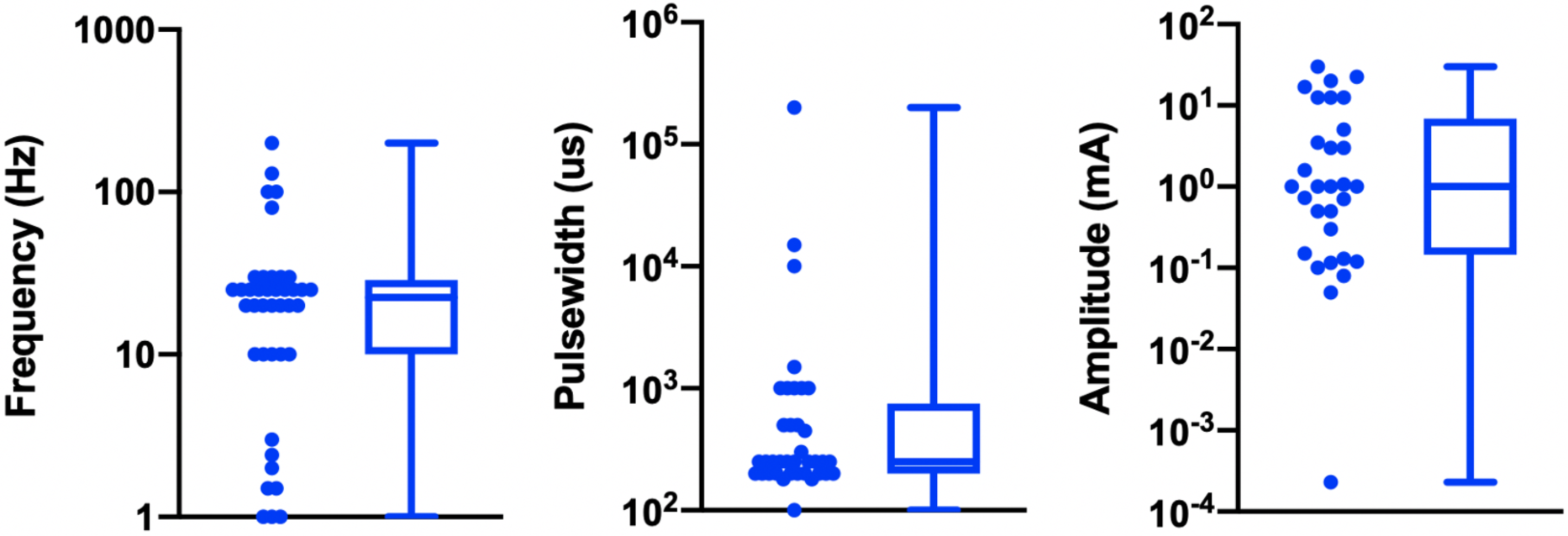
Visualization of reported stimulation waveform parameters in 41 reviewed aVNS RCTs.

The large variation in waveform parameters is indicative of the exploratory nature of aVNS studies and underscores the difficulties in comparisons across studies where similar indications use widely different parameters. The variations in pulse width and stimulation frequency are due to the range of values chosen by investigators. The variations in stimulation amplitude are more nuanced and are discussed later in this section.

Table 1 presents neuromodulation device parameters and is organized by indication type, then primary endpoints, then RoB score. Data is organized in the following columns:

#### Primary endpoints

The main result of clinical interest. Studies are grouped by endpoints measured within their respective indications. For example, within the cardiac diseases indication the studies investigating inflammatory cytokine levels are located adjacent to each other.

#### Active waveform & location

frequency, pulse width (PW), on/off cycle duration (duty cycle), and stimulation location.

#### Active amplitude & electrode type

current amplitude, titration method used to reach that amplitude, and electrode type and stimulator model when available. Titration methods are denoted as sub-sensory, first sensory, strong sensory (not painful), painful, or set at a particular amplitude. These terms reflect the cues that investigators used (Badran et al., 2019) to determine the stimulation amplitude for each subject:

##### Sub-sensory titration

Stimulation was kept just below the threshold of paresthesia sensation.

##### First sensory titration

Subject is barely able to feel a cutaneous sensation.

##### Strong sensory titration

Subject feels a strong, but not painful or uncomfortable sensation from the stimulation.

##### Pain titration

Stimulation amplitude is increased until the subject feels a painful or uncomfortable sensation.

##### Set stimulation

Fixed amplitude across all subjects – resulting in different levels of sensation due to the individual’s unique anatomy and perception.

#### Control

control group stimulation amplitude, control design (sham or placebo), and stimulation location. Following Duarte et al. (2020), we defined sham as when the control group experience from the subject perspective is identical to the active group experience – including paresthesia and device operating behavior. Conversely, placebo control is defined when the control group subjects do not experience the same paresthesia, device operation, or clinician interaction as the active group subjects.

As noted previously, frequency and pulse width are grouped around 10-26 Hz and 200-500 μs, respectively. Stimulation amplitude values are concentrated around 0.2-5 mA. Further statistics on these parameters are presented in table 2.

**Table 2:** Summary statistics of stimulation parameters

| Stimulation Parameter | Median | IQR |
| --- | --- | --- |
| Amplitude (mA) | 1.0 | 0.19 - 4.7 |
| Pulse Width (μs) | 250 | 200 - 500 |
| Frequency (Hz) | 22.5 | 10.0 - 26.3 |

Fig. 3 presents the data from table 2 in box plot form. Illustrated here are the interquartile ranges, maximums, minimums, and medians of the stimulation waveform parameters, including extreme cases. Studies that report ranges for parameters are included as a single value representing the average of the boundaries of that range.

While the large variation in pulse width and frequency parameters can be explained as choices made by investigators, the sources of the large variation in stimulation current amplitude is not as trivial. It is important to consider differences in electrode design, material, area, and stimulation polarity when comparing stimulation current amplitudes across studies. This is because electrode geometry and contact area have the potential to impact target engagement of underlying nerves (Poulsen et al., 2020). Furthermore, nerve activation is a function of current density at the stimulating electrode (Rattay, 1999), and it is not possible to accurately estimate current density without knowing electrode geometry. Another source of variability arises from the fact that different studies used different titration methods to determine stimulation current amplitude. In the studies reviewed, current amplitude was often calibrated to different levels of paresthesia perception. The level of paresthesia subjects feel is related to current density, which is once again related to stimulation current through electrode geometry.

In order to determine an optimal stimulation paradigm, target engagement must be thoroughly quantified with respect to the aforementioned variables. Direct measures of target engagement of the nerve branches exiting the auricle will further our understanding of optimal stimulation parameters (see section 4.3 on directly measuring local neural target engagement).

### 3.2 aVNS trial designs across studies

The way in which studies were designed also varied greatly. Of the 41 RCTs reviewed, 20 used a crossover design while 21 opted for a parallel design. In terms of control group design, 19 studies used a sham, 17 a placebo, 2 used both sham and placebo, and 3 had no intervention as control. Studies also varied in duration: 12 were chronic and 29 were acute. The differences between these study design methods are important to emphasize and explored in section 4.4.

Table 3 summarizes study designs and is organized by indication type, then primary endpoints, then RoB score. Data is organized in the following columns:

**Table 3:** Study designs (sorted by indication type, primary endpoints, and color coded RoB score)

| Author (y)<br>Indication* | Primary Endpoints | Subjects analyzed | Control | Design |
| --- | --- | --- | --- | --- |
| Cardiac |  |  |  |  |
| Andreas (2019) postop atrial fibrillation | ECG post atrial fibrillation | 40, patients undergoing cardiac surgery | Sham | Parallel, acute (1 hr) |
| Stavrakis (2015) Atrial fibrillation | Atrial fibrillation cycle length and duration, TNF-a, CRP | 40, paroxysmal AF ablation patients | Placebo | Parallel, acute (1 hr) |
| Stavrakis (2020) Atrial Fibrillation | Atrial fibrillation burden | 53, diagnosed with Paroxysmal AF | Sham | Parallel, chronic (6 months) |
| Badran (2018) Heart rate | ΔHR during stimulation. | 35, Healthy | Sham | Crossover, acute (duration NR) |
| Afanasiev (2016) Coronary Insufficiency and left ventricular dysfunction | Heart rate and 6 mins walk distance. | 70, coronary insufficiency/LV dysfunction patients | Placebo | Parallel, acute (~1 hr) |
| Tobaldini (2019) Orthostatic stress | ΔHR, LF/HF, systolic arterial BP variance, R-R interval pattern, respiratory rate | 13, Healthy | Placebo | Crossover, acute (~30 mins) |
| Fisher (2018) Hypertension | Percentage decrease in median systolic blood pressure (SBP) | 10, Hypertensive | Placebo | Crossover, acute (duration NR) |
| Stowell (2019) Hypertension | Arterial blood pressure | 12, diagnosed with Primary Hypertension | Placebo | Crossover, acute (5 days) |
| Zamotrinsky (2001) coronary artery disease | HR, BP, LV diastolic function, LV filling | 18. patients with stable angina pectoris class IV | No intervention | Parallel, acute (~10 days) |
| Antonino (2017) Spontaneous cardiac baroreflex sensitivity (cBRS) | cBRS from systolic blood pressure and RR interval<br>HRV (LF/HF) | 13, Healthy | Both placebo and sham | Crossover, acute (duration NR) |
| Bretherton (2019) HRV and baroreflex sensitivity | cBRS and HRV | 14, Healthy | Placebo | Crossover, acute (1 week) |
| Clancy (2014) HRV & Sympathetic Activity | HRV (LF/HF) | 48, Healthy | Placebo | Parallel, acute (~15 mins) |
| De Couck (2017) HRV | ECG with HRV | 30, Healthy | Placebo | Crossover, acute (duration NR) |
| Borges (2019) cardiac vagal activity | HRV | 60, Healthy | Sham | Crossover, acute (duration NR) |
| Tran (2019) Left Ventricular Strain and Autonomic Tone | Left ventricular (LV) global longitudinal strain (GLS) | 24, diagnosed with diastolic dysfunction by echocardiogram | Sham | Crossover, acute (duration NR) |
| Yu (2017) Myocardial Ischemia-Reperfusion Injury (MIRI) | Ventricular premature beat (VPB) incidence | 95, MIRI patients | Placebo | Parallel, acute (~2.5 hrs) |
| Epilepsy |  |  |  |  |
| Bauer (2016) Epilepsy | Reduction in seizure frequency (per 28 days) | 58, Epileptic | Sham | Parallel, chronic (20 weeks) |
| Aihua (2014) Epilepsy | Reduction in seizure frequency (per month) | 47, Epileptic | Sham | Parallel, chronic (12 months) |
| Rong (2014) Epilepsy | Seizure frequency (per 4 weeks) | 144, Epileptic | Sham | Parallel, chronic (24 weeks) |
| Pain |  |  |  |  |
| Straube (2015) Chronic Migraine | Decrease in headache (per 28 days) | 46, chronic migraine patients | Sham | Parallel, chronic (12 weeks) |
| Janner (2018) Pain | Perceived pain intensity and temporal summation of pain | 49, Healthy | Both placebo and sham | Crossover, acute (8 days) |
| Kovacic (2017) GI pain | Change in max abdominal pain intensity and composite of Pain-Frequency-Severity-Duration scale | 104, children with GI pain | Sham | Parallel chronic (4 weeks) |
| Kutlu (2020) Fibromyalgia | Visual Analog Scale, Beck Depression Scale, Beck Anxiety Scale, Fibromyalgia Impact Questionnaire, Short Form-36 for life quality | 52, Fibromyalgia patients | No intervention | Parallel, chronic (4 weeks) |
| Busch (2013) Pain | Thermal, mechanical, and pressure pain thresholds | 48, Healthy | Placebo | Crossover, acute (2 days) |
| Juel (2017) Pain & Gastrointestinal Motility | ECG and PPG, mechanical pain threshold, cold pressor test, and drink test (ultrasound imaging) | 20, chronic pancreatitis patients | Sham | Crossover, acute (7 days) |
| Frøkjær (2016) Gastroduodenal motility and pain threshold | ECG and PPG, mechanical pain threshold, cold pressor test, and drink test (ultrasound imaging) | 18, Healthy | Sham | Crossover, acute (~6 days) |
| Napadow (2012) Pain | Mechanical deep-tissue pain intensity rating, and temporal pain summation | 15, Chronic pelvic pain due to endometriosis patients | Sham | Crossover, acute (~1 week) |
| Johnson (1991) Pain threshold and autonomic function | Electrical pain threshold and autonomic function | 24, Healthy | Sham | Parallel, acute (~15 mins) |
| Laqua (2014) Pain | Electrical pain threshold | 21, Healthy | Placebo | Crossover, acute (~1 week) |
| Psychological |  |  |  |  |
| Hein (2013) Depression | Hamilton Depression Rating Scale, Beck's Depression Inventory | 37, Majorly depressed | Placebo | Parallel, chronic (~2 weeks) |
| Hasan (2015) Schizophrenia | Positive and negative schizophrenia symptom scale | 17, Schizophrenic | Placebo | Parallel, chronic (26 weeks) |
| Burger (2019) Negative Thought Occurrence | Number of negative thought intrusions | 97, High worriers | Sham | Parallel, acute (~20 mins) |
| Others |  |  |  |  |
| Addorizio (2019) Rheumatoid Arthritis | Endotoxin-induced interleukin (IL)-6, IL-1β, and TNF | 19, Healthy | Sham | Crossover, acute (duration NR) |
| Salama (2020) Acute inflammatory response after lung lobectomy | CRP, IL6, IL10, IL-1B, IL-18, TNF-a | 100, lobectomy via thoracotomy in patients with non-small cell lung cancer | No intervention | Parallel, acute (5 days) |
| Huang (2014) Impaired glucose tolerance (IGT) | 2-hr plasma glucose levels | 102, IGT patients | Sham | Parallel, chronic (12 weeks) |
| Cakmak (2017) Parkinson's | Motor symptoms | 24, Parkinson's Hoehn & Yahr stage 2-3 | Placebo | Crossover, acute (duration NR) |
| Maharjan (2018) olfactory function | Odor threshold test and supra-threshold test | 18, Healthy | Sham | Crossover, acute (7 days) |
| Tutar (2020) Tinnitus | Tinnitus Handicap Inventory and Depression Anxiety Stress Scales | 60, 20 per group, each with constant tinnitus >3-month duration | Placebo | Parallel, chronic (~1 month) |
| *Risk of Bias score indicated by box color: red for 'high', orange for 'some concerns', and green for 'low' risk of bias.<br>A list of abbreviations is found in supplementary material. |  |  |  |  |

#### Primary endpoints

The main result of clinical interest. Studies are grouped by endpoints measured within their respective indications. For example, within the cardiac diseases indication the studies investigating inflammatory cytokine levels are located adjacent to each other.

#### Subjects analyzed

sample size and whether subjects were healthy or diseased.

#### Control

Following Duarte et al. (2020), we defined sham as when the control group experience from the subject perspective is identical to the active group experience – including paresthesia and device operating behavior. Conversely, placebo control is defined when the control group subjects do not experience the same paresthesia, device operation, or clinician interaction as the active group subjects.

#### Design

study type (parallel or crossover), study time scale (acute or chronic), and intervention duration. Studies were classified as parallel if they randomized participants to intervention arms and each subject was assigned to only one intervention arm. Studies were classified as crossover if each subject group received every treatment but in a different order from the other subject groups (Nair, 2019). In some instances, the initial experimental group remained on the same intervention for the course of the study, while the control group was switched to the experimental intervention. These studies were classified as parallel, since not every subject received both interventions. Studies were classified as acute or chronic based on their duration being shorter or longer than 30 days, respectively.

### 3.3 Risk of bias tool to assess quality of evidence

The Cochrane 2.0 Risk of Bias (RoB) assessment subscores for each study are summarized in table 4. Explanations for each RoB subscore assignment (L = low, S = some concerns, and H = high) can be found generally explained in section 2.3 and specifically explained for each study reviewed in supplementary material. The RoB tool provides a suggested algorithm to determine overall score based on the subscores of all sections. In several instances, the suggested algorithm was overridden by the reviewer with justification annotated on the individual rubric found in supplementary material.

**Table 4:**
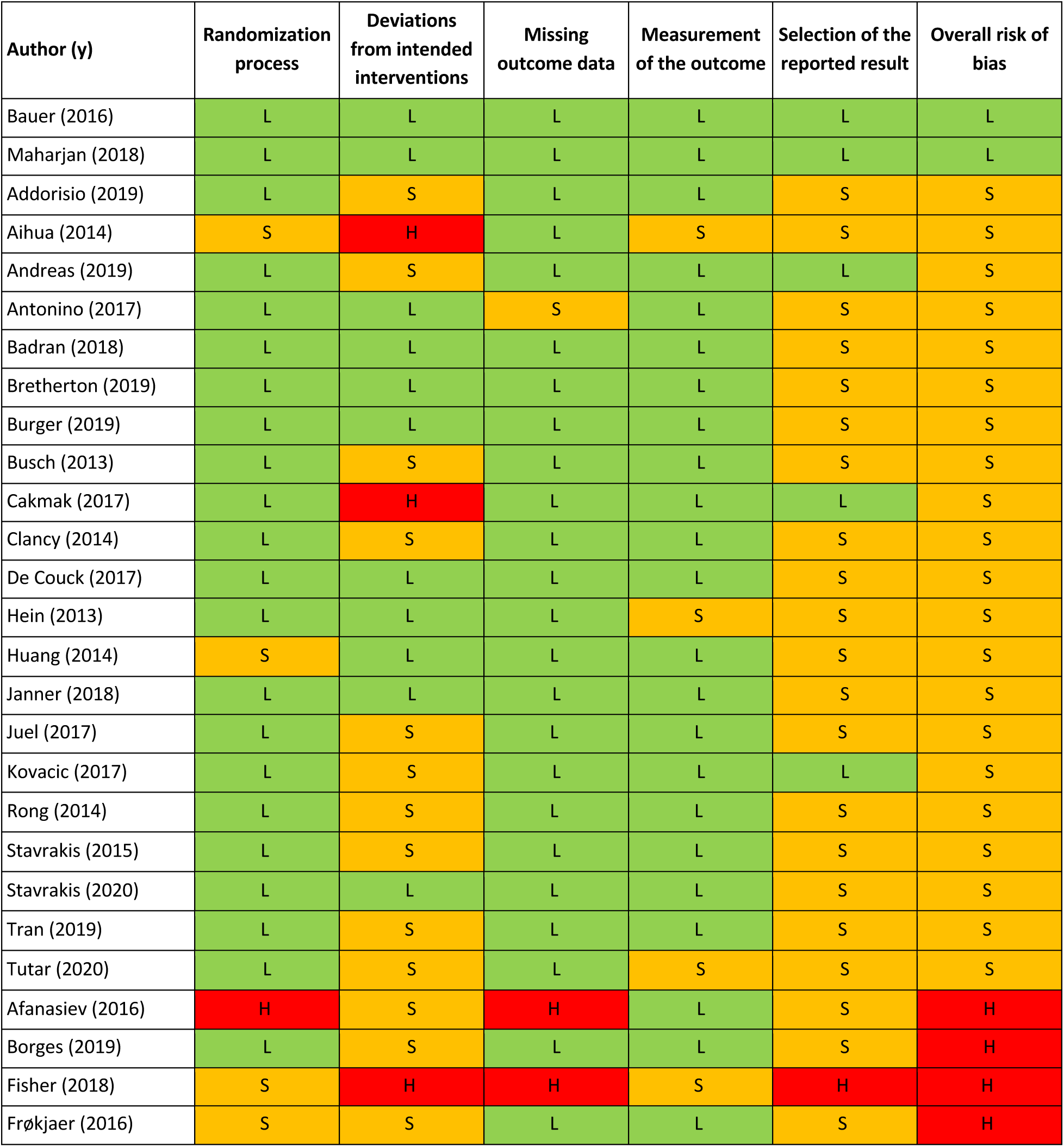
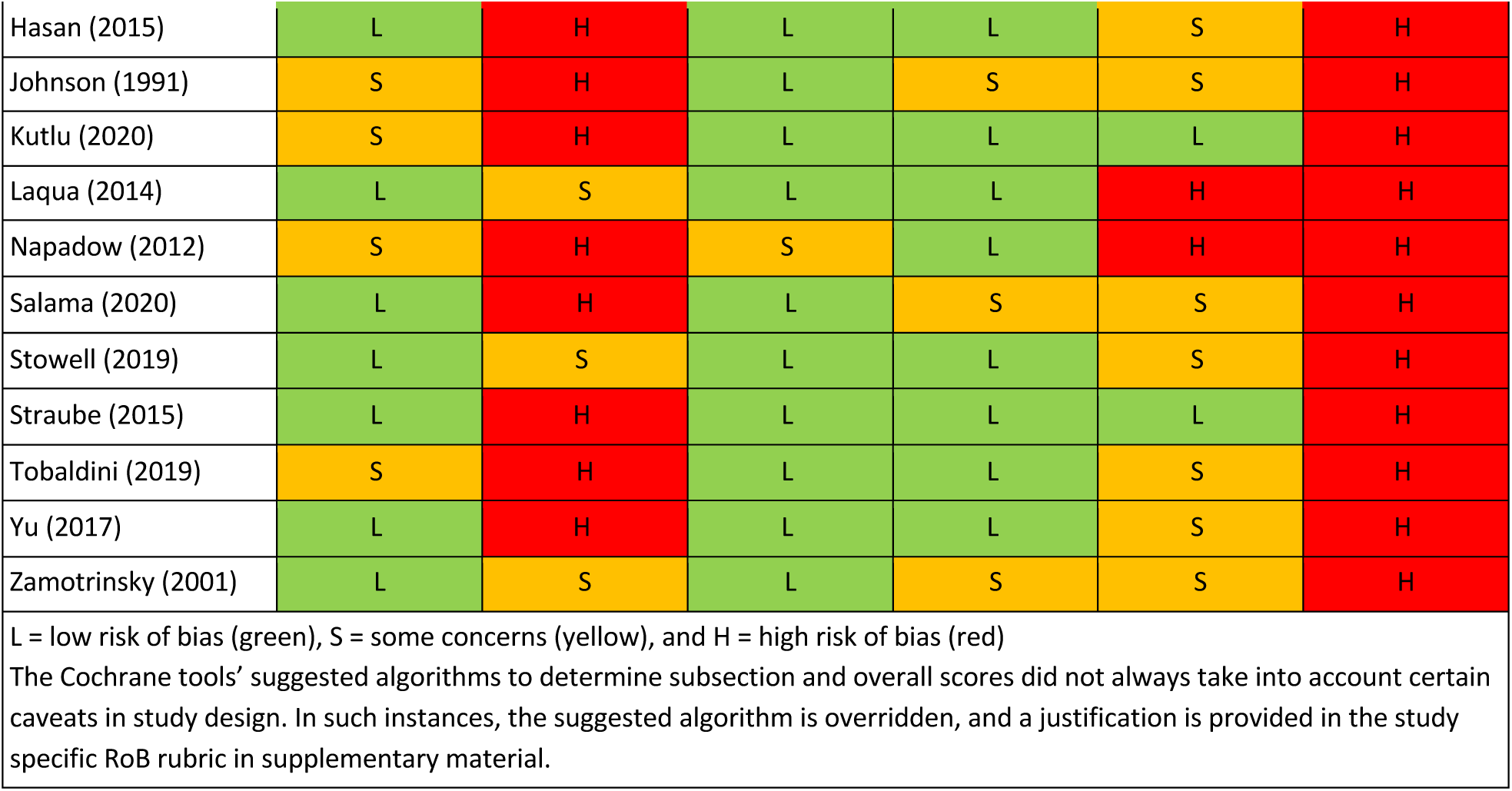
RoB overall and section scores (sorted by overall score)

Only two studies (Bauer et al., 2016 and Maharjan et al., 2018) were assigned an overall ‘low’ risk of bias. This is unsurprising, as the risk of bias assessment is rigorous and aVNS studies are in the less rigorous exploratory stages of investigation. Subsection and overall score percentages are illustrated in Fig. 4.

**Figure 4:**
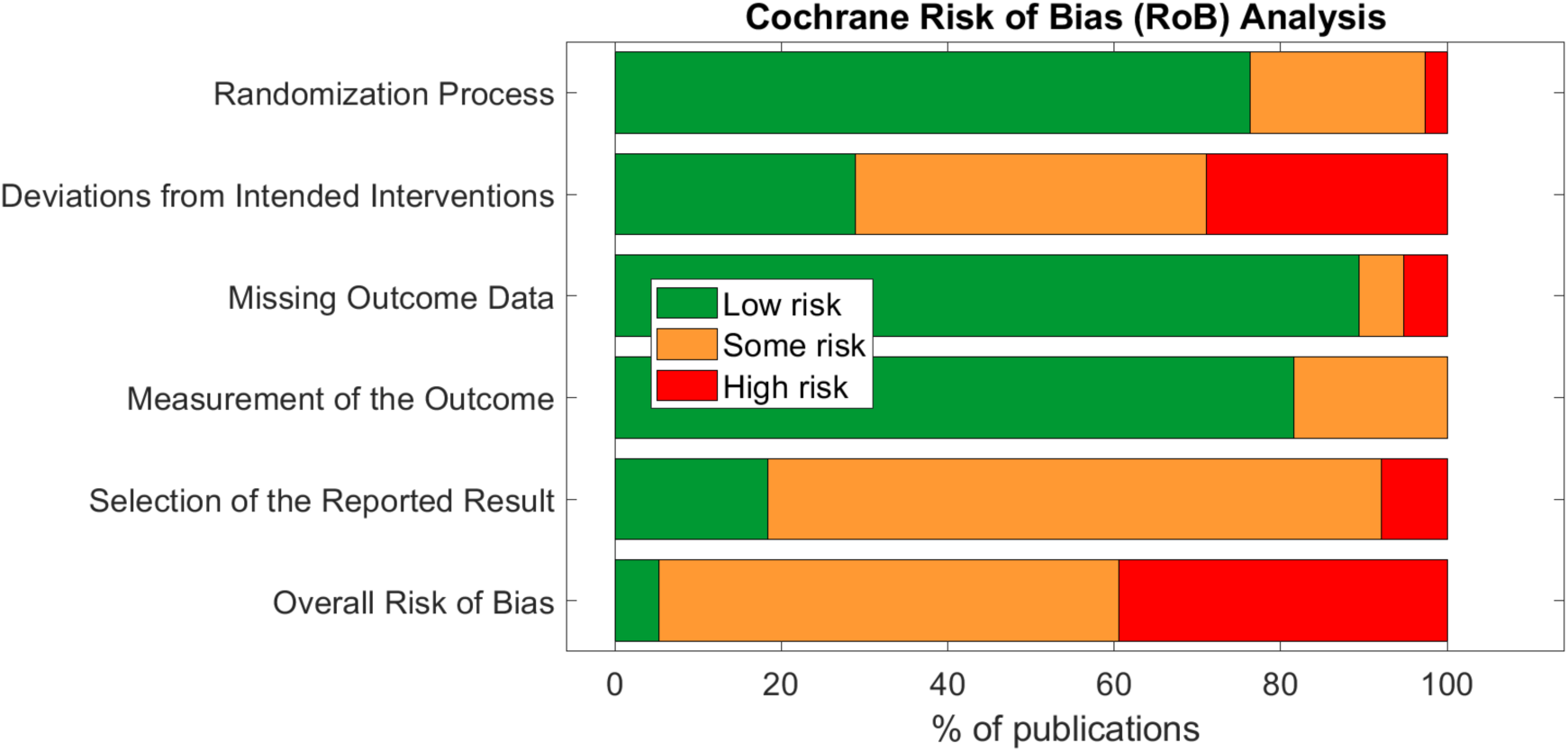
Summary of Cochrane Risk of Bias subsections and overall scores for 38 publications reviewed.

The subsection ‘randomization process’ was one of the best-scoring sections, in part because we assumed randomization was concealed from study investigators and subjects, even if the methodology to do so was not explicit. The studies that scored poorly in this section did not check for baseline imbalances between randomized groups or had baseline imbalances suggesting possible issues with the randomization method.

Notably, the ‘deviations from intended interventions’ subsection tended to have the highest risk of bias. This was mainly due to issues with potential subject unblinding as a result of easily perceptible differences between active and control groups. For example, in a crossover design, the subject experiences both the paresthesia-inducing active intervention and the non-paresthesia-inducing placebo control. The difference in paresthesia may unblind the subject to the identity of the active versus control interventions.

The subsection ‘missing outcome data’ was generally scored as ‘low’ risk of bias across studies. Studies scoring ‘high’ or ‘some concerns’ had unreported outcome data with a non-trivial difference in the proportion of missing data between interventions.

The ‘measurement of the outcome’ subsection also generally scored a ‘low’ risk of bias across studies. In order to score ‘some concerns’ or worse, outcome assessor blinding to subject intervention had to be compromised. For a ‘high’ RoB score in this section, studies measured endpoints that could be influenced by investigator unblinding, such as investigator-assessed disease evaluation questionnaires. Empirical measurements such as heart rate and blood pressure were less susceptible to this kind of bias and hence scored better.

The subsection with the highest risk for bias (‘high’ or ‘some concerns’ scores) was ‘selection of the reported results’. This was primarily due to a lack of pre-registration in most studies. Suggestions to improve study reporting are listed in section 4.1.

### 3.4 Summary of outcome measures across indications

The RoB analysis revealed potential for bias in the outcomes of the studies reviewed. Here, the studies are grouped by common indications – cardiac, inflammatory, epilepsy, and pain. The outcome of the studies for each indication are qualitatively synthesized and couched in the findings of the RoB assessment. Findings from the RoB assessment are used to point out instances where trial design or reporting may have influenced interpretation of the trial outcomes.

#### Cardiac related effects of aVNS

The most common cardiac effects assessed were changes to heart rate (HR) and sympathovagal balance. To measure sympathovagal balance, heart rate variability (HRV) was used. Results conflicted across studies for heart rate changes and sympathovagal balance but suggest aVNS may have an effect on both. However, there are concerns that these results may be attributed to trial design and inconsistent measurement methods.

Studies that reported a change in HR measured a modest mean drop of 2-3 beats per minute (BPM) in the active group. However, almost half of the eleven trials reporting HR effects reported no significant difference in effect between or within control and active stimulation. Stavrakis et al. (2015) and Yu et al. (2017) attained a consistent decrease in HR in every subject by increasing the stimulation amplitude until a decrease in HR was measured. They reported a mean stimulation threshold to elicit a HR decrease that was above the mean threshold for discomfort. In addition, Frøkjaer et al. (2016) and Juel et al. (2017) reported a significant decrease in HR during sham at the earlobe but not during active stimulation at the conchae and tragus. This suggests that decrease in HR may not be vagally mediated but perhaps mediated by the trigeminal or cervical nerve branches in the auricle (see Fig. 1a). Taken together, there is evidence for the effects of auricular stimulation on decreasing HR at high stimulation amplitudes, but it is uncertain if this decrease is mediated by the auricular branch of the vagus.

HRV, used as a measure of sympathovagal balance, was quantified inconsistently across studies and may not be an accurate indicator of whole body sympathovagal balance. Shown at the bottom of table 5 are multiple ways to analyze electrocardiogram (ECG) data for HRV. Different ways to quantify HRV enables multiple comparisons – which were often not appropriately corrected for – in search of statistical significance. HRV was calculated differently across studies making it difficult to uniformly draw conclusions across the aggregate of studies. Furthermore, HRV is not a measure of whole body sympathovagal tone, but of cardiac vagal activity – it relies on the physiological variance in HR with breathing. More variance in HR during breathing indicates more vagal control and a corresponding shift in cardiac sympathovagal balance to parasympathetic (Goldberger, 1999). Contradictory results on the parasympathetic effects of aVNS indicates that either the effects of aVNS on HRV are inconsistent, or that HRV is an unreliable measure of cardiac sympathovagal balance (Bootsma et al., 2003; Billman, 2013; Hayano and Yuda, 2019), or both. Overall, the effects of aVNS on sympathovagal balance conflict between studies. Similarly, Wolf et al. (2021), in a meta-analysis pre-print, concluded that there was “no support for the hypothesis that HRV is a robust biomarker for acute [aVNS]”.

**Table 5:** Summary of cardiac (heart rate and heart rate variability) studies

| Author (y)<br>Indication* | HR Results | HRV Results | Active<br>stimulation<br>level** | Active<br>waveform | Active location | Subjects<br>analyzed,<br>Disease |
| --- | --- | --- | --- | --- | --- | --- |
| Antonino (2017)<br>Spontaneous<br>cardiac<br>baroreflex<br>sensitivity (cBRS) | Active: 2-3 BPM<br>decrease<br>Sham: not significant | Active: LF/HF<br>parasympathetic<br>Sham at earlobe: LF/HF<br>parasympathetic<br>Placebo: LF/HF sympathetic | First<br>sensory | 30 Hz, 200 μs<br>PW | Tragus<br>(bilateral) | 13, Healthy |
| Badran (2018)<br>Heart rate | Active: ~2.40 BPM decrease<br>Sham: not significant | NA | 2x sensory threshold | 10 Hz, 500 μs PW | Tragus (left) | 35, Healthy |
| Bretherton (2019) HRV and baroreflex sensitivity | NA | Active: LF/HF parasympathetic<br>Placebo: not significant | Sensory threshold | 30 Hz, 200 μs PW | Inner & outer tragus (side not reported) | 14, Healthy |
| Burger (2019) Negative Thought Occurrence | NA | Active: not significant (RMSSD)<br>Sham: not significant | Strong sensory | 25 Hz, 250 μs PW | Left cymba concha | 97, High worriers |
| Clancy (2014) HRV & Sympathetic Activity | Active: HR decreased (data not reported)<br>Placebo: not significant | Active: LF/HF parasympathetic<br>Placebo: not significant | First sensory | 30 Hz, 200 μs PW | Inner & outer tragus (side not reported) | 48, Healthy |
| De Couck (2017) HRV | NA | Active: SDNN parasympathetic; no significant changes in LF/HF, LF, HF, RMSSD<br>Placebo: SDNN parasympathetic; no significant changes in LF/HF, LF, HF, RMSSD | Strong sensory | 25Hz, 250 μs PW | Bilateral cymba concha | 30, Healthy |
| Janner (2018) Pain | No significant effects of aVNS on HR (data not reported) | NA | Strong sensory | 100 Hz, 200 μs PW | Cymba concha (bilateral) | 49, Healthy |
| Juel (2017) Pain & Gastrointestinal Motility | Active: not significant<br>Sham: ~2.8 BPM decrease, at earlobe | Active: RMSSD parasympathetic relative to sham, sympathetic relative to baseline<br>Sham at earlobe: RMSSD sympathetic | Pain | 30 Hz, 250 μs PW | Concha (left) | 20, chronic pancreatitis patients |
| Tran (2019) Left ventricular strain and autonomic tone (HRV) | No significant effects of aVNS on HR (data not reported) | Active: SDNN no change, RMSSD parasympathetic, pNN50 sympathetic, LF/HF parasympathetic<br>Sham at earlobe: SDNN parasympathetic, RMSSD parasympathetic, pNN50 no change, LF/HF sympathetic | Strong sensory | 20 Hz, 200 μs PW | Tragus (right) | 24, diagnosed with diastolic dysfunction by ECG |
| Yu (2017) Myocardial Ischemia-Reperfusion Injury (MIRI) | HR decrease in every subject (raw HR decrease data not reported) | NA | Sinus rate slowing threshold | 20 Hz, 1 ms PW | Tragus (right) | 95, MIRI patients |
| Borges (2019)<br>cardiac vagal activity | NA | Active: RMSSD sympathetic between resting and second half stim<br>Sham: not significant | Strong sensory | 25 Hz, PW of 200– 300 μs | Left cymba concha | 60, Healthy |
| Frøkjær (2016)<br>Gastroduodenal motility and pain threshold | Active: not significant<br>Sham: ~2.2BPM decrease, at earlobe | Active: RMSSD parasympathetic at 20 min and sympathetic at 25 min<br>Sham at earlobe: RMSSD sympathetic at 20 min and parasympathetic at 25 min | Strong sensory | 30 Hz, 250 μs PW | Concha (left) | 18, Healthy |
| Johnson (1991)<br>Pain threshold and autonomic function | No significant effects of aVNS on HR (data not reported) | NA | Strong sensory | 100 Hz pulses every 2.4 Hz, 10 ms PW | Concha (right) | 24, Healthy |
| Laqua (2014)<br>Pain | No significant effects of aVNS on HR (data not reported) | NA | Strong sensory | Alternating between 2 Hz and 10 Hz, 200 μs PW | Concha (anode) and mastoid (cathode) (bilateral) | 21, Healthy |
| Napadow (2012)<br>Pain | No significant effects of aVNS on HR (data not reported) | NA | Strong sensory | 30 Hz, 450 μs PW | Cymba concha and antihelix/cavum concha slope (left) | 15, Chronic pelvic pain patients |
| HRV Measurements |  |  |  |  |  |  |
| LF/HF | Ratio of low frequency over high frequency cardiac activity (frequency-domain measurement) |  |  |  |  |  |
| RMSSD | Root mean square of successive differences between heartbeats (time-domain measurement) |  |  |  |  |  |
| pNN50 | The proportion of NN50 divided by the total number of NN (R-R) intervals. NN50 is the number of times successive heartbeat intervals exceed 50ms (time-domain measurement) |  |  |  |  |  |
| SDNN | Standard deviation of the NN (R-R) intervals (time-domain measurement) |  |  |  |  |  |
| *Risk of Bias score indicated by box color: red for ‘high’, orange for ‘some concerns’, and green for ‘low’ risk of bias. |  |  |  |  |  |  |
| **Refer to text above table 1 for definitions of all active stimulation levels (*First sensory titration*: Subject is barely able to feel a cutaneous sensation. *Strong sensory titration*: Subject feels a strong, but not painful or uncomfortable sensation from the stimulation. *Pain titration*: Stimulation amplitude is increased until the subject feels a painful or uncomfortable sensation) |  |  |  |  |  |  |
| A list of abbreviations is found in supplementary material. |  |  |  |  |  |  |

**Table 7:**
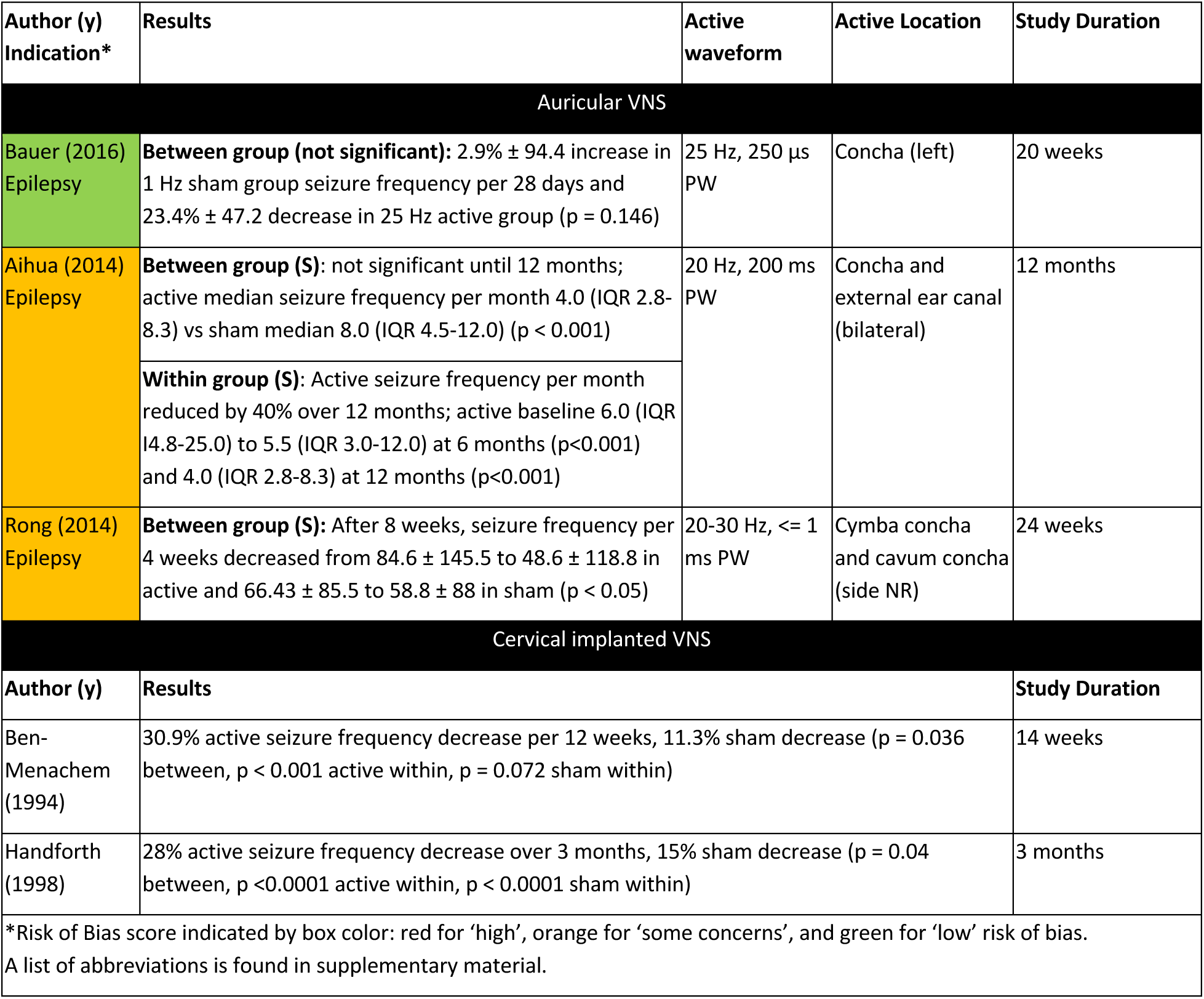
Summary of epilepsy studies

| Author (y)<br>Indication* | Results | Active<br>waveform | Active Location | Study Duration |
| --- | --- | --- | --- | --- |
| Auricular VNS |  |  |  |  |
| Bauer (2016)<br>Epilepsy | **Between group (not significant):** 2.9% ± 94.4 increase in 1 Hz sham group seizure frequency per 28 days and 23.4% ± 47.2 decrease in 25 Hz active group (p = 0.146) | 25 Hz, 250 μs<br>PW | Concha (left) | 20 weeks |
| Aihua (2014)<br>Epilepsy | **Between group (S):** not significant until 12 months; active median seizure frequency per month 4.0 (IQR 2.8-8.3) vs sham median 8.0 (IQR 4.5-12.0) (p < 0.001)<br>**Within group (S):** Active seizure frequency per month reduced by 40% over 12 months; active baseline 6.0 (IQR 4.8-25.0) to 5.5 (IQR 3.0-12.0) at 6 months (p<0.001) and 4.0 (IQR 2.8-8.3) at 12 months (p<0.001) | 20 Hz, 200 ms<br>PW | Concha and external ear canal (bilateral) | 12 months |
| Rong (2014)<br>Epilepsy | **Between group (S):** After 8 weeks, seizure frequency per 4 weeks decreased from 84.6 ± 145.5 to 48.6 ± 118.8 in active and 66.43 ± 85.5 to 58.8 ± 88 in sham (p < 0.05) | 20-30 Hz, ≤ 1 ms PW | Cymba concha and cavum concha (side NR) | 24 weeks |
| Cervical implanted VNS |  |  |  |  |
| Author (y) | Results | Study Duration |  |  |
| Ben-Menachem (1994) | 30.9% active seizure frequency decrease per 12 weeks, 11.3% sham decrease (p = 0.036 between, p < 0.001 active within, p = 0.072 sham within) | 14 weeks |  |  |
| Handforth (1998) | 28% active seizure frequency decrease over 3 months, 15% sham decrease (p = 0.04 between, p < 0.0001 active within, p < 0.0001 sham within) | 3 months |  |  |
| *Risk of Bias score indicated by box color: red for ‘high’, orange for ‘some concerns’, and green for ‘low’ risk of bias.<br>A list of abbreviations is found in supplementary material. |  |  |  |  |

**Table 8:** Summary of pain studies

| Author (y)<br>Indication* | Results (compared to control) | Duration of<br>study | Subjects analyzed,<br>disease |
| --- | --- | --- | --- |
| Busch (2013) Pain | **Not significant:** fourteen parameters were measured on both ipsilateral and contralateral. The pilot study did not correct for multiple comparisons. Mechanical and tonic heat pain parameters were marked for statistical significance and planned to be investigated further. | 2 sessions (48 hrs apart) | 48, Healthy |
| Janner (2018) Pain | **Not significant:** perceived pain intensity and temporal summation of pain not significant between active, placebo, and sham. | 4 sessions (48 hrs apart) | 49, Healthy |
| Juel (2017) Pain & Gastrointestinal Motility | **Not significant:** no significance in evoked pain threshold between active and sham. | 2 sessions (1 week apart) | 20, chronic pancreatitis patients |
| Kovacic (2017) GI pain | **Significant:** sham had significantly higher intensity pain at end of 3 weeks (median 7.0, IQR 5.0–9.0) compared to active (5.0, 4.0–7.0; $p = 0.003$). The composite score of the Pain-Frequency-Severity-Duration (PFSD) was significantly lower in the active group (8.4, IQR 3.2–16.2) than those in the sham group (15.2, 4.4–36.8) $p = 0.003$. | 4 weeks | 104, children with GI pain |
| Frøkjær (2016) Gastroduodenal motility and pain threshold | **Not significant:** compared to sham, evoked pain threshold on bone increased ($p = 0.001$). Muscle evoked pain thresholds were significantly different at baseline ($p = 0.013$). | 2 sessions (~6 days apart) | 18, Healthy |
| Johnson (1991) Pain threshold and autonomic function | **Not significant:** no significance in evoked pain threshold or autonomic measures between any of the 3 active groups and control. | 1 session | 24, Healthy |
| Kutlu (2020)<br>Fibromyalgia | **Not significant:** little improvement in pain visual analog scale (VAS) ($p = 0.084$), physical role difficulty ($p = 0.496$), or emotional role difficulty ($p = 0.194$) between control (exercise) and active group (aVNS and exercise). | 4 weeks | 52, Fibromyalgia patients |
| Laqua (2014) Pain | **Not significant:** no significant evoked pain threshold difference between aVNS and placebo. | 2 sessions (1 week apart) | 21, Healthy |
| Napadow (2012) Pain | **Significant:** reduction in deep pain intensity rating ($p = 0.049$). Reduction in temporal summation ($p = 0.04$). | 2 sessions (1 week apart) | 15, Chronic Pelvic Pain due to endometriosis patients |
| Straube (2015) Chronic Migraine | **Significant:** headache occurrence -7.0 per 28 days in 1 Hz group (36.4% reduction) and -3.3 in 25 Hz group (17.4% reduction) ($p = 0.035$). | 12 weeks | 46, Chronic migraine patients |
| *Risk of Bias score indicated by box color: red for 'high', orange for 'some concerns', and green for 'low' risk of bias. A list of abbreviations is found in supplementary material. |  |  |  |

**Table 6:** Summary of inflammatory studies

| Author (y)<br>Indication* | aVNS group cytokine level results (compared to control) | Measurement method | Subjects, disease |
| --- | --- | --- | --- |
| Addorisio (2019)<br>Rheumatoid Arthritis | TNF-α decrease (p < 0.05) | Endotoxin-induced | 19, Healthy |
|  | IL-6 decrease (p < 0.001) |  |  |
| | IL-1β decrease (p < 0.01) | | |
| Andreas (2019)<br>postop AF | IL-6 not significant | Endotoxin-induced | 40, patients undergoing cardiac surgery |
|  | CRP not significant |  |  |
| Stavrakis (2015) Atrial Fibrillation | TNF-α decrease (p < 0.05) | Endotoxin-induced | 40, paroxysmal AF ablation patients |
|  | IL-6 not significant |  |  |
|  | IL-10 not significant |  |  |
|  | CRP decrease (p < 0.05) |  |  |
| Stavrakis (2020) Atrial Fibrillation | TNF-α decrease (p = 0.0093) | Circulating | 53, diagnosed with Paroxysmal AF |
|  | IL-6 not significant |  |  |
| | IL-1β not significant | | |
|  | IL-17 not significant |  |  |
| Yu (2017) Myocardial Ischemia-Reperfusion Injury (MIRI) | TNF-α lower increase than control (p < 0.05) | Circulating | 95, MIRI patients |
|  | IL-6 lower increase than control (p < 0.05) |  |  |
| | IL-1β lower increase than control (p < 0.05) | | |
|  | HMGB1 lower increase than control (p < 0.05) |  |  |
| Afanasyev (2016)<br>Coronary Insufficiency and left ventricular dysfunction | Active group 1a HSP60 increase (p < 0.05); HSP70 increase p < 0.05) | Circulating | 70, coronary insufficiency/LV dysfunction patients |
|  | Active group 2 HSP60 not significant; HSP70 increase (p < 0.05) |  |  |
| Salama (2020) Acute inflammatory response after lung lobectomy | TNF-α not significant | Circulating | 100, lobectomy via thoracotomy in patients with non-small cell lung cancer |
|  | IL-6 lower increase than control (p = 0.02) |  |  |
|  | IL-10 not significant |  |  |
| | IL-1β not significant | | |
|  | IL-18 not significant |  |  |
|  | CRP lower increase than control (p = 0.01) |  |  |
| *Risk of Bias score indicated by box color: red for ‘high’, orange for ‘some concerns’, and green for ‘low’ risk of bias.<br>A list of abbreviations is found in supplementary material. |  |  |  |

Given that cardiac effects are closely related to a subject’s comfort and stress levels, it is important to consider trial design influences such as subject familiarization. For example, a clinical trial visit could increase stress levels and blood pressure of the subjects (Wright et al., 2015) and mask any potential therapeutic effects of aVNS on blood pressure. Another example of subject familiarization is related to HRV. Borges et al. (2019) tried to accommodate for subject familiarization by delivering a five minute ‘familiarization’ stimulation at the cymba concha before the experimental intervention. Unfortunately, baseline HRV measurements were taken immediately after these ‘familiarization’ stimulation sessions and could be affected by the stimulation delivered. Hence, there is no true baseline measurement – before any stimulation is applied – of HRV and casts doubt on the findings of the study, which claimed no significant effects of aVNS on HRV. In summary, conflicting results on the cardiac effects of aVNS could be attributed to trial design and measurement methods.

#### Inflammatory related effects of aVNS

Several studies measured cytokine levels to investigate the anti-inflammatory effects of aVNS. Cytokine levels were either measured directly in drawn blood (circulating) or after an *in vitro* endotoxin-induced challenge on drawn blood. In the four studies that measured circulating cytokine levels, results are somewhat conflicting. Stavrakis et al. (2020) reported a significant decrease in TNF-*α*and no significant changes in IL-6, IL-1*β*, IL-10, and IL-17, consistent with subjects with moderate atrial fibrillation burden and not suffering from any inflammatory condition. TNF-*α*is one of the most abundant mediators in inflamed tissue and is present in the acute inflammatory response (Parameswaran and Patial, 2010). In subjects being treated for myocardial infarction, Yu et al. (2017) reported that the active group was significantly lower than the control group for all measured cytokine levels (TNF-*α*, IL-6, IL-1*β*, and high-mobility group-box 1 protein (HMGB1)). Unlike Stavrakis et al. (2020) and Yu et al. (2017), Salama et al. (2020) reported no statistically significant change in TNF-*α*, along with lower levels of c-reactive protein (CRP) and IL-6, compared to control. CRP is also an acute phase protein whose release from the liver is stimulated by increased levels of IL-6 (Giudice and Gangestad, 2018). Lastly, Afanasiev et al. (2016) measured HSP60 and HSP70, which are heat shock proteins and responsible for preventing damage to proteins in response to stressors such as high temperature (Morimoto, 1993). Both HSP60 and HSP70 increased significantly in Afanasiev et al. (2016), indicating a potential anti-inflammatory effect.

The limited applicability of *in vitro* endotoxin-induced assays was discussed in Stoddard et al. (2010), Yang et al. (2011), and Thurm et al. (2005). Additionally, Broekman et al. (2015) provided an example where an *in vitro* assay was unsuccessful in identifying disease severity in patients with a quiescent autoimmune disorder. Nonetheless, we summarize the findings on anti-inflammatory effects of aVNS on *in vitro* endotoxin-induced assays. In Stavrakis et al. (2015), acute stimulation was delivered intraoperatively to subjects undergoing ablation treatment for atrial fibrillation. After 1h of stimulation, there was a significant decrease in TNF-*α*and CRP levels in femoral vein draws. In Addorisio et al. (2019), vibrotactile stimulation was applied for only 2 minutes and showed a statistically significant decrease in endotoxin-induced cytokine levels of TNF-*α*, IL-6, and IL-1*β*in blood drawn one hour after stimulation.

Overall, these studies provide evidence that aVNS may reduce circulating levels and endotoxin-induced levels of inflammatory markers and suggest a potential anti-inflammatory effect of aVNS. The clinical relevance of endotoxin-induced measures needs to be further explored and the implications of lowered circulating cytokine levels on disease burden needs to be further investigated in RCTs.

#### Epilepsy related effects of aVNS

The three studies that investigated the antiepileptic effects of aVNS were all chronic studies. All used stimulation frequencies between 20-30 Hz similar to implantable VNS (LivaNova, 2017). However, other stimulation parameters varied widely. All studies reported a 20-40% decrease in seizure frequency from baseline and showed significance from baseline after a few weeks to months of daily prescribed stimulation.

Based on these three chronic studies, there is some evidence to support the anti-epileptic effects of aVNS. The primary outcomes of these non-invasive interventions are comparable to that of implantable VNS in studies of similar duration and sample size (Ben-Menachem et al., 1994; Handforth et al., 1998). However, due to concerns over unblinding and weaker evidence in between group analysis compared to within group analysis, it is possible that the effects may be attributed to placebo. The concern that this effect is placebo is exacerbated by the fact that the studies used widely varying stimulation parameters yet achieved similar results.

#### Pain related effects of aVNS

Ten studies investigated the effects of aVNS on the amelioration of pain. Five studies investigated the effects of aVNS on evoked pain threshold levels in healthy subjects but used varying pain-assessment methods. One study investigated the effects of aVNS on evoked pain threshold levels in chronic pancreatitis patients (Juel et al., 2017). The other four studies examined the effects of aVNS on self-reported pain scores in patients already suffering from pain due to endometriosis (Napadow et al., 2012), chronic migraine (Straube et al., 2015), fibromyalgia (Kutlu et al., 2020), and gastrointestinal (GI) disorders (Kovacic et al., 2017).

Across studies, the observed effects of aVNS for pain are highly varied. In studies that investigated evoked pain thresholds, results showed negligible changes in pain threshold levels due to aVNS therapy. For chronic migraines, Straube et al. (2015) reported a therapeutic effect in both the 1 Hz and 25 Hz stimulation groups. Unexpectedly, the 1 Hz stimulation, originally designated as sham in the trial design, resulted in a reduction of headaches – comparable to medications used in migraine prevention – while the 25 Hz treatment had a smaller effect on the reduction of headaches (−7.0 episodes with 1 Hz vs −3.3 episodes with 25 Hz over 28 days). Studies to isolate stimulation parameters that are therapeutic for pain are needed given that the 1 Hz stimulation, generally considered a sham due to its low frequency, was more effective than the 25 Hz intervention group. For gastrointestinal-related pain and chronic pelvic pain, pain was also significantly ameliorated by aVNS therapy. Overall, these studies provide evidence that aVNS may be therapeutic for some pain conditions, but more studies are needed to further explore the effectiveness for specific medical conditions and rule out significant contributions from placebo effects.

#### Other effects of aVNS

Other potential effects of aVNS investigated clinically included severity of motor symptoms in Parkinson’s disease, depression, schizophrenia, obesity, impaired glucose tolerance, gastroduodenal motility, and tinnitus. Most of these effects were only investigated in a single RCT and there is not sufficient evidence to synthesize and evaluate across trials. The results of these individual studies are summarized in supplementary material. More studies are needed to further explore the effects of aVNS for these indications.

## 4. DISCUSSION

This is the first systematic review applying the Cochrane Risk of Bias (RoB) framework to auricular vagus nerve stimulation (aVNS) clinical trials. Our systematic review of 38 publications, totaling 41 RCTs, shows high heterogeneity in trial design and outcomes - even for the same indication. In the extreme, outcomes for heart rate effects of aVNS ranged from a consistent decrease in every subject in two studies to no heart rate effects in other studies. Findings on heart rate variability (HRV) conflict between studies and were hindered by trial designs including inappropriate washout periods and multiple methods used to quantify HRV. Early-stage evidence suggests aVNS reduces circulating levels and endotoxin-induced levels of inflammatory markers. Studies on epilepsy reached primary endpoints similar to previous RCTs on implantable VNS, albeit with concerns over quality of blinding. Clinical studies that tested aVNS for pain showed preliminary evidence of ameliorating pathological pain but not induced pain. Across the board, there are concerns on the extent of the contributions by placebo effects – especially since novel medical devices, such as aVNS devices, which also produce abnormal sensations (i.e., paresthesia), have a known larger placebo effect (Doherty and Dieppe, 2009).

The highest level of clinical evidence is multiple homogenous high quality RCTs as outlined in the Oxford CEBM Levels of Clinical Evidence Scale. The outcomes of these trials must consistently support the efficacy and safety of the therapy for a specific clinical indication. In the reviewed trials, several root causes - design of control, unblinding, and inconsistent reporting of results - raise the level of concern for bias in the outcomes. The current quality of evidence for aVNS RCTs supporting a particular clinical indication may generally be placed at grade 2, for ‘low quality’ RCT, on the Oxford CEBM scale (Centre for Evidence-Based Medicine, 2009). An RCT is considered ‘low quality’ for reasons including imprecise estimates, variability in results, indirect evidence, and presence of publication bias. For aVNS to reach the highest quality of clinical evidence for a particular indication, multiple RCTs must homogeneously support the safety and efficacy of the therapy for that indication.

In the following sections, we discuss gaps and improvements informed by our systematic review to aid the development of aVNS therapies. In the short term, we suggest improvements in the reporting of clinical trial results to allow meta-analysis of results across aVNS studies. In the long term, we highlight the need for direct measures of target engagement as biomarkers to study therapeutic effects and therapy limiting side effects, and better translate learnings from animal models to humans. Also in the long term, we discuss the needs and associated challenges in careful design of controls and maintenance of blinding.

### 4.1 Short Term Solutions – Guide to Reporting

Several steps can be implemented immediately to increase the quality and consistency of reporting in aVNS studies, enabling comparison of results across studies.

Across the studies, the greatest risk of bias came from the Cochrane section ‘selection of reported results.’ A comprehensive guide to clinical trial reporting is found published by the CONSORT group along with detailed elaborations (Moher et al., 2010). See Kovacic et al. (2017) for an aVNS study that followed the CONSORT reporting recommendations. Pre-registration of trials, use of appropriate statistical analysis, justifying clinical relevance of outcome measures, and contextualizing clinical significance of results are discussed here. If followed across aVNS studies, these suggestions would reduce the risk of bias identified in the RoB section reporting of results and enable the synthesis of knowledge by making reporting more comparable across studies (Farmer et al., 2020).

#### Pre-registration

Pre-registration of planned enrollment, interventions, outcome measures and time points, and statistical plan to reach primary and secondary endpoints reduces risk of bias in the reporting of results. When the trial is reported, commentary should be made on adherence and deviations from the pre-registration with appropriate justifications. Exploratory analysis of the data may still be performed but needs to be denoted. Exploratory analysis can be used to suggest design of future investigations. The amount of exploratory analysis should be limited, and all non-significant exploratory analysis performed before reaching the significant results should also be reported. Of the 41 aVNS RCTs reviewed, 13 RCTs pre-registered, but only 5 had sufficient information to be considered a complete pre-registration. Pre-registration reduces risk of bias in reporting of results by preventing analysis of only select measures (section 5.1 of RoB rubric) and multiple analysis of data (section 5.2 of RoB rubric).

#### Appropriate statistical analysis

Data may be analyzed in many ways to claim the effect of an intervention. For example, studies may report a between group analysis comparing the change in the active arm to the change in the control arm or a within group analysis comparing the active arm after treatment to baseline. The more appropriate method for a controlled study is a between group comparison of the active arm versus the control arm. Several studies claimed statistically significant findings based just on the within group analysis even if the between group analysis was non-significant. An example illustrating this difference is found in supplementary material. Pre-registration of the planned statistical analysis will discourage unjustified multiple analysis of the data.

In crossover design studies, there was a major gap in the reporting of baseline comparison between randomized groups. Even in a crossover design where each subject receives all interventions, it is crucial to compare baseline differences between groups as one would do for a parallel study. This is especially pertinent in pilot studies with small sample sizes, where a baseline imbalance between groups is more likely to occur and affect the trial outcome (Kang et al., 2008). Additionally, if the order of intervention becomes pertinent, due to an incomplete washout period or compromised blinding, then it is essential that the baseline randomization between groups is balanced to enable further analysis.

Another concerning gap in crossover design studies was in the lack of reporting the statistical test for carryover effects. The test detects if the order of intervention received had an effect on the outcome (Shen and Lu, 2006). The test for carryover effects shows significance when there are incomplete washout effects, baseline imbalances, or compromise in blinding. It is perhaps the single most important gauge of the quality of a crossover design and should always be performed and reported – only 4 of 20 crossover design studies reported the carryover effects test. A baseline comparison between groups will ensure that baseline differences do not contribute to significance in the test for crossover effects – allowing effects from incomplete washout periods and compromised blinding to be isolated.

Reporting of individual results is a simple and effective way to convey the average and variance in outcomes, the fraction of responders, and worsening of symptoms (if any) in the non-responders. In Stavrakis et al. (2020) there is worsening of symptoms in the non-responders (53% of the active group) at the three-month evaluation, which is also the only time point at which atrial fibrillation burden is measured concurrently during stimulation. This clinically relevant finding was evident during review because individual results were presented. Individual results were only presented in 10 of 41 aVNS studies reviewed. Reporting of individual results should be considered where allowed by clinical trial protocol.

#### Justify clinical relevance of outcome measures

The outcome measure itself may not be established as clinically relevant. For example, *in vitro* endotoxin-induced cytokine measurements were used to proxy *in vivo* immune response in several aVNS studies including Addorisio et al. (2019). While endotoxin-induced cytokine levels produce a stronger signal, they may not be clinically relevant in the case of an auto-immune disease such as Rheumatoid Arthritis tested in Addorisio et al. (2019).

The clinical accuracy of the measurement tool must also be considered. For example, aVNS studies often used photoplethysmography (PPG) based methods at the finger to measure blood pressure. Given the change in blood pressure signal during aVNS is already small, it is unnecessary to lose statistical power by using less accurate PPG based methods (Elgendi et al., 2019) to measure blood pressure. Clancy et al. (2014) used both a finger-based PPG, Finometer®, and a traditional arm sphygmomanometer to measure blood pressure and concluded that the increase in blood pressure measured using the Finometer® may be due to an artifact of the PPG measurement method. Discussion on clinical relevance of the outcome measure provides justification for the selection of reported results.

#### Contextualize clinical significance of results

An outcome that is statistically significant does not necessarily indicate clinical significance. Contextualizing the trial results allows the reader to better understand the clinical significance of the findings. This may be done by comparing the study outcome to the outcome of the standard of care or another therapy. For example, Cakmak et al. (2017), in an aVNS trial for Parkinson’s disease, showed a 5.3 points improvement on the UPDRS part 3 for motor symptoms (Goetz et al., 2008). Their result could be contextualized with the 18.4 points improvement in DBS (Kahn, 2019).

Clinical significance of results should be discussed in relevance to the subject population – particularly with consideration to disease severity and heterogeneity. For example, Juel et al. (2017) repeated a study in the diseased population after Frøkjaer et al. (2016) first reported a similar trial in healthy subjects. While the study in healthy subjects concluded significant findings, the subsequent study in diseased subjects did not. They cited pathological neural circuitry as a possible reason. Whether the difference was due to pathophysiology or differences in trial design and analysis is uncertain. Regardless, inclusion and exclusion criteria often restrict the subjects enrolled in terms of disease severity and heterogeneity and consideration should be given to the study subject population when discussing clinical significance of the findings.

Risk of bias identified in the selection of reported results can be addressed by pre-registration of trials, use of appropriate statistical analysis, justifying clinical relevance of outcome measures, and contextualizing clinical significance of results. These ideas are summarized in Table 9.

**Table 9:** Checklist for trial reporting

|  | Item |
| --- | --- |
| ✓ | Pre-register |
| ✓ | Use appropriate statistical analysis |
| ✓ | Justify choice of between versus within group analysis |
| ✓ | Perform baseline comparison (even in crossover studies) |
| ✓ | Perform statistical test for carryover effects |
| ✓ | Report individual results |
| ✓ | Justify clinical relevance of outcome measures |
| ✓ | Discuss clinical significance of results |
| ✓ | Compare outcome to existing therapy |
| ✓ | Consider subject population when generalizing findings |

### 4.2 Lessons from drug world

As the translation of drugs into clinical use is more established than neuromodulation therapies, it is instructive to review the translation of drugs for pitfalls in moving towards FDA market approved therapies. Less than 12% of drugs that received an FDA Investigational New Drug approval to begin human studies (the current stage of development of many aVNS based therapies) reached market approval (DiMasia et al., 2016; Paul et al., 2010). Gupta et al. (2011) identified several factors that hindered the successful translation of drug therapies from early-stage results to market approval, which are also relevant to aVNS therapies.

1. Lack of pharmacodynamic measures in early-stage clinical trials to confirm drug activity (Gallo, 2010). This is similar to the lack of evidence that aVNS is activating desired fiber types in the auricular branch of the vagus and not activating other fiber types including those within the great auricular, lesser occipital, facial, and trigeminal nerves which innervate the auricular and periauricular region.
2. Lack of validated biomarkers for on- and off-target engagement – impacting our ability to assess and confirm therapeutic activity versus side effects (Institute of Medicine, 2014). Again, similar to the lack of biomarkers in aVNS trials to confirm on- and off-target nerve activation.
3. Lack of predictability of animal models for humans (Johnson et al., 2001). Relevant in aVNS to translatability of electrode configuration, dosing, and stimulation parameters given changes in size, neuroanatomy, and neurophysiology from animal models to humans. A concern confirmed by other neuromodulation therapies (De Ferrari et al., 2017).

A method to directly measure local neural target engagement will provide an immediate biomarker of on- and off-target activity and forestall some of the hurdles encountered in drug therapy development. A minimally invasive method to measure target engagement percutaneously could be deployed across preclinical models and early clinical studies. Doing so would increase the translatability of findings by providing data to titrate electrode design, placement, and stimulation waveform parameters to optimize for on-target engagement.

### 4.3 Long term solutions – target engagement

On- and off-target nerve activation is especially relevant in the case of the auricle that is innervated by several nerves with uncertainty on the specific areas of innervation in literature and across subjects. Data from direct measures of on- and off-target engagement could be used 1) to titrate the therapy by adjusting electrode design, placement, and stimulation waveform parameters to optimize on-target engagement, to scale pre-clinical animal doses to humans by preserving the fiber types activated, and 3) to help investigate fundamental mechanisms of action by isolating local neural pathways.

aVNS is commonly delivered at the cymba concha with the assumption that the cymba concha is innervated only by the auricular vagus. This is based on two pieces of evidence. Firstly, Peuker and Filler (2002) showed the cymba concha is innervated only by the auricular vagus in 7 of 7 cadavers. Notwithstanding, there may be variation in innervation or spread of the electric field, which could activate the neighboring auriculotemporal branch of the trigeminal nerve and even the great auricular nerve. Variations in peripheral nerve innervation has been well studied for other regions of the body, such as the hand (Guru et al., 2015; Bas and Kleinert, 1999). The reliance on the Peuker and Filler study is concerning due to its small sample size. Given the importance of the claims in Peuker and Filler, a further dissection study with a larger sample size is called for to investigate whether the results hold over a larger population of ethnically diverse individuals. Secondly, functional magnetic resonance imaging (fMRI) evidence is also used to suggest vagal innervation of the conchae (Frangos et al., 2015). However, fMRI is a surrogate measure of target engagement and is especially problematic when imaging deep in the brainstem, as described below.

Target engagement is commonly established using secondary surrogates such as fMRI, somatosensory evoked potentials (SSEPs), and cardiac measures. Secondary surrogates of target engagement are often contaminated with physical and biological noise, leading to potential confounds. For example, Botvinik-Nezer et al. (2020) and Becq et al. (2020) showed that results of fMRI studies were highly dependent on data processing techniques applied. In addition, pathways starting from the trigeminal nerve in the auricle also connect to NTS (Chiluwal et al., 2017), and activate the same region in the brain when in fact the auricular vagus might not be recruited during stimulation. fMRI of the brainstem is further complicated (Napadow et al., 2019) as distance from the measurement coils increases, the effective resolution decreases (Gruber et al., 2018). Still further, novelty, such as being stimulated in the ear or being in an MRI scanner, activates the locus coeruleus (LC) (Wagatsuma, 2017), which has connections with NTS. Thereby confounding potential aVNS effects on NTS with LC induced activity due to novelty effects. Lastly, fMRI has nonstandard results between subjects requiring individual calibration and making subject to subject comparisons challenging. Additionally, SSEP recordings are sometimes contaminated and misinterpreted due to EMG leakage (Usami et al., 2013). The common measures of target engagement are secondary surrogates and prone to confounds – creating a need for direct measures of local target engagement at the nerve trunks innervating the auricle.

Given the lack of direct measures of local target engagement, aVNS studies rely largely on stimulation parameters that are similar to those used for implantable VNS. The assumption that these stimulation parameters will result in similar target engagement and therapeutic effects may not hold due to the differences in target fiber type, fiber orientation, and electrode design and contact area – all of which affect neural recruitment. Cardiac effects of implantable VNS are thought to be mediated by activation of parasympathetic efferent B fibers innervating the heart (Sabbah et al., 2011) or aortic baroreceptor afferents depending on the stimulation parameters used. Studies of the baroreceptors at the aorta and carotid sinus bulb identified fiber types consistent with A*δ* and C fibers (Reynolds et al., 2006; Seagard et al., 1990). Strikingly, consensus workshops have suggested that aVNS is mediated by activation of A*β* fibers (Kaniusas, 2019). There is also no evidence of baroreceptors identified in the ear. These difference between the auricular and cervical vagus suggests a direct porting of stimulation parameters developed for implantable VNS would be insufficient to invoke cardiac responses, unless another yet unidentified mechanism for cardiac responses mediated through NTS is responsible, which can be targeted by A*β* fiber input that then indirectly modulates sympathetic or parasympathetic input to the heart. Unlike stimulation of the cervical vagus nerve trunk, where the electrode contacts are oriented parallel to the target axons, electrode contacts for aVNS do not have consistent orientation with respect to the target axons, which exist as a web of axons in the auricle. Orientation of fibers relative to the stimulation electrode have a large effect on fiber recruitment (Grill, 1999) and could potentially lead to preferential activation of nerve pathways oriented in a particular direction to the stimulation contacts, as well as inconsistent activation of specific fiber types across the auricle. Additionally, target fibers in the auricle transition to unmyelinated fibers as they approach sensory receptor cells (Provitera et al., 2007). For these reasons, while cathodic leading stimulation might have lower recruitment thresholds in implantable VNS, the principle may not hold for aVNS (Anderson et al., 2019). In aVNS, target fiber type, electrode design, electrode size, transcutaneous placement, and orientation of the target fiber relative to the electrode are different both compared to implantable VNS and across aVNS studies. Therefore, it is unsurprising that stimulation parameters ported from implantable VNS may not replicate the physiological effects or recruitment of fiber types that have been observed during implantable VNS.

In relation to electrode design, injected charge density, as opposed to current or voltage, is the most relevant metric of neural activation. This is because stimulation evoked action potentials occur in regions of the neural cell membrane where there is an elevated charge density (Rattay, 1999; McNeal, 1976). For effective comparison across studies using different electrodes, it is imperative to report on the electrode area, especially on the area as it makes contact with tissue, along with stimulation current.

The above discussion stresses a general lack of confidence in ascertaining which of several nerve trunks innervating the auricle are being activated during aVNS that may be generating the on- or off-target effects. Cakmak et al. (2017) further proposed that the therapeutic effects they reported for motor symptoms of Parkinson’s diseases came from direct recruitment of the intrinsic auricular muscles instead of the auricular vagus nerve. The fundamentals of neural stimulation do not support directly porting stimulation parameters from implantable VNS. Therefore, it is important to understand which fiber types on the auricular vagus are being activated, if at all. This knowledge requires direct measures of local target engagement from the nerves innervating the auricle.

To further the development of aVNS, it will be essential to understand local target engagement of the nerve trunks innervating the ear. Ultrasound guided (Ritchie et al., 2016) percutaneous microelectrode recordings (Ottaviani et al., 2020) from the major nerve trunks innervating the ear, similar to the technique to measure muscle sympathetic nerve activity (MSNA), provides a way to directly measure local neural recruitment. Real-time data on neural target engagement would enable optimization of stimulation parameters, electrode, and control designs. These data would also improve translation of stimulation dosages from animal models to humans. Tsaava et al. (2020) showed that the anti-inflammatory effects of implantable VNS are stimulation dose dependent and could lead to the worsening of inflammation at certain dosages. Since target engagement in humans is currently unknown, there is no consistent means of determining dosage accurately – raising potential safety concerns. This minimally invasive method to record neural target engagement is already used clinically and could be rapidly translated to the clinic for use in titrating neuromodulation therapies.

Understanding primary target engagement at the ear will also further our understanding of aVNS mechanisms. For example, large animal recordings of evoked compound action potentials from the major nerve trunks innervating the ear may help in understanding the relationship between on- and off-target nerve engagement and corresponding physiological effects. Simultaneous recordings at the cervical vagus may allow differentiation of direct efferent vagal effects versus NTS mediated effects, which would appear with a longer latency due to synaptic delay and longer conduction path length. Measuring neural target engagement at the auricle provides a first step to systematically studying aVNS mechanisms and optimizing clinical effects.

### 4.4 Long term solutions – improvement in control design and blinding

The design of an indistinguishable yet nontherapeutic control is central to maintaining the blinding in a clinical trial. Stemming from limited understanding of local target engagement and mechanism of action of aVNS, it is difficult to implement an active control (i.e., sham) that has similar perception to the therapeutic group but will not unknowingly engage a therapeutic pathway. This uncertainty in the therapeutic inertness of the control violates some of the basic premises for a RCT and makes it difficult to evaluate aVNS RCTs on the Oxford Scale for clinical evidence. Systematic effort must be made to design controls, which are key to maintaining blinding in aVNS RCTs.

Common control designs used in aVNS studies are summarized in Fig. 5. A placebo is defined when the electrode placement and device are similar to the active intervention, but no stimulation is delivered. A waveform sham is defined when a different – nontherapeutic – waveform is delivered at the same location as active intervention. In a location sham, the same waveform as active intervention is delivered at a different location on the auricle and should not engage a therapeutic nerve. Lastly, no intervention or a pharmacological control may be used. Location sham was the most common control used in 16 of 41 RCTs reviewed. These different control designs are evaluated at length in supplementary material along with recommendations on appropriate control types depending on trial design.

**Figure 5:**
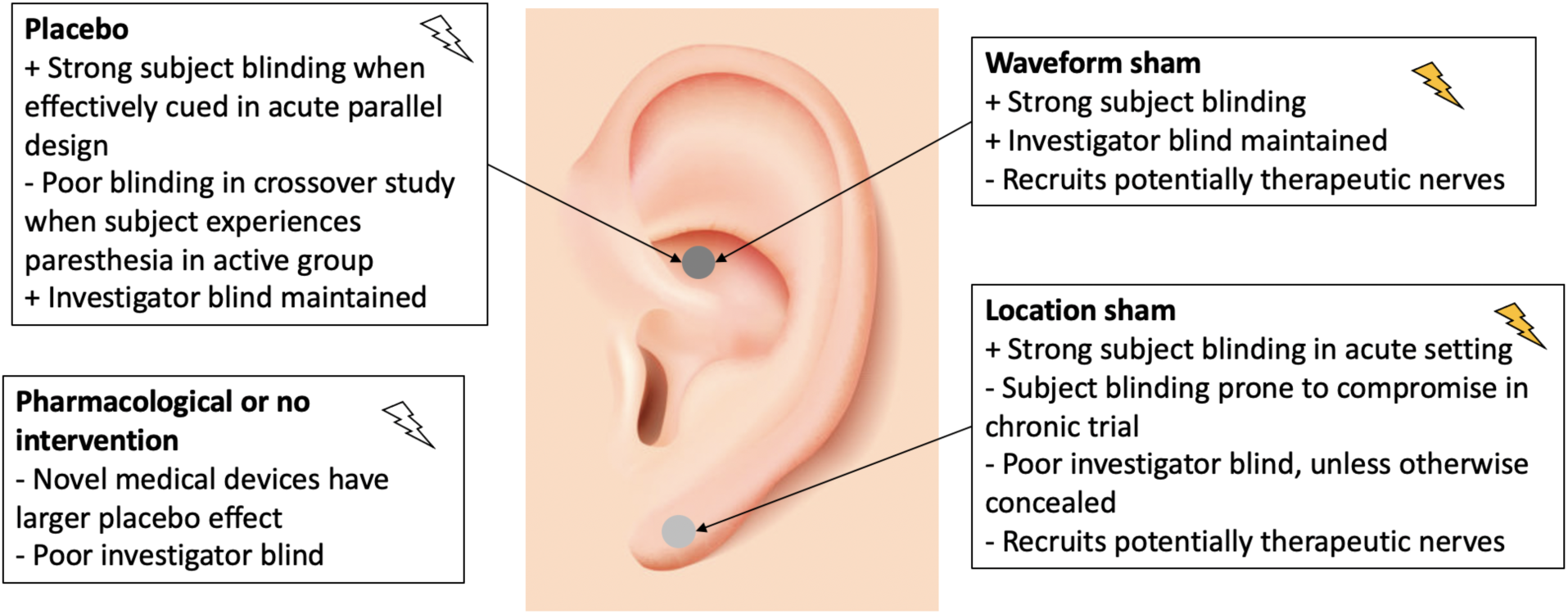
Types of controls used in aVNS clinical trials. The ideal control is indistinguishable from active intervention, to both subject and investigator, yet therapeutically inert.

Inappropriate implementation of the control group resulted in compromised blinding in many studies. Subject unblinding occurred when subjects were able to feel a paresthesia in the active intervention but not in the control. For example, in cross-over trials, where subjects undergo both the active and control intervention, post-hoc assessment of blinding becomes critical given the perception of sensation being unequal could easily break the blind. Investigator unblinding occurred when investigators were able to see differences in electrode placement or device operation. Unblinding due to inappropriate control design is the main contributor leading to risk of bias in the Cochrane section ‘deviation from intended intervention’. The design of appropriate controls is difficult for trials testing non-pharmacological interventions – especially for paresthesia-inducing neuromodulation trials (Robbins and Lipton, 2017; Nature Biotechnology, 2019) – but is essential to establish a double blind.

To aid in the maintenance of the double blind, appropriate design of controls and consistent evaluation of blinding are required. Measurement of target engagement via microneurography of the major nerve trunks innervating the ear will enable understanding of neural recruitment occurring during active and sham stimulation and guide appropriate design of controls. In addition, post-hoc evaluation of blinding in subjects and investigators will gather knowledge on the blind quality setup by respective control designs. Methods to assess the quality of blinding in non-invasive neuromodulation studies are discussed in supplementary material. Over time, the consistent use of control designs and evaluation of blinding will grow our understanding of the concealability and therapeutic inertness of various control methods.

### 4.5. Applicability of findings to similar neuromodulation therapies

The discussion on lack of direct measures of target engagement and unknowns surrounding implementation of perceptually similar yet therapeutically inert controls to maintain the double blind are applicable to other neuromodulation therapies – especially paresthesia inducing therapies such as implantable VNS and spinal cord stimulation (SCS).

Study of local target engagement in neuromodulation therapies such as implantable VNS and SCS will inform stimulation parameters, possible mechanisms of action, and electrode design and placement to maximize on-target nerve recruitment and minimize therapy limiting off-target effects such as muscle contractions (Nicolai et al., 2020; Yoo et al., 2013). To investigate the central mechanisms of action we first have to establish local target engagement to determine which on- and off-target nerves are recruited during stimulation and which fiber types are recruited at therapeutically relevant levels of stimulation. Measurement of local target engagement in preclinical and early clinical studies provides a bottom-up approach to systematically develop and deploy neuromodulation therapies.

Paresthesia inducing neuromodulation therapies make the design of an indistinguishable yet nontherapeutic sham challenging. At the same time, an appropriate control is essential to the maintenance of a double blind and design of an RCT. A systematic review and meta-analysis of SCS RCTs for pain showed that the quality of control used had an impact on the effect size of the outcome, (Duarte et al., 2020). They concluded that thorough consideration of control design and consequent subject and investigator blinding is essential to improve the quality of evidence on SCS therapy for pain (Duarte et al., 2019). A systematic review of dorsal root ganglion (DRG) stimulation for pain also showed serious concern for bias across all studies reviewed due to compromise in subject and investigator blinding (Deer et al., 2013). The suggestions laid forth on measuring local target engagement and consistent post-hoc evaluation of blinding in subjects and investigators will expand our knowledge of effective control design for paresthesia inducing neuromodulation therapies such as aVNS, SCS, and implantable VNS.

### 4.6 Limitations of this review

This review is based on experience in other neuromodulation clinical trials and pre-clinical studies, existing frameworks for analysis such as the Cochrane Risk of Bias and Oxford clinical scale, and literature review motivated by an interest in conducting future aVNS clinical trials. However, at the writing of this article, none of the authors have conducted an aVNS clinical trial. Secondly, this is a systematic review but not a meta-analysis. Due to insufficient reporting in trials, a meta-analysis could not be conducted. The analysis was more qualitative with the intention of summarizing the quality of evidence in the field and making recommendations to improve clinical translatability. Thirdly, this review was not pre-registered, blinded, or formally randomized. Additionally, while the RoB tool provides a consistent method to evaluate trials where the shortcoming is stated explicitly, the ability to identify confounds is often reliant on the critical reading of the reviewer. This made it possible for a secondary reviewer to find additional risk of bias in several instances, which were initially missed by the primary reviewer but included upon identification and consensus. Lastly, numerous instances of missing information in trial reporting were identified and attempts were made to reach out to the authors for that information. These attempts were not always successful. Overall, the points made in the review are robust and withstand the limitations.

## 5. Conclusion

Based on our review of 38 publications, which reported on 41 aVNS clinical RCTs, aVNS shows physiological effects but has not yet shown strong clinically significant effects consistently supported by multiple studies. This review: 1) Identifies concerns in the design of trials, particularly control and blinding, and incomplete reporting of information using the Cochrane Risk of Bias analysis. 2) Finds aVNS studies are presently exploratory in nature, which is appropriate given the early stage of research of the aVNS field. Proposes guidelines for the reporting of aVNS clinical trials, which can be implemented immediately to improve the quality of evidence. 4) Proposes progress in the field has been limited by lack of direct measures of neural target engagement at the site of stimulation. Measures of target engagement will inform therapy optimization, translation, and mechanistic understanding. 5) Proposes consistent post-hoc evaluation of subject and investigator blinding and direct measures of local neural target engagement to improve the design of controls for maintenance of blinding.

As a field, neuromodulation has not yet attained social normality or gained widespread adoption as a first-line therapy (Payne and Prudic, 2009; Li et al., 2020; Daniel et al., 2004). To that end, our responsibility as pioneers is to move the field forward and build its credibility by thoroughly reporting on appropriately designed clinical trials. Given the conflicting data across aVNS studies, it is critical to implement high standards for rigor and quality of evidence to better assess the state of aVNS for a given indication.

## Supporting information

Supplementary Material 3 - Sortable table of aVNS studies and list of abbreviations

Supplementary Material 9 - Individual RoB Rubrics

## Data Availability

Not applicable.

## Acknowledgements

Eric H. Chang for notes on endotoxin-induced cytokine assays. Carly Frieders, Arkaprabha Banerjee, Maria LaLuzerne, Megan Settell, Robbin Miranda, Carolyn Huff, and Harrell Huff for reviewing drafts of this publication. Members of the Wisconsin Institute for Translational Neuroengineering (WITNe) for feedback during group meetings.

