## Supplementary Materials for "Auricular Vagus Neuromodulation – A Systematic Review on Quality of Evidence and Clinical Effects"

Supplementary Material Table of Contents

Supplementary 1 - Full search strings

Supplementary 2 - Articles excluded after full-text review

Supplementary 3 - User sortable summary of aVNS studies and list of abbreviations (separate Excel file)

Supplementary 4 - Design of controls in non-invasive neuromodulation clinical studies

Supplementary 5 - Assessing quality of blinding in non-invasive neuromodulation clinical studies

Supplementary 6 - Within versus between group comparison example

Supplementary 7 - Time to onset and chronic effects of aVNS

Supplementary 8 - Hypothesized mechanisms of action for aVNS

Supplementary 9 - Individual RoB Rubrics (separate Microsoft Word documents)

**SUPPLEMENTARY 1 - Full search strings**

The key terms used were the same for both databases. Due to the differences in the search engines, some additions were made to the Scopus search string to exclude articles before 1991 and include full-text search of the articles in the database.

|  | Scopus | PubMed |
| --- | --- | --- |
| Strategy 1 | ( sham OR placebo OR control OR controlled ) AND ( stimulation OR acupuncture OR acupoint OR stimulates OR electroacupuncture OR stimulate OR stimulated ) AND ( vagal OR vagus OR "autonomic pathway" ) AND ( ear OR auricular OR auricle ) AND ( random OR randomized OR "clinical trial" OR "controlled-trial" OR "cross-over" OR parallel OR paralleled OR "three-arm" OR "two-arm" OR "four-arm" ) AND ( EXCLUDE ( PUBYEAR , 1990 ) OR EXCLUDE ( PUBYEAR , 1989 ) OR EXCLUDE ( PUBYEAR , 1988 ) OR EXCLUDE ( PUBYEAR , 1987 ) OR EXCLUDE ( PUBYEAR , 1986 ) OR EXCLUDE ( PUBYEAR , 1985 ) OR EXCLUDE ( PUBYEAR , 1984 ) OR EXCLUDE ( PUBYEAR , 1983 ) OR EXCLUDE ( PUBYEAR , 1982 ) OR EXCLUDE ( PUBYEAR , 1981 ) OR EXCLUDE ( PUBYEAR , 1979 ) OR EXCLUDE ( PUBYEAR , 1978 ) OR EXCLUDE ( PUBYEAR , 1976 ) OR EXCLUDE ( PUBYEAR , 1975 ) OR EXCLUDE ( PUBYEAR , 1974 ) OR EXCLUDE ( PUBYEAR , 1972 ) OR EXCLUDE ( PUBYEAR , 1970 ) ) AND ( LIMIT-TO ( DOCTYPE , "ar" ) ) AND ( LIMIT-TO ( LANGUAGE , "English" ) ) AND ( LIMIT-TO ( SRCTYPE , "j" ) OR LIMIT-TO ( SRCTYPE , "p" ) ) | ( sham OR placebo OR control OR controlled ) AND ( stimulation OR acupuncture OR acupoint OR stimulates OR electroacupuncture OR stimulate OR stimulated ) AND ( vagal OR vagus OR "autonomic pathway" ) AND ( ear OR auricular OR auricle ) AND ( random OR randomized OR "clinical trial" OR "controlled-trial" OR "cross-over" OR parallel OR paralleled OR "three-arm" OR "two-arm" OR "four-arm" ) |
| Strategy 2 | ( sham OR placebo OR control OR controlled ) AND ( cerbomed OR nemos OR "Parasym health" OR "Parasym device" ) AND ( vagal OR vagus OR "autonomic pathway" ) AND ( ear OR auricular OR auricle ) AND ( random OR randomized OR "clinical trial" OR "controlled-trial" OR "cross-over" OR parallel OR paralleled OR "three-arm" OR "two-arm" OR "four-arm" ) AND ( LIMIT-TO ( DOCTYPE , "ar" ) OR LIMIT-TO ( DOCTYPE , "cp" ) ) AND ( LIMIT-TO ( LANGUAGE , "English" ) ) AND ( LIMIT-TO ( SRCTYPE , "j" ) OR LIMIT-TO ( SRCTYPE , "p" ) ) | ( sham OR placebo OR control OR controlled ) AND (CerboMed OR Nemos OR "Parasym health" OR "Parasym device" ) AND ( vagal OR vagus OR "autonomic pathway" ) AND ( ear OR auricular OR auricle ) AND ( random OR randomized OR "clinical trial" OR "controlled-trial" OR "cross-over" OR parallel OR paralleled OR "three-arm" OR "two-arm" OR "four-arm" ) |

**SUPPLEMENTARY 2 - Articles excluded after full-text review**

| **1st Author's last name** | **Published** | **Testing For** | **Category** | **RCT** | **Electrical Stimulation** | **DOI** | **Reason for exclusion** |
| --- | --- | --- | --- | --- | --- | --- | --- |
| De Icco | 2018 | nociceptive withdrawal reflex | Pain | Y | Y | 10.1177/0333102417742347 | This study was about cervical VNS. |
| Fang | 2016 | Depression | Psychological | N | Y | 10.1016/j.biopsych.2015.03.025 | This study was not randomized. Therefore, it did not meet the randomized clinical trial inclusion criteria. |
| Hamer | 2019 | Seizure reduction | Drug-resistant epilepsy | Y | Y | 10.1016/j.eplepsyres.2019.02.015 | This article was a summary of Bauer (2016). Bauer (2016) is already included in this review. Hamer does not provide any additional insights on the study design. |
| Kaut | 2019 | Gastrointestinal dysfunctions | Parkinson's disease | Y | Y | 10.3233/NRE-192909 | This study was about cervical VNS. |
| Krasaelap | 2020 | IBS | Gastric | Y | Y, percutaneous | 10.1016/j.cgh.2019.10.012 | This article was a sub-analysis of Kovacic (2017). Kovacic (2017) is already included in this review. Krasaelap does not provide any additional insights on the study design. |
| Lerman | 2016 | cytokines/immunity | Immune | Y | Y | 10.1111/ner.12398 | This study was about cervical VNS. |
| Liu | 2016 | Depression | Psychological | N | Y | 10.1016/j.jad.2016.08.003 | This study was not randomized. Therefore, it did not meet the randomized clinical trial inclusion criteria. |
| Olshan-Perlmutter | 2019 | anxiety and burnout | Psychological | Y | N, Acupressure | 10.1016/j.apnr.2019.05.011 | This study used acupressure as the stimulation method. It meets our exclusion criteria. |
| Rinaldini | 2011 | stress | Psychological | Y | Y, radio electric | 10.1186/1477-7525-9-54 | The main text does not mention vagus nerve stimulation; it does not meet our inclusion criteria. |
| Ruffini | 2015 | Heart rate variability | Cardiac | Y | Y | 10.3389/fnins.2015.00272 | This study was not about auricular VNS. |
| Silberstein | 2016 | chronic migraine | Pain | Y | Y | 10.1212/WNL.0000000000002918 | This study was about cervical VNS. |
| Teckentrup | 2020 | gastric frequency and energy expenditure | Gastric | Y | Y | 10.1016/j.brs.2019.12.018 | This article is not directly clinically relevant. It explores gastric frequency as a marker for taVNS. The authors do not directly link results to a clinical problem. |
| Yu | 2020 | surgery recovery | Pain | Y | Y | 10.1186/s13063-019-3892-4 | This study refers to acupoints; acupressure was a method of stimulation excluded in our review |
| Zamontrisky | 1997 | coronary artery diseases | Cardiac | Y | N, Acupressure | PMID: 9431484 | This study uses acupuncture; acupressure was a method of stimulation excluded in our review |

**Supplementary 3 - User sortable summary of reviewed studies (separate Excel file)**

The Excel presents the entirety of the information extracted from the aVNS trials reviewed. The intention of sharing the Excel is to give readers additional insight into specific parameters, outcomes, and assessments of the studies reviewed within the main article – according to their interest. The reader will find one table per tab.

The first tab, “Overview”, provides an overview of the most pertinent information. The second tab, “All Data (Sortable)”, contains all information extracted from the aVNS trials reviewed. This table can be daunting if viewed by itself. It is therefore recommended to explore the subsequent tabs based on the reader’s research interest – “Results”, “Design”, “RoB Scores”, and “Appendix”. The “Appendix” contains a list of acronyms found throughout this publication.

**SUPPLEMENTARY 4 - Design of controls in non-invasive neuromodulation clinical studies**

Inappropriate implementation of the control group resulted in compromised blinding in many studies and concern for bias in the RoB section ‘deviation from intended intervention’. In this section, design of control in non-invasive neuromodulation studies is discussed. In the following section, methods to evaluate the quality of blinding created by the appropriate use of controls is reviewed.

The design of appropriate controls is difficult for trials testing non-pharmacological interventions - especially so for paresthesia-inducing neuromodulation trials (Robbins and Lipton, 2017). The ideal control group is both therapeutically inert and indistinguishable from the active intervention. A good control emulates the placebo effect – effects from treatment that do not originate from the mechanism of action of the intervention. Where paresthesia is part of the active intervention, which is often the case in non-invasive neuromodulation, an indistinguishable control must also induce paresthesia. Here, we discuss the types of controls used in aVNS clinical trials and their suitability.

**Sham at same location as control (waveform sham).** The ideal control group is to have sham at the same location using a nontherapeutic waveform – providing a similar sensation as the active group but without therapeutic effects. From the subject’s point of view, both the location and general sense of paresthesia is maintained. The danger is that the target nerve might still be recruited and could provide a therapeutic effect. For example, Bauer et al. (2016) and Straube et al. (2015) both used sham at same location as control in their chronic parallel group studies. Both studies used a 25 Hz active group and a 1 Hz waveform sham group. Bauer et al. (2016) reasoned the low dosage of the 1 Hz stimulation could be considered subtherapeutic while Straube et al. (2015) did not provide justification for why 1 Hz stimulation would be a non-therapeutic control. The active control behaved for Bauer (2016) with a mean reduction in seizures of 23.4% in the active 25 Hz group and an increase in 2.9% in the control 1 Hz group. Unfortunately, in Straube (2015) there was a 17.4% reduction in headache frequency in the originally designated active 25 Hz group but a 36.4% reduction in the control 1 Hz group. The active and control group switched from the hypothesized therapeutic outcomes.

To establish an alternative waveform at the same location as non-therapeutic, a pilot study may be performed testing several waveform parameters against a placebo. This can be done as part of the pilot study to explore the parameter space for determination of optimal stimulation parameters. Yarnitsky et al. (2017) report on their pilot crossover design study to find a waveform sham for a therapy to alleviate migraine pain using a non-invasive neuromodulation device on the arm. In Yarnitsky et al. (2019) they report on a successful multicenter study using the sham parameters determined in 2017 – a low ~0.1 Hz frequency that is still perceived. Badran et al. (2018) may be referred to as an aVNS study that investigated several stimulation parameters in a pilot study.

A waveform sham may also be developed by using different stimulation electrodes. For example, Poulsen et al. (2020) show the effects of electrode geometry on fiber recruitment type and area. While this may not apply to aVNS due to the sensory nature of the nerve branch, it may be well suited for non-invasive neurostimulation modalities targeting deeper nerve fibers such as the cervical vagus nerve (Mourdoukoutas et al., 2018).

Waveform shams could be the gold standard for non-invasive neuromodulation trials by providing a sense of paresthesia without therapeutic effects. Sham at same location may therefore be considered for chronic studies, where subject unblinding is of high concern as subjects may question the benefit of a therapy requiring daily compliance. However, given the unknowns surrounding the mechanism of action behind aVNS and the sparse clinical data exploring waveform designs, it may currently be challenging to design nontherapeutic waveforms to achieve a waveform sham. Alternatives are sham with same waveform at a different non-therapeutic location (location sham) and placebo at the same location with no stimulation delivered.

**Sham at a different location as control (location sham).** A common control is to provide therapeutic level stimulation at a different non-vagally innervated location of the auricle. In fact, 16 of 41 RCTs used a location sham at the earlobe or scapha. Waveform parameters are typically kept the same as the active location although stimulation current amplitude might be set to match the sensory perception rather than the absolute current value of the active. It is important for location sham to have some sensation, which might not happen if the absolute current value is ported directly. The danger of using a location sham is that the neuroanatomy of the auricle and the mechanisms of action of auricular stimulation are not understood well enough to make definite determinations of therapeutic versus non-therapeutic areas of the auricle.

Assuming that the earlobe is innervated only by the great auricular nerve, the possible therapeutic effects of stimulating the great auricular nerve come into question. For example, the product Zing by Thync (Los Gatos, CA, USA) claims to target the great auricular nerve to enhance sympathetic nervous system activity. Badran et al. (2018) showed earlobe sham has a non-zero effect on bradycardia and tachycardia rebound after stimulation – cardiac hallmarks of vagus nerve stimulation. Badran et al. (2018) suggested the earlobe may indeed have a physiological response due to anatomical variation or electric field spread and future studies should look at a third off-ear stimulation site. For example, in Addorisio et al. (2019), knowing they couldn’t contain the vibratory stimulation to an off-target location on the auricle, the calf was used as the control stimulation site.

Taken together, caution should be made in assuming the earlobe to be therapeutically inert. Other regions of the ear may be explored, or an off-ear stimulation site may be used. Furthermore, location sham is a poor blind for the investigator and may also be compromised by the subjects in chronic studies, where they may look up the stimulation device for the active location. A location sham may be considered in acute lab settings, especially when a crossover design is used – where a non-paresthesia inducing placebo would easily be identified as non-therapeutic by the subjects.

**Placebo as control.** Lastly, the same location as active but with no stimulation can be used as a placebo control – sometimes with deliberate concealment by informing the subject of a “sub-threshold therapeutic stimulation” or using a device that still beeps or blinks when stimulation would have been delivered. Disclosure on the existence of a control group made explicit in the informed consent may render the concealment less effective. A placebo controls’ contribution to investigator blind and non-therapeutic effect makes it appropriate for use in acute lab settings where a parallel study design is used, where subjects may effectively be cued to believe that a non-paresthesia inducing placebo is therapeutic.

However, in a crossover design where subjects experience both the paresthesia inducing active and the non-paresthesia inducing placebo, they may be less inclined to believe the placebo as a therapeutic intervention. This may show up as a cross over effect between patients who experience the placebo before the active versus the active before the placebo. This may be the case in Cakmak et al. (2017) arm 1, a crossover design testing an active percutaneous stimulation against a surface placebo at the same location. 10 subjects are randomized, presumably allocated equally to active and placebo group first. Individual results presented in their fifth figure shows that subjects 2, 4, 5, 8, and 10 respond best to active intervention and subjects 2, 3, 5, and 8 have the larger placebo responses. It could be that these subjects experienced placebo first and then the active intervention. The patient randomization order is not reported, so a definitive conclusion cannot be drawn. A statistical test for crossover effects would have assuaged concerns. The non-paresthesia inducing nature of a placebo control makes it appropriate in acute parallel studies but not in acute crossover studies.

**Placebo effect from novel therapies.** Doherty and Dieppe (2009) discusses factors that lead to a larger placebo effect including needle-based therapies compared to oral, subject expectations, and how novel the treatment is. The expectation effect behind the placebo response with more novel technologies is termed the ‘halo effect’ and is discussed along with other expectation effects in Olshansky (2007). Overall, this lends to novel neuromodulation devices having larger placebo effects than traditional therapies such as oral drugs. A different therapeutic modality or no intervention as control may therefore fail to account for the strong placebo effects associated with novel medical devices.

Similarly, while the experience of taking a drug is relatively standardized (pill size, color, frequency, etc.) devices are more variable in their user experience and learnings from previous trials on what contributes to placebo effect might not aggregate as well as they do for drugs. While the placebo effect is a nuisance in clinical trial design, it is an eventual benefit for patients in the clinic. Understanding and controlling for the placebo effect during early clinical trials will allow for a therapy’s primary mechanism of action effect to be understood and developed. An optimized placebo effect can then be added to the primary therapeutic mechanism to deliver a final therapy for patients. Given the unknowns surrounding what contributes to the placebo effect in neuromodulation devices, extreme care should be taken in the design of effective controls to account for the effect from placebo in early clinical trials.

**Appropriate control by trial type.** With the discussion on types of controls in non-invasive neuromodulation studies, we may draw some conclusions on appropriate control by trial type. An acute parallel study may use a well concealed placebo. An acute crossover study may consider sham at a different location – its acute nature makes it feasible to design pilot studies to test therapeutic inertness of sham sites. A chronic study should develop a sham at same location if feasible through pilot studies. Eventually, the consistent use of control designs and evaluating of blinding success will grow our understanding of the concealability and therapeutic potency of various control methods.

**Design of control group affects trial outcome.** Blinding continues to be a defining factor of study integrity, and possible lapses in subject blinding due to quality of control provide an opportunity for performance bias to occur (Jüni et al., 2001). In general, one would expect trials that use a placebo control to tend towards reporting larger effect sizes, as they often have more blinding issues (see above). Conversely, trials using sham control might produce a smaller effect size due to improved quality of blinding. In a preliminary analysis of the standardized effect sizes reported in supplementary material 3, a one-tailed t test yielded a p-value of 0.033 between trials using sham and placebo, supporting this conclusion. Similar investigations have been done (Probst et al., 2019; Balk et al., 2002) on the significance of trial outcomes compared to quality of blind and yielded corroborating results. It is evident that the quality and evaluation of study blinding cannot not be overlooked.

**SUPPLEMENTARY 5 - Assessing quality of blinding in non-invasive neuromodulation clinical studies**

Most of the studies reviewed did not assess if the blinding implemented was successful. Evaluating the quality of blinding in subjects assesses if they were aware of the treatment received. Awareness creates bias due to ‘deviation from intended treatment’ (RoB section 2) in the form of a larger placebo effect in the active group and smaller placebo effect in the control group – increasing the overall effect size of the study.

Evaluating the quality of blinding in investigators is also imperative. When unblinded, investigators may bias outcomes when making subjective assessments. Investigators can also bias the measurement of objective outcomes. The provider effect refers to when a patient is influenced by the expectations and behavior of the practitioner (Doherty and Dieppe, 2009). In a double-blind pain study evaluating an agent that increases pain, an agent that decrease pain, and a placebo, it was found that the clinician’s optimism or pessimism towards the efficacy of a treatment had a significant effect on the placebo pain relief received by the subject even with a successful double-blind (Gracely et al., 1985). Similarly, objective measures such as heart rate, blood pressure, and heart rate variability may succumb to the provider effect given the interactions between provider and subject may induce anxiety or relief in the subject.

Several methods to evaluate the quality of blinding in subjects and investigators of a trial are discussed. Namely, objective measure of compliance in chronic studies, crossover effects statistical test in crossover studies, and questionnaires in any study type.

**Compliance to evaluate blinding.** In chronic studies an objective measure of compliance can be used to evaluate blinding success. With the rationale that if a subject was unblinded they would not maintain compliance. The measure of compliance should be objective, such as a stimulation device log, preferably also registering impedance to be within that expected for the subject, as opposed to a device that may be left switched on for the daily required duration.

For example, Bauer et al. (2016), using a waveform sham, reported 88.2% compliance in the active group and 84% in the sham group defined as [stimulation duration]/[prescribed duration of stimulation]. Stavrakis et al. (2020), using a location sham, reported 96%, 80%, and 74% compliance at 1, 3, and 6 months, respectively, in the active group and 85%, 88%, and 82% compliance, respectively, in the control group (p = 0.49 between group comparison). Stavrakis et al. (2020) defined compliance as adherence to daily stimulation (=<4 sessions missed on average per month) based on subjective reporting that was corroborated to cumulative number of hours on stimulation device logs. Straube et al. (2015), using a waveform sham, reported 86% compliance in both the active and sham group to the prescribed 4h of daily stimulation based on daily stimulation time on device logs. Note that the definition of compliance is not consistent between studies and it is unclear if device logs were a cumulative reading averaged over days or a per day reading.

As a caveat, patient compliance may also be influenced by perceived treatment benefit. If a subject does not perceive a benefit from using the device, they will be less inclined to comply as opposed to a subject who perceives a positive benefit from usage. If the active group has a greater number of therapeutic responders, we may expect compliance in the active group to be higher. Large differences in compliance between active and control group not reflected in treatment efficacy are indicative of subject unblinding.

**Carryover effects statistical test to evaluate blinding.** In a crossover study where every subject receives all interventions in a randomized order, unblinding can occur especially when a placebo control is used instead of a sham control. In non-invasive electrical neuromodulation, where paresthesia is a hallmark of therapy, a subject receiving multiple interventions in succession can discriminate between a paresthesia-inducing active intervention and a non-paresthesia inducing control intervention. A subject receiving placebo after an active intervention might show a smaller placebo effect while a subject receiving active after placebo may show a larger active treatment effect.

A statistical test for crossover effects tests if the order of intervention received had an effect on the outcome (Shen and Lu, 2006). It therefore picks up effects of unblinding caused by order of intervention. However, this test also picks up on incomplete washout periods and baseline differences in randomization. Hence, it is important to ensure a sufficient washout period based on literature or preliminary testing as well as to perform a baseline test between randomized groups. For example, De Couck et al. (2017) performed the carryover effects test that showed “randomization was not significantly correlated with HRV at various time points”. Unfortunately, they did not perform a baseline comparison that would allow them to relate the results of the carryover effects test to an evaluation of blinding. In crossover study designs, a statistical test for carryover effects can be used as a proxy to evaluate blinding success if sufficient washout period is ensured and an initial baseline comparison between randomized groups is not significant.

**Questionnaires to evaluate blinding.** The most straightforward way to determine maintenance of blinding is to ask the subject and investigators to guess the assigned treatment group. For example, in a non-invasive non-aVNS neuromodulation study, Yarnitsky et al. (2019) asked subjects upon completion of treatment their presumed group assignment with options of active, sham, and do not know. They detail two types of analyses to examine the effects of perceived assignment on treatment outcome. Kolahi et al. (2009) further discusses the assessment of blinding in clinical trials using questionnaires.

Similar to evaluation of blinding using compliance, perceived effect of treatment can contribute to compromised blinding in the questionnaire method. This can be addressed by deploying the questionnaire after treatment application rather than after treatment completion (Kolahi et al., 2009). Multiple time points are sometimes assessed for trends and compared to treatment efficacy trends. Questionnaires serve as a low-cost and convenient method to assess quality of blinding. Potential confounds include assessment of blinding based on perceived treatment outcome, which can be addressed by deploying the questionnaire at an earlier time point before therapeutic effects are evident.

Methods to assess quality of blinding are summarized in table S5A.

Table S5A: Methods to assess quality of blinding

|  | **Questionnaire** | **Carryover effects test** | **Compliance** |
| --- | --- | --- | --- |
| **Method** | Subject and investigator asked to guess assigned treatment group | Order of intervention (active before control vs. control before active) is tested for significance – if unblinded, subject may show greater placebo effect in active and less in control | Objective measure of compliance (e.g. device logs) used to proxy blinding – unblinded subject would stop complying |
| **Study design** | All | Crossover | Chronic |
| **Subject or investigator blind assessed** | Both | Primarily Subject | Primarily Subject |
| **Potential Confounds** | Guess may be based on perceived treatment effect – addressed by deploying questionnaire at earlier time point before treatment effects are perceived | Baseline imbalance or incomplete washout period also detected by carryover test – randomized groups must be balanced at baseline with sufficient washout period between interventions | Compliance may be based on perceived treatment effect – assess compliance in early part of study before treatment effects are evident |
| **aVNS Study Examples** | Bauer (2016), Stavrakis (2020), Straube (2015) | De Couck (2017) | Hein (2013), Kovacic (2017) |

**Device controlled randomization to improve blinding.** Device controlled randomization could improve both subject and investigator blinding. Using an electrode design that simultaneously covers both the active and sham location, stimulation may be directed to either the active or sham electrode pair by the device. The investigator is blinded to the stimulation location, leaving the device to deliver the randomized protocol such as in Yarnitsky et al. (2017), a non-invasive neuromodulation study. Many of the aVNS studies reviewed were not able to maintain assessor blinding as the electrode location was visible in the case of a location sham or the assessor was the investigator who turned off the stimulation in the case of placebo control. The importance of investigator blind is discussed in the introduction of this section. In addition, the patient maintains the same electrode contact sensation – relevant given the cutaneous sensory nature of the auricular nerves. Device controlled randomization could improve blinding and additionally control for sensation of the electrode on the ear.

**Supplementary 6 – Within versus between group comparison example**

The following hypothetical scenario shows the distinction in a between versus within group analysis and consequently the importance of pre-registering the intended analysis type. Consider a study with twenty samples per group that measures a stimulation induced decrease in heart rate in the active group of 3.1 $\pm$ 2.3 BPM from 83.5 $\pm$ 4.2BPM to 80.4 $\pm$ 5.1 BPM and a decrease in heart rate of 2.2 $\pm$ 1.5 BPM from 84.1 $\pm$ 4.9 BPM to 81.9 $\pm$ 3.7 BPM in the control group. In a between group analysis when the change in the active group of 3.1 $\pm$ 2.3 BPM is compared to the 2.2 $\pm$ 1.5 BPM of the control group, the results are not statistically significant (p = 0.15). However, a within group analysis of the active group comparing the during stimulation heart rate of 80.4 $\pm$ 5.1 BPM to the baseline heart rate of 83.5 $\pm$ 4.2 BPM yields a statistically significant result (p = 0.04). Statistical t-test was done using an online calculator: <https://www.medcalc.org/calc/comparison_of_means.php>

**Supplementary 7 – Time to onset and chronic effects of aVNS**

There are several reports on the transient versus lasting effects of aVNS. Conventional implantable VNS of the cervical trunk is delivered throughout the day for epilepsy patients with an option to increase dosage during a seizure. While stimulation parameters of implantable VNS have been directly ported to aVNS for several clinical indications, an assumption was made that a shorter duration of aVNS will provide the whole day effects of continuous VNS. No aVNS study reviewed cited the intermittent effects of VNS lasting the whole day. Salavatian et al. (2016) reported that the antiarrhythmic effect of cervical implantable VNS in dogs lasts 26 mins on average after stimulation. In aVNS clinical trials, Stowell et al. (2019) in their Fig 2B and 3B showed a quick rebound in blood pressure at 100 Hz when stimulation is switched off. Badran et al. (2018) showed immediate rebound tachycardia when stimulation is turned off. De Couck et al. (2017) had 5 mins rest between left, right, and placebo and shows no effect of randomization order on HRV suggesting that the effects of aVNS on HRV are abated within 5 mins after stimulation. On the other hand, Cakmak et al. (2017) showed tremor reducing effects of aVNS that lasted even 30 mins after stimulation was removed. In Stavrakis et al. (2020) aVNS study on atrial fibrillation, 1 hour of stimulation a day for 6 months reduced atrial fibrillation burden in the active group compared to the sham group. While this lasting effect may be possible, the jump in reasoning is worth highlighting and verifying.

At the same time, it is worth considering that it may take a while for the effects of aVNS to set in. This is plausible given it often takes weeks for the effects of implantable VNS on epilepsy to develop. In Aihua et al. (2014), investigating aVNS for epilepsy, they also reported several weeks for results between groups to reach statistical significance. When considering mechanisms, this suggests plasticity effects or neural remodeling effects instead of an acute circuit modulation. Possible chronic durations of therapy before an effect takes place calls for aVNS animal studies that can be done chronically and where more invasive but sensitive biomarkers can be used in an acute setting.

**Supplementary 8 - Hypothesized mechanisms of action for aVNS**

Several mechanisms have been proposed to explain the clinical effects of aVNS. Firstly, afferent activation of NTS which causes a vago-vagal response. Although it is unclear how somatic input of the ear to NTS modulates other functions, such as cardiac, mediated in the NTS. Intracortical electrophysiological recordings in animals may shine some light on what is happening in the NTS during aVNS. Secondly, direct efferent vagal effects proposed in Badran et al. (2018). Thirdly, activation of further brain structures after NTS such as the locus coeruleus (LC) causing increase in norepinephrine levels mediating analgesia and improved memory. Fourthly, conditioned pain modulation where a remote noxious stimulus produces a generalized analgesic effect as described in a non-invasive non-aVNS study Yarnitsky et al. (2017). Lastly, motor spindle fiber activation as a gateway into the motor control system proposed in Cakmak et al. (2017) by targeting the intrinsic auricular muscle zones of the ear – muscles that shape the ear during development and atrophy in adults. aVNS trials often focus on a single hypothesized mechanism of action and do not always consider the mechanisms proposed by other studies that may have bearing on interpretation of their results. See Cakmak et al. (2019) for a further discussion on the neural circuits modulated during auricular stimulation, including possible recruitment of the auricular branch of the trigeminal nerve, auricular cervical nerves, and intrinsic auricular muscles.

**Supplementary 9 - Individual RoB Rubrics (separate Microsoft word documents)**

During the Cochrane Risk of Bias (RoB) analysis, a rubric was filled for each study reviewed. The rubric details section scores and provides justifications for the scores. Additional justification is found when the suggested algorithm for section or overall score was overridden by the reviewers.

All papers were reviewed using the per-protocol (adherence to treatment analysis) RoB rubric. The aim of a per-protocol analysis and evaluation is to identify whether a treatment effect would occur in optimal conditions. By using the per-protocol analysis, we were not evaluating results from patients excluded by the trials, or with any major protocol deviations. In addition, all RoB rubrics used were from the 2016 update.
