## Supplementary Material 9 - Individual RoB Rubrics for "Auricular Vagus Neuromodulation – A Systematic Review on Quality of Evidence and Clinical Effects": Afanasiev (2016) Parallel (Individually Randomised) - Per Protocol Analysis.docx

### The RoB 2.0 tool (individually randomized, parallel group trials)

| **Assessor name/initials** | RC |
| --- | --- |
| **Study ID and/or reference(s)** | Afanasiev (2016) |

**Study design**

| 🗹 | Randomized parallel group trial |
| --- | --- |
| □ | Cluster-randomized trial |
| □ | Randomized cross-over or other matched design |

| **Specify which outcome is being assessed for risk of bias** | HSP60, HSP70 and 6-minute walk test |
| --- | --- |

| **Specify the numerical result being assessed.** In case of multiple alternative analyses being presented, specify the numeric result (e.g. RR = 1.52 (95% CI 0.83 to 2.77) and/or a reference (e.g. to a table, figure or paragraph) that uniquely defines the result being assessed. | Active group's patient walk distance within 6 minutes significantly increased from 265.4 SD 38.5 to 376.4 SD 39.4 meters (n=63) when compared with the placebo group increased from from 211.4 SD 40.1 to 213.6 SD 38.5 meters (n=7). Hsp 60 increased from 131.5 SD 34.3 to 133.8 SD 34.0 and Hsp 70 increased from 9.4 SD 6.7 to 14.3 SD 12.3 for placebo group. For active subgroup1 (initial HR<80), Hsp 60 increased 103.9 SD 75.1 to 154.3 SD 92.6 and Hsp70 increased from 12.9 SD 5.8 to 20.5 SD 9.1. For active subgroup2 (initial HR>80), Hsp 60 remain the same, before sitmulation is 166.5 SD 97.2 and after stimulation is 165.0 SD 168.7; Hsp70 increased from 12.0 SD 6.8 to 22.4 SD 22.0. |
| --- | --- |

**Is your aim for this study…?**

| □ | to assess the effect of *assignment to intervention* |
| --- | --- |
| 🗹 | to assess the effect of *starting and adhering to intervention* |

**Which of the following sources have you obtained to help inform your risk of bias judgements (tick as many as apply)?**

🗹	Journal article(s) with results of the trial

□	Trial protocol

□	Statistical analysis plan (SAP)

□	Non-commercial trial registry record (e.g. ClinicalTrials.gov record)

□	Company-owned trial registry record (e.g. GSK Clinical Study Register record)

□	“Grey literature” (e.g. unpublished thesis)

□	Conference abstract(s) about the trial

□	Regulatory document (e.g. Clinical Study Report, Drug Approval Package)

□	Research ethics application

□	Grant database summary (e.g. NIH RePORTER, Research Councils UK Gateway to Research)

□	Personal communication with trialist

□	Personal communication with the sponsor

#### Risk of bias assessment for a parallel group trial with interest in the effect of starting and adhering to intervention

| **Domain** | **Signalling questions** | **Response options** | **Description/Support for judgement** |
| --- | --- | --- | --- |
| **Bias arising from the randomization process** | 1.1 Was the allocation sequence random? | Y | Randomly assigned  No reason to suspect revealing the allocation sequence before intervention |
|  | 1.2 Was the allocation sequence concealed until participants were recruited and assigned to interventions? | PY |  |
|  | 1.3 Were there baseline imbalances that suggest a problem with the randomization process? | Y | Large difference in group size design. No significant difference in clinical characteristics but the baseline has a obvious difference in 6-minuts walk test between two groups, but no between group comparison was reported. |
|  | **Risk of bias judgement** | High | From algorithm |
|  | Optional: What is the predicted direction of bias arising from the randomization process? | Blank |  |
| **Bias due to deviations from intended interventions** | 2.1. Were participants aware of their assigned intervention during the trial? | PY | The stimulation procedure has 15 sessions with a total time up to one hours. The placebo has no current and no sensation for all these sessions. |
|  | 2.2. Were carers and trial personnel aware of participants' assigned intervention during the trial? | NI |  |
|  | 2.3. If Y/PY/NI to 2.1 or 2.2: Were important co-interventions balanced across intervention groups? | PN | Placebo rather than sham meant that stimulation sensation was not experienced by control group for entirety of stimulation period |
|  | 2.4. Was the intervention implemented successfully? | Y | No issues reported. |
|  | 2.5. Did study participants adhere to the assigned intervention regimen? | PY | No non-compliance reported. |
|  | 2.6. If N/PN/NI to 2.3, 2.4 or 2.5: Was an appropriate analysis used to estimate the effect of starting and adhering to the intervention? | PY | Analysis appeared sufficient (Mann-Whitney-Wilcoxon test) |
|  | **Risk of bias judgement** | Some concerns | From algorithm |
|  | Optional: What is the predicted direction of bias due to deviations from intended interventions? | Favours experimental |  |
| **Bias due to missing outcome data** | 3.1 Were outcome data available for all, or nearly all, participants randomized? | N | Many data points missing. |
|  | 3.2 If N/PN/NI to 3.1: Are the proportions of missing outcome data and reasons for missing outcome data similar across intervention groups? | N | No reasoning was given. |
|  | 3.3 If N/PN/NI to 3.1: Is there evidence that results were robust to the presence of missing outcome data? | N | No since more close to half of the data points were missing |
|  | **Risk of bias judgement** | High | From algorithm |
|  | Optional: What is the predicted direction of bias due to missing outcome data? | Blank |  |
| **Bias in measurement of the outcome** | 4.1 Were outcome assessors aware of the intervention received by study participants? | NI |  |
|  | 4.2 If Y/PY/NI to 4.1: Was the assessment of the outcome likely to be influenced by knowledge of intervention received? | PN | Physiological and physical measurements not susceptible to change due to observer bias |
|  | **Risk of bias judgement** | Low | From algorithm |
|  | Optional: What is the predicted direction of bias due to measurement of the outcome? | Blank |  |
| **Bias in selection of the reported result** | Are the reported outcome data likely to have been selected, on the basis of the results, from... |  | No evidence indicating selection from multiple outcome measurements, but possibility of selection from multiple time points; intentions not specified |
|  | 5.1. ... multiple outcome measurements (e.g. scales, definitions, time points) within the outcome domain? | NI |  |
|  | 5.2 ... multiple analyses of the data? | NI | Only reported on within group analysis. No between group analysis was reported |
|  | **Risk of bias judgement** | Some concerns | From algorithm |
|  | Optional: What is the predicted direction of bias due to selection of the reported result? | Blank |  |
| **Overall bias** | **Risk of bias judgement** | High | Some issues with differences between control (placebo, n=7) and experimental (n = 53); also No comment on missing outcomes. |
|  | Optional:  What is the overall predicted direction of bias for this outcome? | Favours experimental |  |
