## Supplementary Material 9 - Individual RoB Rubrics for "Auricular Vagus Neuromodulation – A Systematic Review on Quality of Evidence and Clinical Effects": Aihua (2014) Parallel (Individually Randomised) Per Protocol Analysis.docx

### The RoB 2.0 tool (individually randomized, parallel group trials)

| **Assessor name/initials** | JM |
| --- | --- |
| **Study ID and/or reference(s)** | Aihua (2014) |

**Study design**

| 🗹 | Randomized parallel group trial |
| --- | --- |
| □ | Cluster-randomized trial |
| □ | Randomized cross-over or other matched design |

| **Specify which outcome is being assessed for risk of bias** | Seizure frequency |
| --- | --- |

| **Specify the numerical result being assessed.** In case of multiple alternative analyses being presented, specify the numeric result (e.g. RR = 1.52 (95% CI 0.83 to 2.77) and/or a reference (e.g. to a table, figure or paragraph) that uniquely defines the result being assessed. | Seizure frequency reduced by 40% over 12 months |
| --- | --- |

**Is your aim for this study…?**

| □ | to assess the effect of *assignment to intervention* |
| --- | --- |
| 🗹 | to assess the effect of *starting and adhering to intervention* |

**Which of the following sources have you obtained to help inform your risk of bias judgements (tick as many as apply)?**

🗹	Journal article(s) with results of the trial

□	Trial protocol

□	Statistical analysis plan (SAP)

□	Non-commercial trial registry record (e.g. ClinicalTrials.gov record)

□	Company-owned trial registry record (e.g. GSK Clinical Study Register record)

□	“Grey literature” (e.g. unpublished thesis)

□	Conference abstract(s) about the trial

□	Regulatory document (e.g. Clinical Study Report, Drug Approval Package)

□	Research ethics application

□	Grant database summary (e.g. NIH RePORTER, Research Councils UK Gateway to Research)

□	Personal communication with trialist

□	Personal communication with the sponsor

#### Risk of bias assessment for a parallel group trial with interest in the effect of starting and adhering to intervention

| **Domain** | **Signalling questions** | **Response options** | **Description/Support for judgement** |
| --- | --- | --- | --- |
| **Bias arising from the randomization process** | 1.1 Was the allocation sequence random? | PY | Individually randomised  Inferred that allocation sequence was concealed |
|  | 1.2 Was the allocation sequence concealed until participants were recruited and assigned to interventions? | PY |  |
|  | 1.3 Were there baseline imbalances that suggest a problem with the randomization process? | PY | Significant difference between baseline Self-Rating Anxiety Scores (SAS). Experimental scored higher (more anxious) than control. Easier to see decrease in anxiety? All other baseline values seem fine. |
|  | **Risk of bias judgement** | Some concerns | Randomised but some accidental differences in SAS baseline values. |
|  | Optional: What is the predicted direction of bias arising from the randomization process? | Blank |  |
| **Bias due to deviations from intended interventions** | 2.1. Were participants aware of their assigned intervention during the trial? | NI | Patients or their guardians were in charge of stimulation. An internet search would easily show if they were in the control or sham, should the attempt one over the course of the year.  Not specified, difficult to infer given limited involvement in treatment |
|  | 2.2. Were carers and trial personnel aware of participants' assigned intervention during the trial? | NI |  |
|  | 2.3. If Y/PY/NI to 2.1 or 2.2: Were important co-interventions balanced across intervention groups? | PN | Patients on different types of anti-epileptic drugs (AEDs), no specifications as to which. |
|  | 2.4. Was the intervention implemented successfully? | Y | No reported issues |
|  | 2.5. Did study participants adhere to the assigned intervention regimen? | PY | Very heavily up to the participants, but assumed that they followed protocol. |
|  | 2.6. If N/PN/NI to 2.3, 2.4 or 2.5: Was an appropriate analysis used to estimate the effect of starting and adhering to the intervention? | PN | Analysis accounted for number of AEDs being used, but not type or effect of each. |
|  | **Risk of bias judgement** | High | No sham effect observed, and multiple issues with possible breach of participant blinding, AED use, and subsequent statistical analysis of AED effect. |
|  | Optional: What is the predicted direction of bias due to deviations from intended interventions? | Favours experimental |  |
| **Bias due to missing outcome data** | 3.1 Were outcome data available for all, or nearly all, participants randomized? | PY | The only participants whose data was not analysed are those who dropped out of the study due to extreme dizziness, loss of follow-up, or personal reasons. |
|  | 3.2 If N/PN/NI to 3.1: Are the proportions of missing outcome data and reasons for missing outcome data similar across intervention groups? | NA |  |
|  | 3.3 If N/PN/NI to 3.1: Is there evidence that results were robust to the presence of missing outcome data? | NA |  |
|  | **Risk of bias judgement** | Low | No reason to suspect tampering of data to skew results away from null. |
|  | Optional: What is the predicted direction of bias due to missing outcome data? | Blank |  |
| **Bias in measurement of the outcome** | 4.1 Were outcome assessors aware of the intervention received by study participants? | NI | Not stated specifically, important to note that the participants themselves were outcome assessors for many variables (SAS, etc.) |
|  | 4.2 If Y/PY/NI to 4.1: Was the assessment of the outcome likely to be influenced by knowledge of intervention received? | PY | If patients figure out their intervention (see above), easy for SAS and other subjective scores to be affected. |
|  | **Risk of bias judgement** | Some concerns | Dependent on if the patients take it upon themselves to figure out their assignment, possible risk of skewing their self-reported data. |
|  | Optional: What is the predicted direction of bias due to measurement of the outcome? | Blank |  |
| **Bias in selection of the reported result** | Are the reported outcome data likely to have been selected, on the basis of the results, from... |  |  |
|  | 5.1. ... multiple outcome measurements (e.g. scales, definitions, time points) within the outcome domain? | NI | Multiple time points available for analysis, no intentions reported |
|  | 5.2 ... multiple analyses of the data? | NI | Between and within-group analyses were performed, with no intentions reported |
|  | **Risk of bias judgement** | Some concerns | From algorithm |
|  | Optional: What is the predicted direction of bias due to selection of the reported result? | Blank |  |
| **Overall bias** | **Risk of bias judgement** | Some concerns | Overrode from algorithm. Many causes of bias depend on the assumption that control participants took the initiative to research their treatment more thoroughly; difficult to prove or disprove. No compliance reporting available. |
|  | Optional:  What is the overall predicted direction of bias for this outcome? | Blank |  |
