## Supplementary Material 9 - Individual RoB Rubrics for "Auricular Vagus Neuromodulation – A Systematic Review on Quality of Evidence and Clinical Effects": Antonino (2017) Cross-over (Individually Randomised) - Per Protocol Analysis.docx

### The RoB 2.0 tool (individually randomized, cross-over trials)

| **Assessor name/initials** | RC |
| --- | --- |
| **Study ID and/or reference(s)** | Antonino (2017) |

**Study design**

| □ | Randomized parallel group trial |
| --- | --- |
| □ | Cluster-randomized trial |
| 🗹 | Randomized cross-over or other matched design |

| **Specify which outcome is being assessed for risk of bias** | Cardiac baroreflex sensitivity (cBRS is accessed using a spontaneous measure, calculated from the slopes of the systolic Blood Pressure and RR interval); LF/HF ratio of HR variability (a measure of autonomic balance) |
| --- | --- |

| **Specify the numerical result being assessed.** In case of multiple alternative analyses being presented, specify the numeric result (e.g. RR = 1.52 (95% CI 0.83 to 2.77) and/or a reference (e.g. to a table, figure or paragraph) that uniquely defines the result being assessed. | Table 3 and table 6. |
| --- | --- |

**Is your aim for this study…?**

| □ | to assess the effect of *assignment to intervention* |
| --- | --- |
| 🗹 | to assess the effect of *starting and adhering to intervention* |

**Which of the following sources have you obtained to help inform your risk of bias judgements (tick as many as apply)?**

🗹	Journal article(s) with results of the trial

□	Trial protocol

□	Statistical analysis plan (SAP)

□	Non-commercial trial registry record (e.g. ClinicalTrials.gov record)

□	Company-owned trial registry record (e.g. GSK Clinical Study Register record)

□	“Grey literature” (e.g. unpublished thesis)

□	Conference abstract(s) about the trial

□	Regulatory document (e.g. Clinical Study Report, Drug Approval Package)

□	Research ethics application

□	Grant database summary (e.g. NIH RePORTER, Research Councils UK Gateway to Research)

□	Personal communication with trialist

□	Personal communication with the sponsor

#### Risk of bias assessment for a cross-over trial with interest in the effect of starting and adhering to intervention

| **Domain** | **Signalling questions** | **Response options** | **Description/Support for judgement** |
| --- | --- | --- | --- |
| **Bias arising from the randomization process** | 1.1 Was the allocation sequence random? | PY | In the visit one, the participants' body weight and height were assessed, and a familiarization session of tVNS was performed. Then, on visits two, three and four the subjects were exposed in a random order to: 1) active tVNS 2) sham, and 3) placebo. |
|  | 1.2 Was the allocation sequence concealed until participants were recruited and assigned to interventions? | PY |  |
|  | 1.3 Were there baseline imbalances that suggest a problem with the randomization process? | N | Baseline characteristics of the subjects are present in Table 1.No  significant differences are found in any resting physiological vari- ables between active, sham-T and tLS protocols (P > 0.05). During both active tVNS and tLS, the range of the stimulation amplitude was between 40 and 50 mA, with an average of 45 ± 1mA. |
|  | 1.4 Is a roughly equal proportion of participants allocated to each of the two groups? | PY | There are 13 patients that attend all 3 treatment group. No text mentions any missing data. |
|  | 1.5 If N/PN to 1.4: Are period effects included in the analysis? | NA |  |
|  | **Risk of bias judgement** | Low |  |
|  | Optional: What is the predicted direction of bias arising from the randomization process? | Blank |  |
| **Bias due to deviations from intended interventions** | 2.1. Were participants aware of their assigned intervention during each period of the trial? | PN | In the visit one, the participants' body weight and height were assessed, and a familiarization session of tVNS was performed. Then, on visits two, three and four the subjects were exposed in a random order to: 1) active tVNS 2) sham, where electrodes were placed on the tragus of the ear but no current was applied (sham tragus e sham-T), and 3) electrodes were placed on the ear lobe and current applied ac- cording with active tVNS (transcutaneous lobe stimulation etLS).  Fig. 1. Experimental protocol of the study. A. The shaded area shows the distribution of the auricular branch of the vagus nerve to the external ear. B. Position of the electrodes which were placed on the tragus of the ear during active and sham-T protocols. C. Position of the electrodes which were placed on the ear lobe during tLS protocol. |
|  | 2.2. Were carers and trial personnel aware of participants' assigned intervention during each period of the trial? | Y |  |
|  | 2.3. If Y/PY/NI to 2.1 or 2.2: Were important co-interventions balanced across the two interventions? | NI |  |
|  | 2.4. Was the intervention implemented successfully? | PY | No mention of the negative related information. |
|  | 2.5. Did study participants adhere to the assigned intervention regimen? | PY | No mention of the negative related information. |
|  | 2.6. If N/PN/NI to 2.3, 2.4 or 2.5: Was an appropriate analysis used to estimate the effect of starting and adhering to the intervention? | NA |  |
|  | 2.7 Was there sufficient time for any carry-over effects to have disappeared before outcome assessment in the second period? | PY | The washout period was at least 48 hours. No statistical test w |
|  | **Risk of bias judgement** | Low |  |
|  | Optional: What is the predicted direction of bias due to deviations from intended interventions? | Blank |  |
| **Bias due to missing outcome data** | 3.1 Were outcome data available for all, or nearly all, participants randomized? | NI | The data is probably all available to the trialer. But no information is available. |
|  | 3.2 If N/PN/NI to 3.1: Are the proportions of missing outcome data and reasons for missing outcome data similar across interventions? | NI | No information was given. |
|  | 3.3 If N/PN/NI to 3.1: Is there evidence that results were robust to the presence of missing outcome data? | NI | No information was given. |
|  | **Risk of bias judgement** | Some concerns |  |
|  | Optional: What is the predicted direction of bias due to missing outcome data? | Blank |  |
| **Bias in measurement of the outcome** | 4.1 Were outcome assessors aware of the intervention received by study participants? | Y | The outcome assessor is the same person who setup of the experiment protocol for the attendee. |
|  | 4.2 If Y/PY/NI to 4.1: Was the assessment of the outcome likely to be influenced by knowledge of intervention received? | N | Outcome are measurement of heart pulsation and blood pressure which were not accessed subjectively by the assessors. |
|  | **Risk of bias judgement** | Low |  |
|  | Optional: What is the predicted direction of bias due to measurement of the outcome? | Blank |  |
| **Bias in selection of the reported result** | Are the reported outcome data likely to have been selected, on the basis of the results, from... |  |  |
|  | 5.1. ... multiple outcome measurements (e.g. scales, definitions, time points) within the outcome domain? | NI | No information was given. But measurements are taken at multiple times points where potential selections can occur at the basis of outcome interest, in particular, the protocol was not pre-registered. |
|  | 5.2 ... multiple analyses of the data? | NI | No information was given. The reported measurement for baroreflex sensitivity is one of the ways that one can use to analyse the heartbeat data. The study is not pre-registered. |
|  | 5.3 … the outcome of a statistical test for carry-over? | NI | No information was given. |
|  | **Risk of bias judgement** | Some concerns |  |
|  | Optional: What is the predicted direction of bias due to selection of the reported result? | Blank |  |
| **Overall bias** | **Risk of bias judgement** | Some concerns |  |
|  | Optional:  What is the overall predicted direction of bias for this outcome? | Blank |  |
