## Supplementary Material 9 - Individual RoB Rubrics for "Auricular Vagus Neuromodulation – A Systematic Review on Quality of Evidence and Clinical Effects": Bauer (2016) - Parallel (Individually Randomised) - Per Protocol Analysis.docx

### The RoB 2.0 tool (individually randomized, parallel group trials)

| **Assessor name/initials** | MK |
| --- | --- |
| **Study ID and/or reference(s)** | Bauer (2016) |

**Study design**

| 🗹 | Randomized parallel group trial |
| --- | --- |
| □ | Cluster-randomized trial |
| □ | Randomized cross-over or other matched design |

| **Specify which outcome is being assessed for risk of bias** | Reduction in seizure frequencies from baseline compared between active and sham |
| --- | --- |

| **Specify the numerical result being assessed.** In case of multiple alternative analyses being presented, specify the numeric result (e.g. RR = 1.52 (95% CI 0.83 to 2.77) and/or a reference (e.g. to a table, figure or paragraph) that uniquely defines the result being assessed. | Mean seizure frequency reduction per 28 days at end of treatment was –2.9% (SD: in the 1 Hz group [ increase in seizure frequency] and 23.4% in the 25 Hz group [decrease in seizure frequency] (p = 0.146) |
| --- | --- |

**Is your aim for this study…?**

| □ | to assess the effect of *assignment to intervention* |
| --- | --- |
| 🗹 | to assess the effect of *starting and adhering to intervention* |

**Which of the following sources have you obtained to help inform your risk of bias judgements (tick as many as apply)?**

🗹	Journal article(s) with results of the trial

□	Trial protocol

□	Statistical analysis plan (SAP)

□	Non-commercial trial registry record (e.g. ClinicalTrials.gov record)

□	Company-owned trial registry record (e.g. GSK Clinical Study Register record)

□	“Grey literature” (e.g. unpublished thesis)

□	Conference abstract(s) about the trial

□	Regulatory document (e.g. Clinical Study Report, Drug Approval Package)

□	Research ethics application

□	Grant database summary (e.g. NIH RePORTER, Research Councils UK Gateway to Research)

□	Personal communication with trialist

□	Personal communication with the sponsor

#### Risk of bias assessment for a parallel group trial with interest in the effect of starting and adhering to intervention

| **Domain** | **Signalling questions** | **Response options** | **Description/Support for judgement** |
| --- | --- | --- | --- |
| **Bias arising from the randomization process** | 1.1 Was the allocation sequence random? | PY | “This was a randomized, two-arm, parallel group, prospective, double-blind actively controlled study conducted at 9 sites in Germany and 1 site in Austria between 2019 and 2014”  The study states that the allocation sequence was randomized. However, there are no details about the sequence generation process. |
|  |  |  | “This was a randomized, two-arm, parallel group, prospective, double-blind actively controlled study conducted at 9 sites in Germany and 1 site in Austria between 2019 and 2014”  Since it’s double-blind, the patient did not know which group they were allocated to. |
|  | 1.2 Was the allocation sequence concealed until participants were recruited and assigned to interventions? | Y |  |
|  | 1.3 Were there baseline imbalances that suggest a problem with the randomization process? | PN | Based on “Table 1”, I do not see a lot of baseline imbalances. However, though the patients are drug-resistant, it seems like 2/3 were on some type of medication.  The medication is pretty balanced between the group but for the drug called “LEV”, there is more people treated with the 25Hz stimulation (17 vs 9)  Overall, the baseline characteristics are not incompatible with randomization/chance. |
|  | **Risk of bias judgement** | Low | The randomization process seemed fine and it was double blinded. There was nothing concerning. |
|  | Optional: What is the predicted direction of bias arising from the randomization process? | NA |  |
| **Bias due to deviations from intended interventions** | 2.1. Were participants aware of their assigned intervention during the trial? | PN | “Since tVNS causes a tingling sensation in the skin area touched by the electrode contacts, an active control was necessary to ensure blinding”  Both groups of patients had the same stimulation device. They just received stimulation at different frequencies.  They did not evaluate blinding so PN. They still measured compliance and it was very high. |
|  | 2.2. Were carers and trial personnel aware of participants' assigned intervention during the trial? | N | “This was a randomized, two-arm, parallel group, prospective, double-blind actively controlled study conducted at 9 sites in Germany and 1 site in Austria between 2019 and 2014”  The carers and trial personnel were supposed to be blinded. The only things that they could control was the intensity of the current. |
|  | 2.3. If Y/PY/NI to 2.1 or 2.2: Were important co-interventions balanced across intervention groups? | NA |  |
|  | 2.4. Was the intervention implemented successfully? | PY | “ The stimulation device recorded intensity and daily percentage of target duration achieved.”  “Treatment compliance was on a similar high level in both groups.” |
|  | 2.5. Did study participants adhere to the assigned intervention regimen? | Y | Reference Table 2.  Treatment compliance was high. |
|  | 2.6. If N/PN/NI to 2.3, 2.4 or 2.5: Was an appropriate analysis used to estimate the effect of starting and adhering to the intervention? | NA |  |
|  | **Risk of bias judgement** | Low | Treatment compliance were high and there were no important co-interventions. |
|  | Optional: What is the predicted direction of bias due to deviations from intended interventions? | NA |  |
| **Bias due to missing outcome data** | 3.1 Were outcome data available for all, or nearly all, participants randomized? | PY | “For patients with missing seizure-frequency per 28 days at end of the treatment period the last available seizure frequency per 28 days was used (last-observation-carried-forward method).”  It is not specified how many patients this was done for.  “Seventy-six patients were randomized to the high-level (n = 37) or low-level (n = 39) stimulation group. The study was completed by 58 patients (76 %); 8 patients of the 1 Hz group and 10 patients of the 25 Hz group prematurely discontinued the study.”  It was mITT |
|  | 3.2 If N/PN/NI to 3.1: Are the proportions of missing outcome data and reasons for missing outcome data similar across intervention groups? | NA |  |
|  | 3.3 If N/PN/NI to 3.1: Is there evidence that results were robust to the presence of missing outcome data? | NA |  |
|  | **Risk of bias judgement** | Low | The study was mITT |
|  | Optional: What is the predicted direction of bias due to missing outcome data? | NA |  |
| **Bias in measurement of the outcome** | 4.1 Were outcome assessors aware of the intervention received by study participants? | PN | “Seizure frequency was prospectively recorded in patient diaries.” The patients were blinded. |
|  | 4.2 If Y/PY/NI to 4.1: Was the assessment of the outcome likely to be influenced by knowledge of intervention received? | NA |  |
|  | **Risk of bias judgement** | Low | Low |
|  | Optional: What is the predicted direction of bias due to measurement of the outcome? | NA |  |
| **Bias in selection of the reported result** | Are the reported outcome data likely to have been selected, on the basis of the results, from... |  | “The study had been registered in the German Clinical Trials Register (ID: DRKS00003689)”  It was pre-registered and the outcome measurements were outlined |
|  | 5.1. ... multiple outcome measurements (e.g. scales, definitions, time points) within the outcome domain? | PN | They accounted for all data, even non-per protocol data as it was mITT. |
|  | 5.2 ... multiple analyses of the data? | PN | Primary outcome was pre-registered and  Baseline groups both had more individuals get better (100% reduction of seizure) and worsen (see figure 4)  “Mean seizure reduction per 28 days at end of treatment was –2.9% in the 1 Hz group and 23.4% in the 25 Hz group (p = 0.146).” is misleading |
|  | **Risk of bias judgement** | Low | The study was mITT |
|  | Optional: What is the predicted direction of bias due to selection of the reported result? | NA |  |
| **Overall bias** | **Risk of bias judgement** | Low | It was a well-designed study. The reporting of data seemed honest**.** If they had used EEG videos instead of seizure diaries, they might have seen more significant results |
|  | Optional:  What is the overall predicted direction of bias for this outcome? | NA |  |
