## Supplementary Material 9 - Individual RoB Rubrics for "Auricular Vagus Neuromodulation – A Systematic Review on Quality of Evidence and Clinical Effects": Borges (2019) Cross-over (Individually Randomised) - Per Protocol Analysis.docx

### The RoB 2.0 tool (individually randomized, cross-over trials)

| **Assessor name/initials** | JM |
| --- | --- |
| **Study ID and/or reference(s)** |  |

**Study design**

| □ | Randomized parallel group trial |
| --- | --- |
| □ | Cluster-randomized trial |
| 🗹 | Randomized cross-over or other matched design |

| **Specify which outcome is being assessed for risk of bias** | Root mean square of successive differences (RMSSD) |
| --- | --- |

| **Specify the numerical result being assessed.** In case of multiple alternative analyses being presented, specify the numeric result (e.g. RR = 1.52 (95% CI 0.83 to 2.77) and/or a reference (e.g. to a table, figure or paragraph) that uniquely defines the result being assessed. | Experiment 1 compared 0.5 mA, 1.0 mA, and 1.5 mA stimulation intensities, and experiment 2 compared set (unchangeable by user) and free (set by user) stimulation, but neither could be regarded as RCT. Experiment 3 showed no significant difference between resting and stimulation first half (61.64 SD 23.57 to 63.79 SD 25.20, p=0.034), significant difference between resting and stimulation second half (61.64 SD 23.57 to 65.19 SD 26.90, p=0.003), but no significant effects of stimulation condition (p=0.860) or method (p=0.334), nor interaction effect (p=0.759) |
| --- | --- |

**Is your aim for this study…?**

| □ | to assess the effect of *assignment to intervention* |
| --- | --- |
| 🗹 | to assess the effect of *starting and adhering to intervention* |

**Which of the following sources have you obtained to help inform your risk of bias judgements (tick as many as apply)?**

🗹	Journal article(s) with results of the trial

□	Trial protocol

□	Statistical analysis plan (SAP)

□	Non-commercial trial registry record (e.g. ClinicalTrials.gov record)

□	Company-owned trial registry record (e.g. GSK Clinical Study Register record)

□	“Grey literature” (e.g. unpublished thesis)

□	Conference abstract(s) about the trial

□	Regulatory document (e.g. Clinical Study Report, Drug Approval Package)

□	Research ethics application

□	Grant database summary (e.g. NIH RePORTER, Research Councils UK Gateway to Research)

□	Personal communication with trialist

□	Personal communication with the sponsor

#### Risk of bias assessment for a cross-over trial with interest in the effect of starting and adhering to intervention

| **Domain** | **Signalling questions** | **Response options** | **Description/Support for judgement** |
| --- | --- | --- | --- |
| **Bias arising from the randomization process** | 1.1 Was the allocation sequence random? | Y | Random assignment to each arm and the order of treatment conditions within each arm  No reason to suspect that the allocation sequence was revealed prematurely |
|  | 1.2 Was the allocation sequence concealed until participants were recruited and assigned to interventions? | PY |  |
|  | 1.3 Were there baseline imbalances that suggest a problem with the randomization process? | NI | None reported |
|  | 1.4 Is a roughly equal proportion of participants allocated to each of the two groups? | Y | Around 60 per group |
|  | 1.5 If N/PN to 1.4: Are period effects included in the analysis? | NA |  |
|  | **Risk of bias judgement** | Low | Randomization project was adequate |
|  | Optional: What is the predicted direction of bias arising from the randomization process? | Blank |  |
| **Bias due to deviations from intended interventions** | 2.1. Were participants aware of their assigned intervention during each period of the trial? | PN | Patients were blinded to intervention in sham vs experimental  Specified as “single blind” |
|  | 2.2. Were carers and trial personnel aware of participants' assigned intervention during each period of the trial? | PY |  |
|  | 2.3. If Y/PY/NI to 2.1 or 2.2: Were important co-interventions balanced across the two interventions? | Y | No unbalance reported |
|  | 2.4. Was the intervention implemented successfully? | Y | No intervention issues reported |
|  | 2.5. Did study participants adhere to the assigned intervention regimen? | Y | No non-compliance reported on the part of the study participants |
|  | 2.6. If N/PN/NI to 2.3, 2.4 or 2.5: Was an appropriate analysis used to estimate the effect of starting and adhering to the intervention? | NA |  |
|  | 2.7 Was there sufficient time for any carry-over effects to have disappeared before outcome assessment in the second period? | PN | Some evidence for the existence of carry-over effects in experiment 3 |
|  | **Risk of bias judgement** | Some concerns | No sham effect relative to experimental which was strange, also some evidence for carry-over |
|  | Optional: What is the predicted direction of bias due to deviations from intended interventions? | Favours experimental | Possibility for carry-over to add to and increase subsequent trial effect |
| **Bias due to missing outcome data** | 3.1 Were outcome data available for all, or nearly all, participants randomized? | Y | No missing participants from analysis |
|  | 3.2 If N/PN/NI to 3.1: Are the proportions of missing outcome data and reasons for missing outcome data similar across interventions? | NA |  |
|  | 3.3 If N/PN/NI to 3.1: Is there evidence that results were robust to the presence of missing outcome data? | NA |  |
|  | **Risk of bias judgement** | Low | All outcome data appeared to be present |
|  | Optional: What is the predicted direction of bias due to missing outcome data? | Blank |  |
| **Bias in measurement of the outcome** | 4.1 Were outcome assessors aware of the intervention received by study participants? | PY | Single-blind specified |
|  | 4.2 If Y/PY/NI to 4.1: Was the assessment of the outcome likely to be influenced by knowledge of intervention received? | PN | Measurement of RMSSD unlikely to be affected |
|  | **Risk of bias judgement** | Low | Measurements were adequately taken to avoid bias |
|  | Optional: What is the predicted direction of bias due to measurement of the outcome? | Blank |  |
| **Bias in selection of the reported result** | Are the reported outcome data likely to have been selected, on the basis of the results, from... |  | Use of multiple time points (resting vs 1^st^ half vs 2^nd^ half) |
|  | 5.1. ... multiple outcome measurements (e.g. scales, definitions, time points) within the outcome domain? | NI |  |
|  | 5.2 ... multiple analyses of the data? | NI | Multiple post-hoc analyses were performed, though they were reported seemingly non-selectively |
|  | 5.3 … the outcome of a statistical test for carry-over? | PN | Tests for carry-over were performed, but no outcomes appear to have been selected solely based on test results |
|  | **Risk of bias judgement** | Some concerns | From algorithm |
|  | Optional: What is the predicted direction of bias due to selection of the reported result? | Unpredictable |  |
| **Overall bias** | **Risk of bias judgement** | High | Serious concerns with the fact that HRV values were only collected after familiarization stimulation; not really a baseline since measurements taken after some stim had already occurred. Additional issues with only aggregate statistics reporting for RMSSD and lack of intention specificity |
|  | Optional:  What is the overall predicted direction of bias for this outcome? | Unpredictable |  |
