## Supplementary Material 9 - Individual RoB Rubrics for "Auricular Vagus Neuromodulation – A Systematic Review on Quality of Evidence and Clinical Effects": Busch (2013) Cross-over (Individually Randomised) - Per Protocol Analysis.docx

### The RoB 2.0 tool (individually randomized, cross-over trials)

| **Assessor name/initials** | JM |
| --- | --- |
| **Study ID and/or reference(s)** | Busch (2013) |

**Study design**

| □ | Randomized parallel group trial |
| --- | --- |
| □ | Cluster-randomized trial |
| 🗹 | Randomized cross-over or other matched design |

| **Specify which outcome is being assessed for risk of bias** | Cold detection threshold (CDT), warm detection threshold (WDT), thermal sensory limen (TSL), cold pain threshold (CPT), heat pain threshold (HPT), mechanical detection threshold (MDT), mechanical pain threshold (MPS), wind up ratio (WUR), pressure pain threshold (PPT), tonic heat pain (THP) |
| --- | --- |

| **Specify the numerical result being assessed.** In case of multiple alternative analyses being presented, specify the numeric result (e.g. RR = 1.52 (95% CI 0.83 to 2.77) and/or a reference (e.g. to a table, figure or paragraph) that uniquely defines the result being assessed. | Significant bilateral THP decrease (19.49 SD 9.87 to 15.16 SD 8.14 ipsilateral, 20.06 SD 9.83 to 15.14 SD 8.35 contralateral, p<0.001), ipsilateral MPT decrease (197.9 SD 176.28 to 169.93 SD 156.95, p=0.008), ipsilateral MPS decrease (2.72 SD 4.36 to 2.09 SD 2.77 p=0.014), contralateral PPT decrease (449.01 SD 171.88 to 431.51 SD 146.37, p=0.037) , MDT side effect (during stim 1.59 SD 1.85 ipsilateral vs 1.47 SD 2.23, p=0.037) All other non-significant |
| --- | --- |

**Is your aim for this study…?**

| □ | to assess the effect of *assignment to intervention* |
| --- | --- |
| 🗹 | to assess the effect of *starting and adhering to intervention* |

**Which of the following sources have you obtained to help inform your risk of bias judgements (tick as many as apply)?**

🗹	Journal article(s) with results of the trial

□	Trial protocol

□	Statistical analysis plan (SAP)

□	Non-commercial trial registry record (e.g. ClinicalTrials.gov record)

□	Company-owned trial registry record (e.g. GSK Clinical Study Register record)

□	“Grey literature” (e.g. unpublished thesis)

□	Conference abstract(s) about the trial

□	Regulatory document (e.g. Clinical Study Report, Drug Approval Package)

□	Research ethics application

□	Grant database summary (e.g. NIH RePORTER, Research Councils UK Gateway to Research)

□	Personal communication with trialist

□	Personal communication with the sponsor

#### Risk of bias assessment for a cross-over trial with interest in the effect of starting and adhering to intervention

| **Domain** | **Signalling questions** | **Response options** | **Description/Support for judgement** |
| --- | --- | --- | --- |
| **Bias arising from the randomization process** | 1.1 Was the allocation sequence random? | Y | Individually randomized  No reason to suspect pre-emptive sequence revealing |
|  | 1.2 Was the allocation sequence concealed until participants were recruited and assigned to interventions? | Y |  |
|  | 1.3 Were there baseline imbalances that suggest a problem with the randomization process? | NI | Differences in baseline values were used as covariates but their significance was not clearly reported |
|  | 1.4 Is a roughly equal proportion of participants allocated to each of the two groups? | Y | Randomly assigned, half in each group |
|  | 1.5 If N/PN to 1.4: Are period effects included in the analysis? | NA |  |
|  | **Risk of bias judgement** | Low | From algorithm |
|  | Optional: What is the predicted direction of bias arising from the randomization process? | Blank |  |
| **Bias due to deviations from intended interventions** | 2.1. Were participants aware of their assigned intervention during each period of the trial? | PY | The control was a no current placebo; difference in perception could alter results  Experimenters were careful to maintain double-blind |
|  | 2.2. Were carers and trial personnel aware of participants' assigned intervention during each period of the trial? | PN |  |
|  | 2.3. If Y/PY/NI to 2.1 or 2.2: Were important co-interventions balanced across the two interventions? | PN | Placebo did not replicate sensations that those in active group would receive. |
|  | 2.4. Was the intervention implemented successfully? | PY | No reported issues with intervention implementation |
|  | 2.5. Did study participants adhere to the assigned intervention regimen? | N | Two participants were non-compliant; their data was removed and they were replaced with two new participants at the end of the study |
|  | 2.6. If N/PN/NI to 2.3, 2.4 or 2.5: Was an appropriate analysis used to estimate the effect of starting and adhering to the intervention? | PY | As long as the new participants adhered to intervention, it seems like they solve the problem created by the non-compliant ones |
|  | 2.7 Was there sufficient time for any carry-over effects to have disappeared before outcome assessment in the second period? | PY | 48 hours is likely sufficient |
|  | **Risk of bias judgement** | Some concerns | Placebo brings up issues of blinding, participant non-compliance an issue as well |
|  | Optional: What is the predicted direction of bias due to deviations from intended interventions? | Favours experimental |  |
| **Bias due to missing outcome data** | 3.1 Were outcome data available for all, or nearly all, participants randomized? | Y | Only noncompliant were not analyzed |
|  | 3.2 If N/PN/NI to 3.1: Are the proportions of missing outcome data and reasons for missing outcome data similar across interventions? | NA |  |
|  | 3.3 If N/PN/NI to 3.1: Is there evidence that results were robust to the presence of missing outcome data? | NA |  |
|  | **Risk of bias judgement** | Low | From algorithm |
|  | Optional: What is the predicted direction of bias due to missing outcome data? | Blank |  |
| **Bias in measurement of the outcome** | 4.1 Were outcome assessors aware of the intervention received by study participants? | PN | Investigators were careful to maintain double blind |
|  | 4.2 If Y/PY/NI to 4.1: Was the assessment of the outcome likely to be influenced by knowledge of intervention received? | NA |  |
|  | **Risk of bias judgement** | Low | From algorithm |
|  | Optional: What is the predicted direction of bias due to measurement of the outcome? | Blank |  |
| **Bias in selection of the reported result** | Are the reported outcome data likely to have been selected, on the basis of the results, from... |  |  |
|  | 5.1. ... multiple outcome measurements (e.g. scales, definitions, time points) within the outcome domain? | PN | Reported results were appropriately recorded corresponding to intended outcome measurements |
|  | 5.2 ... multiple analyses of the data? | NI | Multiple analyses were performed, intentions were not specified |
|  | 5.3 … the outcome of a statistical test for carry-over? | PN | No reason to suspect reporting of results from only one period of the cross-over in any case |
|  | **Risk of bias judgement** | Some concerns | From algorithm |
|  | Optional: What is the predicted direction of bias due to selection of the reported result? | Blank |  |
| **Overall bias** | **Risk of bias judgement** | Some concerns | Some issues with blinding, non-compliance. Otherwise well-constructed and executed study |
|  | Optional:  What is the overall predicted direction of bias for this outcome? | Favours experimental |  |
