## Supplementary Material 9 - Individual RoB Rubrics for "Auricular Vagus Neuromodulation – A Systematic Review on Quality of Evidence and Clinical Effects": Cakmak (2017) Cross-over (Individually Randomised) - Per Protocol Analysis.docx

### The RoB 2.0 tool (individually randomized, cross-over trials)

| **Assessor name/initials** | NV |
| --- | --- |
| **Study ID and/or reference(s)** | Cakmak (2017) |

**Study design**

| □ | Randomized parallel group trial |
| --- | --- |
| □ | Cluster-randomized trial |
| 🗹 | Randomized cross-over or other matched design |

| **Specify which outcome is being assessed for risk of bias** | Change in UPDRS III score after intervention. |
| --- | --- |

| **Specify the numerical result being assessed.** In case of multiple alternative analyses being presented, specify the numeric result (e.g. RR = 1.52 (95% CI 0.83 to 2.77) and/or a reference (e.g. to a table, figure or paragraph) that uniquely defines the result being assessed. | 5.3 UPDRS III improvement in active, 1.5 in placebo, and 0.2 in different location sham. |
| --- | --- |

**Is your aim for this study…?**

| □ | to assess the effect of *assignment to intervention* |
| --- | --- |
| 🗹 | to assess the effect of *starting and adhering to intervention* |

**Which of the following sources have you obtained to help inform your risk of bias judgements (tick as many as apply)?**

🗹	Journal article(s) with results of the trial

□	Trial protocol

□	Statistical analysis plan (SAP)

□	Non-commercial trial registry record (e.g. ClinicalTrials.gov record)

□	Company-owned trial registry record (e.g. GSK Clinical Study Register record)

□	“Grey literature” (e.g. unpublished thesis)

□	Conference abstract(s) about the trial

□	Regulatory document (e.g. Clinical Study Report, Drug Approval Package)

□	Research ethics application

□	Grant database summary (e.g. NIH RePORTER, Research Councils UK Gateway to Research)

🗹	Personal communication with trialist

□	Personal communication with the sponsor

#### Risk of bias assessment for a cross-over trial with interest in the effect of starting and adhering to intervention

| **Domain** | **Signalling questions** | **Response options** | **Description/Support for judgement** |
| --- | --- | --- | --- |
| **Bias arising from the randomization process** | 1.1 Was the allocation sequence random? | PY | “Randomised” mentioned in consort flow chart but no mention in main body text.  Inferring that randomization sequence was concealed. |
|  | 1.2 Was the allocation sequence concealed until participants were recruited and assigned to interventions? | PY |  |
|  | 1.3 Were there baseline imbalances that suggest a problem with the randomization process? | PN | Individual patient disease severity and dopaminergic drug dose reported are comparable. |
|  | 1.4 Is a roughly equal proportion of participants allocated to each of the two groups? | Y | Closest integer split possible. |
|  | 1.5 If N/PN to 1.4: Are period effects included in the analysis? | NA |  |
|  | **Risk of bias judgement** | Low | Randomization not detailed but inferred to be low bias. |
|  | Optional: What is the predicted direction of bias arising from the randomization process? | Blank |  |
| **Bias due to deviations from intended interventions** | 2.1. Were participants aware of their assigned intervention during each period of the trial? | PY | Crossover study design with difference in perception and location between active, placebo, and sham likely influenced extent of the placebo/sham effect. Participants were cued to expect a therapeutic subthreshold stimulation.  Motor score evaluators were well blinded, but the same evaluators were used in arm 1 and 2 increasing likelihood of compromising the blind. |
|  | 2.2. Were carers and trial personnel aware of participants' assigned intervention during each period of the trial? | PN |  |
|  | 2.3. If Y/PY/NI to 2.1 or 2.2: Were important co-interventions balanced across the two interventions? | PN | A subject experiencing active after placebo versus placebo after active in the crossover design will likely have varying placebo/sham effects. Statistical test not done to test for this effect. Note how individual data shows strong placebo/sham responders tend to be weaker active responders. |
|  | 2.4. Was the intervention implemented successfully? | Y | Successful adherence to protocol reported. |
|  | 2.5. Did study participants adhere to the assigned intervention regimen? | Y | Acute study. No mention of non-compliance from participants. |
|  | 2.6. If N/PN/NI to 2.3, 2.4 or 2.5: Was an appropriate analysis used to estimate the effect of starting and adhering to the intervention? | NI | All participants adhered to the intervention. However, effect of intervention order in crossover design on UPDRS III improvement was not investigated. |
|  | 2.7 Was there sufficient time for any carry-over effects to have disappeared before outcome assessment in the second period? | NI | Washout period not mentioned despite a sustained effect 30 mins after a 20 mins intervention. |
|  | **Risk of bias judgement** | High |  |
|  | Optional: What is the predicted direction of bias due to deviations from intended interventions? | Favours experimental |  |
| **Bias due to missing outcome data** | 3.1 Were outcome data available for all, or nearly all, participants randomized? | Y | Individual results are also reported. |
|  | 3.2 If N/PN/NI to 3.1: Are the proportions of missing outcome data and reasons for missing outcome data similar across interventions? | NA |  |
|  | 3.3 If N/PN/NI to 3.1: Is there evidence that results were robust to the presence of missing outcome data? | NA |  |
|  | **Risk of bias judgement** | Low |  |
|  | Optional: What is the predicted direction of bias due to missing outcome data? |  |  |
| **Bias in measurement of the outcome** | 4.1 Were outcome assessors aware of the intervention received by study participants? | PN | Motor score evaluators were well blinded, but the same evaluators were used in arm 1 and 2 increasing likelihood of compromising the blind. |
|  | 4.2 If Y/PY/NI to 4.1: Was the assessment of the outcome likely to be influenced by knowledge of intervention received? | NA |  |
|  | **Risk of bias judgement** | Low |  |
|  | Optional: What is the predicted direction of bias due to measurement of the outcome? |  |  |
| **Bias in selection of the reported result** | Are the reported outcome data likely to have been selected, on the basis of the results, from... |  | Only single measurements taken. |
|  | 5.1. ... multiple outcome measurements (e.g. scales, definitions, time points) within the outcome domain? | N |  |
|  | 5.2 ... multiple analyses of the data? | PN | Before and after intervention measurements were made for each group. Statistical significance was claimed based on before and after active intervention. For arm 2, significance between improvement of active versus improvement of control was also reported. Pre-registration not done. |
|  | 5.3 … the outcome of a statistical test for carry-over? | PN | Carry-over effects not mentioned. |
|  | **Risk of bias judgement** | Low |  |
|  | Optional: What is the predicted direction of bias due to selection of the reported result? | Blank |  |
| **Overall bias** | **Risk of bias judgement** | Some concerns | Overrode “High” from algorithm. Despite subject blinding concerns, study is well designed and reported. |
|  | Optional:  What is the overall predicted direction of bias for this outcome? | Blank |  |
