## Supplementary Material 9 - Individual RoB Rubrics for "Auricular Vagus Neuromodulation – A Systematic Review on Quality of Evidence and Clinical Effects": De Couck (2017) Cross-over (Individually Randomised) - Per Protocol Analysis.docx

### The RoB 2.0 tool (individually randomized, cross-over trials)

| **Assessor name/initials** | NV |
| --- | --- |
| **Study ID and/or reference(s)** | De Couck (2017) |

**Study design**

| □ | Randomized parallel group trial |
| --- | --- |
| □ | Cluster-randomized trial |
| 🗹 | Randomized cross-over or other matched design |

| **Specify which outcome is being assessed for risk of bias** | Study 1 HRV measurements |
| --- | --- |

| **Specify the numerical result being assessed.** In case of multiple alternative analyses being presented, specify the numeric result (e.g. RR = 1.52 (95% CI 0.83 to 2.77) and/or a reference (e.g. to a table, figure or paragraph) that uniquely defines the result being assessed. | Study 1 HRV results |
| --- | --- |

**Is your aim for this study…?**

| □ | to assess the effect of *assignment to intervention* |
| --- | --- |
| 🗹 | to assess the effect of *starting and adhering to intervention* |

**Which of the following sources have you obtained to help inform your risk of bias judgements (tick as many as apply)?**

🗹	Journal article(s) with results of the trial

□	Trial protocol

□	Statistical analysis plan (SAP)

□	Non-commercial trial registry record (e.g. ClinicalTrials.gov record)

□	Company-owned trial registry record (e.g. GSK Clinical Study Register record)

□	“Grey literature” (e.g. unpublished thesis)

□	Conference abstract(s) about the trial

□	Regulatory document (e.g. Clinical Study Report, Drug Approval Package)

□	Research ethics application

□	Grant database summary (e.g. NIH RePORTER, Research Councils UK Gateway to Research)

□	Personal communication with trialist

□	Personal communication with the sponsor

#### Risk of bias assessment for a cross-over trial with interest in the effect of starting and adhering to intervention

| **Domain** | **Signalling questions** | **Response options** | **Description/Support for judgement** |
| --- | --- | --- | --- |
| **Bias arising from the randomization process** | 1.1 Was the allocation sequence random? | Y | Method not detailed. 6 randomization orders possible (left, right, and placeb0)  Concealment assumed |
|  | 1.2 Was the allocation sequence concealed until participants were recruited and assigned to interventions? | PY |  |
|  | 1.3 Were there baseline imbalances that suggest a problem with the randomization process? | PN | Statistical test for randomization order on HRV results done and not significant so randomization likely unbiased |
|  | 1.4 Is a roughly equal proportion of participants allocated to each of the two groups? | Y |  |
|  | 1.5 If N/PN to 1.4: Are period effects included in the analysis? | NA |  |
|  | **Risk of bias judgement** | Low |  |
|  | Optional: What is the predicted direction of bias arising from the randomization process? | Blank |  |
| **Bias due to deviations from intended interventions** | 2.1. Were participants aware of their assigned intervention during each period of the trial? | PN | Statistical test for randomization order on HRV results done and not significant so participants likely did not realise placebo, if they had placebo randomised first would have responded differently from placebo randomised 2^nd^/last. |
|  | 2.2. Were carers and trial personnel aware of participants' assigned intervention during each period of the trial? | Y |  |
|  | 2.3. If Y/PY/NI to 2.1 or 2.2: Were important co-interventions balanced across the two interventions? | PY |  |
|  | 2.4. Was the intervention implemented successfully? | PY |  |
|  | 2.5. Did study participants adhere to the assigned intervention regimen? | Y |  |
|  | 2.6. If N/PN/NI to 2.3, 2.4 or 2.5: Was an appropriate analysis used to estimate the effect of starting and adhering to the intervention? | NA |  |
|  | 2.7 Was there sufficient time for any carry-over effects to have disappeared before outcome assessment in the second period? | PY | Washout period is much shorter than other aVNS studies. However, a statistical test for randomization order on HRV results done and not significant suggesting no carryover effects (order does not matter) |
|  | **Risk of bias judgement** | Low | Although placebo versus baseline (essentially same condition) were different for SDNN (p = 0.025) non significant with Bonferroni correction of alpha = 0.05/4 |
|  | Optional: What is the predicted direction of bias due to deviations from intended interventions? | Blank |  |
| **Bias due to missing outcome data** | 3.1 Were outcome data available for all, or nearly all, participants randomized? | Y |  |
|  | 3.2 If N/PN/NI to 3.1: Are the proportions of missing outcome data and reasons for missing outcome data similar across interventions? | NA |  |
|  | 3.3 If N/PN/NI to 3.1: Is there evidence that results were robust to the presence of missing outcome data? | NA |  |
|  | **Risk of bias judgement** | Low |  |
|  | Optional: What is the predicted direction of bias due to missing outcome data? | Blank |  |
| **Bias in measurement of the outcome** | 4.1 Were outcome assessors aware of the intervention received by study participants? | Y |  |
|  | 4.2 If Y/PY/NI to 4.1: Was the assessment of the outcome likely to be influenced by knowledge of intervention received? | PN | Objective ECG measure. However, assessor could influence subject. |
|  | **Risk of bias judgement** | Low |  |
|  | Optional: What is the predicted direction of bias due to measurement of the outcome? | Blank |  |
| **Bias in selection of the reported result** | Are the reported outcome data likely to have been selected, on the basis of the results, from... |  | All collected data was commented on. In cases of non significance, details of data were not given. Example, no breathing rate data is provided. |
|  | 5.1. ... multiple outcome measurements (e.g. scales, definitions, time points) within the outcome domain? | PN |  |
|  | 5.2 ... multiple analyses of the data? | NI | Comparison was done to baseline rather than to sham. Not pre-registered. |
|  | 5.3 … the outcome of a statistical test for carry-over? | PN | Carryover test was done, and no significant effect was reported. |
|  | **Risk of bias judgement** | Some concerns |  |
|  | Optional: What is the predicted direction of bias due to selection of the reported result? | Blank |  |
| **Overall bias** | **Risk of bias judgement** | Some concerns |  |
|  | Optional:  What is the overall predicted direction of bias for this outcome? | Blank |  |
