## Supplementary Material 9 - Individual RoB Rubrics for "Auricular Vagus Neuromodulation – A Systematic Review on Quality of Evidence and Clinical Effects": Fisher (2018) Cross-over (Individually Randomised) - Per Protocol Analysis.docx

### The RoB 2.0 tool (individually randomized, cross-over trials)

| **Assessor name/initials** | NV |
| --- | --- |
| **Study ID and/or reference(s)** | Fisher (2018) |

**Study design**

| □ | Randomized parallel group trial |
| --- | --- |
| □ | Cluster-randomized trial |
| 🗹 | Randomized cross-over or other matched design |

| **Specify which outcome is being assessed for risk of bias** | Percentage decrease in median systolic blood pressure (SBP). Blood pressure measured using non-invasive finger cuff. |
| --- | --- |

| **Specify the numerical result being assessed.** In case of multiple alternative analyses being presented, specify the numeric result (e.g. RR = 1.52 (95% CI 0.83 to 2.77) and/or a reference (e.g. to a table, figure or paragraph) that uniquely defines the result being assessed. | For responders (9 of 18; 8 excluded due to "data quality") increase in median SBP compared to baseline was 0.86% (SD 8.34) for active and 7.94% (SD 7.06) in placebo. Active SBP increased less. |
| --- | --- |

**Is your aim for this study…?**

| □ | to assess the effect of *assignment to intervention* |
| --- | --- |
| 🗹 | to assess the effect of *starting and adhering to intervention* |

**Which of the following sources have you obtained to help inform your risk of bias judgements (tick as many as apply)?**

🗹	Journal article(s) with results of the trial

□	Trial protocol

□	Statistical analysis plan (SAP)

□	Non-commercial trial registry record (e.g. ClinicalTrials.gov record)

□	Company-owned trial registry record (e.g. GSK Clinical Study Register record)

□	“Grey literature” (e.g. unpublished thesis)

□	Conference abstract(s) about the trial

□	Regulatory document (e.g. Clinical Study Report, Drug Approval Package)

□	Research ethics application

□	Grant database summary (e.g. NIH RePORTER, Research Councils UK Gateway to Research)

□	Personal communication with trialist

□	Personal communication with the sponsor

#### Risk of bias assessment for a cross-over trial with interest in the effect of starting and adhering to intervention

| **Domain** | **Signalling questions** | **Response options** | **Description/Support for judgement** |
| --- | --- | --- | --- |
| **Bias arising from the randomization process** | 1.1 Was the allocation sequence random? | PY | Randomisation mentioned but details not mentioned.  Allocation sequence was likely concealed, although not mentioned. |
|  | 1.2 Was the allocation sequence concealed until participants were recruited and assigned to interventions? | PY |  |
|  | 1.3 Were there baseline imbalances that suggest a problem with the randomization process? | NI | Not tested or reported. |
|  | 1.4 Is a roughly equal proportion of participants allocated to each of the two groups? | NI | Not mentioned. |
|  | 1.5 If N/PN to 1.4: Are period effects included in the analysis? | NA |  |
|  | **Risk of bias judgement** | Some concerns |  |
|  | Optional: What is the predicted direction of bias arising from the randomization process? | Blank |  |
| **Bias due to deviations from intended interventions** | 2.1. Were participants aware of their assigned intervention during each period of the trial? | PY | Participations were likely aware due to differing levels of perception (medium, low, and none).  Investigators not blinded. |
|  | 2.2. Were carers and trial personnel aware of participants' assigned intervention during each period of the trial? | Y |  |
|  | 2.3. If Y/PY/NI to 2.1 or 2.2: Were important co-interventions balanced across the two interventions? | N | N. Crossover design not tested for significance of intervention order on outcome. Autonomic tasks performed before and after stimulation in this study not controlled for. |
|  | 2.4. Was the intervention implemented successfully? | PN | Data from 8 of 18 subjects had to be removed, suggesting measurement or intervention were largely unsuccessful. |
|  | 2.5. Did study participants adhere to the assigned intervention regimen? | PY | Single session intervention resulting in likely high adherence. |
|  | 2.6. If N/PN/NI to 2.3, 2.4 or 2.5: Was an appropriate analysis used to estimate the effect of starting and adhering to the intervention? | N | Co-intervention of autonomic task was not tracked. |
|  | 2.7 Was there sufficient time for any carry-over effects to have disappeared before outcome assessment in the second period? | NI | Carryover effects not discussed |
|  | **Risk of bias judgement** | High | Large sham effect, attempted explanation is suppressive effect of active on increasing SBP. |
|  | Optional: What is the predicted direction of bias due to deviations from intended interventions? | Blank |  |
| **Bias due to missing outcome data** | 3.1 Were outcome data available for all, or nearly all, participants randomized? | N | Data from 8 of 18 subjects was excluded “through a preliminary preprocessing of the data based on quality” |
|  | 3.2 If N/PN/NI to 3.1: Are the proportions of missing outcome data and reasons for missing outcome data similar across interventions? | PN | Reason given is not strong and relation of missing data to intervention order is not reported. |
|  | 3.3 If N/PN/NI to 3.1: Is there evidence that results were robust to the presence of missing outcome data? | NI | Not discussed. |
|  | **Risk of bias judgement** | High |  |
|  | Optional: What is the predicted direction of bias due to missing outcome data? |  |  |
| **Bias in measurement of the outcome** | 4.1 Were outcome assessors aware of the intervention received by study participants? | Y | Investigators not blinded |
|  | 4.2 If Y/PY/NI to 4.1: Was the assessment of the outcome likely to be influenced by knowledge of intervention received? | PN | SBP measurement is made automatically with readout. However investigator behaviour may have influenced patient and hence outcome. |
|  | **Risk of bias judgement** | Some concerns |  |
|  | Optional: What is the predicted direction of bias due to measurement of the outcome? | Blank |  |
| **Bias in selection of the reported result** | Are the reported outcome data likely to have been selected, on the basis of the results, from... |  |  |
|  | 5.1. ... multiple outcome measurements (e.g. scales, definitions, time points) within the outcome domain? | PY | Each 15 mins session (baseline, stimulation, recovery) is split into three 5 mins windows, any of which could have been used. Systolic vs diastolic vs mean pressure. |
|  | 5.2 ... multiple analyses of the data? | PY | Data could have been analysed compared to baseline or comparing before and after intervention or recovery. Several were done and reported with no corrections for multiple group comparisons. |
|  | 5.3 … the outcome of a statistical test for carry-over? | NI | Carryover effects not discussed. |
|  | **Risk of bias judgement** | High | Poorly designed, conducted, and reported study. |
|  | Optional: What is the predicted direction of bias due to selection of the reported result? | Blank |  |
| **Overall bias** | **Risk of bias judgement** | High |  |
|  | Optional:  What is the overall predicted direction of bias for this outcome? | Blank |  |
