## Supplementary Material 9 - Individual RoB Rubrics for "Auricular Vagus Neuromodulation – A Systematic Review on Quality of Evidence and Clinical Effects": Frokjaer (2016) Cross-over (Individually Randomised) - Per Protocol Analysis.docx

### The RoB 2.0 tool (individually randomized, cross-over trials)

| **Assessor name/initials** | NV |
| --- | --- |
| **Study ID and/or reference(s)** | Frokjaer (2016) |

**Study design**

| □ | Randomized parallel group trial |
| --- | --- |
| □ | Cluster-randomized trial |
| 🗹 | Randomized cross-over or other matched design |

| **Specify which outcome is being assessed for risk of bias** | ECG (HR, cardiac vagal tone), PPG finger cuff (SBP, DBP, MBP), pain threshold (right quadricep muscle and right tibia bone), cold pressor test (conditioned pain modulation), and drink test with ultrasoudn imaging (gastric emptying rate, frequency of antral contractions, amplitude of antral contractions) of active compared to sham. |
| --- | --- |

| **Specify the numerical result being assessed.** In case of multiple alternative analyses being presented, specify the numeric result (e.g. RR = 1.52 (95% CI 0.83 to 2.77) and/or a reference (e.g. to a table, figure or paragraph) that uniquely defines the result being assessed. | Table 1 in text. Effect of aVNS versus deep breathing cannot be isolated. |
| --- | --- |

**Is your aim for this study…?**

| □ | to assess the effect of *assignment to intervention* |
| --- | --- |
| 🗹 | to assess the effect of *starting and adhering to intervention* |

**Which of the following sources have you obtained to help inform your risk of bias judgements (tick as many as apply)?**

🗹	Journal article(s) with results of the trial

□	Trial protocol

□	Statistical analysis plan (SAP)

□	Non-commercial trial registry record (e.g. ClinicalTrials.gov record)

□	Company-owned trial registry record (e.g. GSK Clinical Study Register record)

□	“Grey literature” (e.g. unpublished thesis)

□	Conference abstract(s) about the trial

□	Regulatory document (e.g. Clinical Study Report, Drug Approval Package)

□	Research ethics application

□	Grant database summary (e.g. NIH RePORTER, Research Councils UK Gateway to Research)

□	Personal communication with trialist

□	Personal communication with the sponsor

#### Risk of bias assessment for a cross-over trial with interest in the effect of starting and adhering to intervention

| **Domain** | **Signalling questions** | **Response options** | **Description/Support for judgement** |
| --- | --- | --- | --- |
| **Bias arising from the randomization process** | 1.1 Was the allocation sequence random? | Y | Randomised using centrally generated randomised link at a website.  Inferred. |
|  | 1.2 Was the allocation sequence concealed until participants were recruited and assigned to interventions? | PY |  |
|  | 1.3 Were there baseline imbalances that suggest a problem with the randomization process? | PY | Baseline imbalance in muscle pain threshold. Baseline measurements not taken for several parameters. |
|  | 1.4 Is a roughly equal proportion of participants allocated to each of the two groups? | Y | Crossover study, all completed. |
|  | 1.5 If N/PN to 1.4: Are period effects included in the analysis? | NA |  |
|  | **Risk of bias judgement** | Some concerns |  |
|  | Optional: What is the predicted direction of bias arising from the randomization process? | Blank |  |
| **Bias due to deviations from intended interventions** | 2.1. Were participants aware of their assigned intervention during each period of the trial? | PN | Sham for both stimulation and breathing. Although sham breathing did not receive continuous feedback from investigator.  Not investigator blinded. |
|  | 2.2. Were carers and trial personnel aware of participants' assigned intervention during each period of the trial? | Y |  |
|  | 2.3. If Y/PY/NI to 2.1 or 2.2: Were important co-interventions balanced across the two interventions? | PN | Sham breathing did not receive continuous feedback from investigator. Active deep breathing was “guide(ed) the subjects” |
|  | 2.4. Was the intervention implemented successfully? | PY | Reported as successful |
|  | 2.5. Did study participants adhere to the assigned intervention regimen? | Y | Short term intervention. |
|  | 2.6. If N/PN/NI to 2.3, 2.4 or 2.5: Was an appropriate analysis used to estimate the effect of starting and adhering to the intervention? | NI | Statistical analysis was preliminary and appropriately intended for an exploratory study. Co-intervention applied to all in active group. |
|  | 2.7 Was there sufficient time for any carry-over effects to have disappeared before outcome assessment in the second period? | PY | At least 1 week between active and sham. |
|  | **Risk of bias judgement** | Some concerns |  |
|  | Optional: What is the predicted direction of bias due to deviations from intended interventions? | Blank |  |
| **Bias due to missing outcome data** | 3.1 Were outcome data available for all, or nearly all, participants randomized? | PY | All data available except 4 of 18 for ultrasound due to inadequate visualization.  Does this mean 4 bad recordings or 8 (cross over)? |
|  | 3.2 If N/PN/NI to 3.1: Are the proportions of missing outcome data and reasons for missing outcome data similar across interventions? | NI | Not specified which group missing data is from. |
|  | 3.3 If N/PN/NI to 3.1: Is there evidence that results were robust to the presence of missing outcome data? | PY | Antral contraction frequency signal is strong (although no baseline comparison) |
|  | **Risk of bias judgement** | Low |  |
|  | Optional: What is the predicted direction of bias due to missing outcome data? | Blank |  |
| **Bias in measurement of the outcome** | 4.1 Were outcome assessors aware of the intervention received by study participants? | Y | Not investigator blinded |
|  | 4.2 If Y/PY/NI to 4.1: Was the assessment of the outcome likely to be influenced by knowledge of intervention received? | PN | Most metrics were not subjective for the assessor. Although assessor behaviour could influence participants such as their response to the pain threshold. |
|  | **Risk of bias judgement** | Low |  |
|  | Optional: What is the predicted direction of bias due to measurement of the outcome? | Blank |  |
| **Bias in selection of the reported result** | Are the reported outcome data likely to have been selected, on the basis of the results, from... |  | Outcomes were measured at multiple time points and the study was not pre-registered. |
|  | 5.1. ... multiple outcome measurements (e.g. scales, definitions, time points) within the outcome domain? | NI |  |
|  | 5.2 ... multiple analyses of the data? | NI | Analysis could be done between sham and active as well as baseline and later time point. Baseline imbalances could also be adjusted for. |
|  | 5.3 … the outcome of a statistical test for carry-over? | PN | Statistical test for carryover not performed but likely no carryover due to 1 week washout period. |
|  | **Risk of bias judgement** | Some concerns |  |
|  | Optional: What is the predicted direction of bias due to selection of the reported result? | Blank |  |
| **Overall bias** | **Risk of bias judgement** | High | Largely due to concern of increased sham effect when feedback on breathing is given, measurement of many variables at several time points without correction for multiple comparison, and unadjusted baseline imbalances. |
|  | Optional:  What is the overall predicted direction of bias for this outcome? | Blank |  |
