## Supplementary Material 9 - Individual RoB Rubrics for "Auricular Vagus Neuromodulation – A Systematic Review on Quality of Evidence and Clinical Effects": Hein (2013) Parallel (Individually Randomised) - Per Protocol Analysis.docx

### The RoB 2.0 tool (individually randomized, parallel group trials)

| **Assessor name/initials** | JM |
| --- | --- |
| **Study ID and/or reference(s)** | Hein (2013) |

**Study design**

| 🗹 | Randomized parallel group trial |
| --- | --- |
| □ | Cluster-randomized trial |
| □ | Randomized cross-over or other matched design |

| **Specify which outcome is being assessed for risk of bias** | Within and between-group Hamilton Depression Rating Scale (HAMD) - (clinician rated) good against patient bias, but improvements in scale subcategories such as sleep, anxiety, or appetite can mistakenly categorize a stimulus as antidepressant if only overall score is accounted for. Significant effect size should be >=0.40 (PMID: 16889106). Beck's Depression Inventory (BDI) (self-scored) has issues with patient exaggeration/minimization, and is designed as more of a screening device than a diagnostic tool; similar issues to HAMD with mistaken antidepressant categorization due to physical category improvement (doi:10.1093/pubmed/fdi03)) |
| --- | --- |

| **Specify the numerical result being assessed.** In case of multiple alternative analyses being presented, specify the numeric result (e.g. RR = 1.52 (95% CI 0.83 to 2.77) and/or a reference (e.g. to a table, figure or paragraph) that uniquely defines the result being assessed. | Within-group: Study 1: BDI decrease from 27.0 to 14.0 (p<0.000 [typo?], change = -13.0 SD 6.7), HAMD decreased from 15.4 to 10.6 (no significance, change = -4.8 SD 6.5). Study 2: BDI decreased from 29.4 to 17.4 (p<0.05, change = -12.0 SD 4.7) and HAMD decreased from 18.4 to 12.1 (no significance, change = -6.3 SD 4.0). Between group (only reported for pooled results): significant BDI difference (active -12.6 SD 6.0, control -4.4 SD 9.9, p = 0.004) but no HAMD difference |
| --- | --- |

**Is your aim for this study…?**

| □ | to assess the effect of *assignment to intervention* |
| --- | --- |
| 🗹 | to assess the effect of *starting and adhering to intervention* |

**Which of the following sources have you obtained to help inform your risk of bias judgements (tick as many as apply)?**

🗹	Journal article(s) with results of the trial

□	Trial protocol

□	Statistical analysis plan (SAP)

□	Non-commercial trial registry record (e.g. ClinicalTrials.gov record)

□	Company-owned trial registry record (e.g. GSK Clinical Study Register record)

□	“Grey literature” (e.g. unpublished thesis)

□	Conference abstract(s) about the trial

□	Regulatory document (e.g. Clinical Study Report, Drug Approval Package)

□	Research ethics application

□	Grant database summary (e.g. NIH RePORTER, Research Councils UK Gateway to Research)

□	Personal communication with trialist

□	Personal communication with the sponsor

#### Risk of bias assessment for a parallel group trial with interest in the effect of starting and adhering to intervention

| **Domain** | **Signalling questions** | **Response options** | **Description/Support for judgement** |
| --- | --- | --- | --- |
| **Bias arising from the randomization process** | 1.1 Was the allocation sequence random? | Y | Specified as randomised.  No reason to suspect pre-emptive allocation sequence revealing. |
|  | 1.2 Was the allocation sequence concealed until participants were recruited and assigned to interventions? | PY |  |
|  | 1.3 Were there baseline imbalances that suggest a problem with the randomization process? | PN | No reason to suspect baseline imbalances |
|  | **Risk of bias judgement** | Low | From algorithm |
|  | Optional: What is the predicted direction of bias arising from the randomization process? | Blank |  |
| **Bias due to deviations from intended interventions** | 2.1. Were participants aware of their assigned intervention during the trial? | PY | When asked about which intervention they thought they were in, 30-40% of control group participants stated they believed they had not really been in the stimulation group.  Single-blind study. |
|  | 2.2. Were carers and trial personnel aware of participants' assigned intervention during the trial? | Y |  |
|  | 2.3. If Y/PY/NI to 2.1 or 2.2: Were important co-interventions balanced across intervention groups? | PY | active group was stimulated at setting below sensory threshold, so no-current placebo would match lack of sensation |
|  | 2.4. Was the intervention implemented successfully? | Y | No reported issues |
|  | 2.5. Did study participants adhere to the assigned intervention regimen? | Y | No reported non-compliance |
|  | 2.6. If N/PN/NI to 2.3, 2.4 or 2.5: Was an appropriate analysis used to estimate the effect of starting and adhering to the intervention? | NA |  |
|  | **Risk of bias judgement** | Low | From algorithm |
|  | Optional: What is the predicted direction of bias due to deviations from intended interventions? | Blank |  |
| **Bias due to missing outcome data** | 3.1 Were outcome data available for all, or nearly all, participants randomized? | Y | No evidence to believe that data were withheld |
|  | 3.2 If N/PN/NI to 3.1: Are the proportions of missing outcome data and reasons for missing outcome data similar across intervention groups? | NA |  |
|  | 3.3 If N/PN/NI to 3.1: Is there evidence that results were robust to the presence of missing outcome data? | NA |  |
|  | **Risk of bias judgement** | Low |  |
|  | Optional: What is the predicted direction of bias due to missing outcome data? | Blank |  |
| **Bias in measurement of the outcome** | 4.1 Were outcome assessors aware of the intervention received by study participants? | NI | Not stated that they were blinded or not. |
|  | 4.2 If Y/PY/NI to 4.1: Was the assessment of the outcome likely to be influenced by knowledge of intervention received? | PY | Hamilton Depression Rating Scale (HAMD) has definite risks for bias of assessors impacting ratings given for patients |
|  | **Risk of bias judgement** | Some concerns | Not stated whether assessors were blind, additionally, depression rating inventories can be biased relative to assessors’ beliefs. However, interesting to note that there were no significant differences between HAMD control and experimental. |
|  | Optional: What is the predicted direction of bias due to measurement of the outcome? | Favours experimental |  |
| **Bias in selection of the reported result** | Are the reported outcome data likely to have been selected, on the basis of the results, from... |  |  |
|  | 5.1. ... multiple outcome measurements (e.g. scales, definitions, time points) within the outcome domain? | PN | Only two metrics (HAMD and BDI) were used |
|  | 5.2 ... multiple analyses of the data? | NI | Multiple analyses were performed, no intentions specified |
|  | **Risk of bias judgement** | Some concerns | From algorithm |
|  | Optional: What is the predicted direction of bias due to selection of the reported result? | Unpredictable |  |
| **Overall bias** | **Risk of bias judgement** | Some concerns | Concerns with participant unblinding, assessor unblinding (which was never stated) |
|  | Optional:  What is the overall predicted direction of bias for this outcome? | Favours experimental |  |
