## Supplementary Material 9 - Individual RoB Rubrics for "Auricular Vagus Neuromodulation – A Systematic Review on Quality of Evidence and Clinical Effects": Huang (2014) Parallel (Individually Randomised) - Per Protocol Analysis.docx

### The RoB 2.0 tool (individually randomized, parallel group trials)

| **Assessor name/initials** | JM |
| --- | --- |
| **Study ID and/or reference(s)** | Huang 2014) |

**Study design**

| 🗹 | Randomized parallel group trial |
| --- | --- |
| □ | Cluster-randomized trial |
| □ | Randomized cross-over or other matched design |

| **Specify which outcome is being assessed for risk of bias** | Between-group 2hPG levels (2-hr plasma glucose - patient is fasts for 12 hours, then eats 75 g of carbohydrates. 2 hours after this, blood plasma glucose readings taken) |
| --- | --- |

| **Specify the numerical result being assessed.** In case of multiple alternative analyses being presented, specify the numeric result (e.g. RR = 1.52 (95% CI 0.83 to 2.77) and/or a reference (e.g. to a table, figure or paragraph) that uniquely defines the result being assessed. | Measurements taken at baseline, 6 weeks, and 12 weeks. Repeated measures ANOVA showed significant decrease in 2hPG between active (9.7 SD 1.2 baseline, 7.3 SD 1.8 6 week, 7.5 SD 1.3 12 week) relative to sham (9.1 SD 1.2, 8.0 SD 1.6, 8.0 SD 1.4) (p=0.004) and relative to control (9.3 SD 1.1, 9.5 SD 1.9, 10.0 SD 2.7)(p<0.01) |
| --- | --- |

**Is your aim for this study…?**

| □ | to assess the effect of *assignment to intervention* |
| --- | --- |
| 🗹 | to assess the effect of *starting and adhering to intervention* |

**Which of the following sources have you obtained to help inform your risk of bias judgements (tick as many as apply)?**

🗹	Journal article(s) with results of the trial

□	Trial protocol

□	Statistical analysis plan (SAP)

□	Non-commercial trial registry record (e.g. ClinicalTrials.gov record)

□	Company-owned trial registry record (e.g. GSK Clinical Study Register record)

□	“Grey literature” (e.g. unpublished thesis)

□	Conference abstract(s) about the trial

□	Regulatory document (e.g. Clinical Study Report, Drug Approval Package)

□	Research ethics application

□	Grant database summary (e.g. NIH RePORTER, Research Councils UK Gateway to Research)

□	Personal communication with trialist

□	Personal communication with the sponsor

#### Risk of bias assessment for a parallel group trial with interest in the effect of starting and adhering to intervention

| **Domain** | **Signalling questions** | **Response options** | **Description/Support for judgement** |
| --- | --- | --- | --- |
| **Bias arising from the randomization process** | 1.1 Was the allocation sequence random? | Y | Randomly computationally generated.  Assignments kept in envelopes until interventions delivered. |
|  | 1.2 Was the allocation sequence concealed until participants were recruited and assigned to interventions? | Y |  |
|  | 1.3 Were there baseline imbalances that suggest a problem with the randomization process? | PY | One instance of significant difference in baseline systolic BP in analysis of variance between the 3 groups (p=0.01) |
|  | **Risk of bias judgement** | Some concerns | From algorithm |
|  | Optional: What is the predicted direction of bias arising from the randomization process? | Unpredictable |  |
| **Bias due to deviations from intended interventions** | 2.1. Were participants aware of their assigned intervention during the trial? | PY | Participants were kept blinded; however, possible that they could have figured out allocation after 12 weeks.  Only single-blinded. |
|  | 2.2. Were carers and trial personnel aware of participants' assigned intervention during the trial? | PY |  |
|  | 2.3. If Y/PY/NI to 2.1 or 2.2: Were important co-interventions balanced across intervention groups? | PY | Sham and active sensation were balanced |
|  | 2.4. Was the intervention implemented successfully? | Y | No reported issues |
|  | 2.5. Did study participants adhere to the assigned intervention regimen? | PY | 2 dropped out due to reported dizziness from the taVNS, but unlikely to affect outcomes |
|  | 2.6. If N/PN/NI to 2.3, 2.4 or 2.5: Was an appropriate analysis used to estimate the effect of starting and adhering to the intervention? | PY | The analysis performed make sense and seem sufficient. |
|  | **Risk of bias judgement** | Low | From algorithm. |
|  | Optional: What is the predicted direction of bias due to deviations from intended interventions? | Blank |  |
| **Bias due to missing outcome data** | 3.1 Were outcome data available for all, or nearly all, participants randomized? | Y | All data were included (ITT) |
|  | 3.2 If N/PN/NI to 3.1: Are the proportions of missing outcome data and reasons for missing outcome data similar across intervention groups? | NA |  |
|  | 3.3 If N/PN/NI to 3.1: Is there evidence that results were robust to the presence of missing outcome data? | NA |  |
|  | **Risk of bias judgement** | Low | From algorithm |
|  | Optional: What is the predicted direction of bias due to missing outcome data? | Blank |  |
| **Bias in measurement of the outcome** | 4.1 Were outcome assessors aware of the intervention received by study participants? | Y | Single blind; assessors were not blinded |
|  | 4.2 If Y/PY/NI to 4.1: Was the assessment of the outcome likely to be influenced by knowledge of intervention received? | PN | Measurements were either biological markers or objective calculations, unlikely to have been affected by bias. |
|  | **Risk of bias judgement** | Low | From algorithm |
|  | Optional: What is the predicted direction of bias due to measurement of the outcome? | Blank |  |
| **Bias in selection of the reported result** | Are the reported outcome data likely to have been selected, on the basis of the results, from... |  | Multiple time points available for analysis, intentions not specified. |
|  | 5.1. ... multiple outcome measurements (e.g. scales, definitions, time points) within the outcome domain? | NI |  |
|  | 5.2 ... multiple analyses of the data? | NI | Multiple analyses performed between active, sham, and control groups; intentions not specified |
|  | **Risk of bias judgement** | Some concerns | From algorithm |
|  | Optional: What is the predicted direction of bias due to selection of the reported result? | Unpredictable |  |
| **Overall bias** | **Risk of bias judgement** | Some concerns | From algorithm |
|  | Optional:  What is the overall predicted direction of bias for this outcome? | Unpredictable |  |
