## Supplementary Material 9 - Individual RoB Rubrics for "Auricular Vagus Neuromodulation – A Systematic Review on Quality of Evidence and Clinical Effects": Johnson (1991) Parallel (Individually Randomised) - Per Protocol Analysis.docx

### The RoB 2.0 tool (individually randomized, parallel group trials)

| **Assessor name/initials** | NV |
| --- | --- |
| **Study ID and/or reference(s)** | Johnson (1991) |

**Study design**

| 🗹 | Randomized parallel group trial |
| --- | --- |
| □ | Cluster-randomized trial |
| □ | Randomized cross-over or other matched design |

| **Specify which outcome is being assessed for risk of bias** | Electrical pain threshold at ipsilateral (right) index finger (pain threshold recorded in mA above sensory threshold) delivered by carbon rubber electrodes and autonomic function (temperature, systolic and diastolic blood pressure, heart rate). Difference before and after stimulation compared between groups. |
| --- | --- |

| **Specify the numerical result being assessed.** In case of multiple alternative analyses being presented, specify the numeric result (e.g. RR = 1.52 (95% CI 0.83 to 2.77) and/or a reference (e.g. to a table, figure or paragraph) that uniquely defines the result being assessed. | Figure 2 and 3. |
| --- | --- |

**Is your aim for this study…?**

| □ | to assess the effect of *assignment to intervention* |
| --- | --- |
| 🗹 | to assess the effect of *starting and adhering to intervention* |

**Which of the following sources have you obtained to help inform your risk of bias judgements (tick as many as apply)?**

🗹	Journal article(s) with results of the trial

□	Trial protocol

□	Statistical analysis plan (SAP)

□	Non-commercial trial registry record (e.g. ClinicalTrials.gov record)

□	Company-owned trial registry record (e.g. GSK Clinical Study Register record)

□	“Grey literature” (e.g. unpublished thesis)

□	Conference abstract(s) about the trial

□	Regulatory document (e.g. Clinical Study Report, Drug Approval Package)

□	Research ethics application

□	Grant database summary (e.g. NIH RePORTER, Research Councils UK Gateway to Research)

□	Personal communication with trialist

□	Personal communication with the sponsor

#### Risk of bias assessment for a parallel group trial with interest in the effect of starting and adhering to intervention

| **Domain** | **Signalling questions** | **Response options** | **Description/Support for judgement** |
| --- | --- | --- | --- |
| **Bias arising from the randomization process** | 1.1 Was the allocation sequence random? | PY | Mentions randomised but no details on method.  Concealment assumed. |
|  | 1.2 Was the allocation sequence concealed until participants were recruited and assigned to interventions? | PY |  |
|  | 1.3 Were there baseline imbalances that suggest a problem with the randomization process? | NI | Baseline comparison not done. Difference in initial pain threshold between active and placebo, but not statistically significant. |
|  | **Risk of bias judgement** | Some concerns |  |
|  | Optional: What is the predicted direction of bias arising from the randomization process? | Blank |  |
| **Bias due to deviations from intended interventions** | 2.1. Were participants aware of their assigned intervention during the trial? | PN | PN. Blinding assumed, not mentioned. Placebo was likely aware that they were control.  Blinding not mentioned. |
|  | 2.2. Were carers and trial personnel aware of participants' assigned intervention during the trial? | Y |  |
|  | 2.3. If Y/PY/NI to 2.1 or 2.2: Were important co-interventions balanced across intervention groups? | PN | PN. Placebo group did not have the sensation of stimulation that is likely associated with increased sham effect. |
|  | 2.4. Was the intervention implemented successfully? | PY | Not reported otherwise. |
|  | 2.5. Did study participants adhere to the assigned intervention regimen? | PY | Not reported otherwise. Short-term study. |
|  | 2.6. If N/PN/NI to 2.3, 2.4 or 2.5: Was an appropriate analysis used to estimate the effect of starting and adhering to the intervention? | NA |  |
|  | **Risk of bias judgement** | High |  |
|  | Optional: What is the predicted direction of bias due to deviations from intended interventions? | Blank |  |
| **Bias due to missing outcome data** | 3.1 Were outcome data available for all, or nearly all, participants randomized? | Y | Not reported otherwise. |
|  | 3.2 If N/PN/NI to 3.1: Are the proportions of missing outcome data and reasons for missing outcome data similar across intervention groups? | NA / Y / PY / PN / N / NI |  |
|  | 3.3 If N/PN/NI to 3.1: Is there evidence that results were robust to the presence of missing outcome data? | NA / Y / PY / PN / N / NI |  |
|  | **Risk of bias judgement** | Low |  |
|  | Optional: What is the predicted direction of bias due to missing outcome data? | Blank |  |
| **Bias in measurement of the outcome** | 4.1 Were outcome assessors aware of the intervention received by study participants? | Y | Blinding not mentioned in report. Subject blind assumed by reviewer. |
|  | 4.2 If Y/PY/NI to 4.1: Was the assessment of the outcome likely to be influenced by knowledge of intervention received? | NI | Manual measurements of blood pressure and heart rate were taken. Investigators could also cue subjects to a response. |
|  | **Risk of bias judgement** | Some concerns |  |
|  | Optional: What is the predicted direction of bias due to measurement of the outcome? | Blank |  |
| **Bias in selection of the reported result** | Are the reported outcome data likely to have been selected, on the basis of the results, from... |  | Selecting times for analysis seems to have been done after data collection. |
|  | 5.1. ... multiple outcome measurements (e.g. scales, definitions, time points) within the outcome domain? | NI |  |
|  | 5.2 ... multiple analyses of the data? | PN | Both within and between group analysis reported on. |
|  | **Risk of bias judgement** | Some concerns |  |
|  | Optional: What is the predicted direction of bias due to selection of the reported result? | Blank |  |
| **Overall bias** | **Risk of bias judgement** | High |  |
|  | Optional:  What is the overall predicted direction of bias for this outcome? | Blank |  |
