## Supplementary Material 9 - Individual RoB Rubrics for "Auricular Vagus Neuromodulation – A Systematic Review on Quality of Evidence and Clinical Effects": Juel (2017) Cross-over (Individually Randomised) - Per Protocol Analysis.docx

### The RoB 2.0 tool (individually randomized, cross-over trials)

| **Assessor name/initials** | NV |
| --- | --- |
| **Study ID and/or reference(s)** | Juel (2017) |

**Study design**

| □ | Randomized parallel group trial |
| --- | --- |
| □ | Cluster-randomized trial |
| 🗹 | Randomized cross-over or other matched design |

| **Specify which outcome is being assessed for risk of bias** | ECG (HR, cardiac vagal tone - measure of efferent vagal activity), PPG finger cuff (SBP, DBP, MBP), ECG and PPG combined to calculated cardiac sensitivity to baroreflex (CSB) - measure of PNS afferent activity, pain threshold (right quadricep muscle and right tibia bone), cold pressor test (conditioned pain modulation), and drink test with ultrasound imaging (gastric emptying rate, frequency of antral contractions, amplitude of antral contractions) of active compared to sham. |
| --- | --- |

| **Specify the numerical result being assessed.** In case of multiple alternative analyses being presented, specify the numeric result (e.g. RR = 1.52 (95% CI 0.83 to 2.77) and/or a reference (e.g. to a table, figure or paragraph) that uniquely defines the result being assessed. | Within-group: Significant decrease in HR during sham (p<0.001), cardiac vagal tone in active (p<0.01), systolic blood pressure in active (p<0.01), and mean blood presure in active (p<0.05). Rest not significant. Between-group: Higher cardiac vagal tone in active, heart rate, and lower CPM. Rest not significant (including all GI motility parameters). |
| --- | --- |

**Is your aim for this study…?**

| □ | to assess the effect of *assignment to intervention* |
| --- | --- |
| 🗹 | to assess the effect of *starting and adhering to intervention* |

**Which of the following sources have you obtained to help inform your risk of bias judgements (tick as many as apply)?**

🗹	Journal article(s) with results of the trial

□	Trial protocol

□	Statistical analysis plan (SAP)

□	Non-commercial trial registry record (e.g. ClinicalTrials.gov record)

□	Company-owned trial registry record (e.g. GSK Clinical Study Register record)

□	“Grey literature” (e.g. unpublished thesis)

(other options deleted)

#### Risk of bias assessment for a cross-over trial with interest in the effect of starting and adhering to intervention

| **Domain** | **Signalling questions** | **Response options** | **Description/Support for judgement** |
| --- | --- | --- | --- |
| **Bias arising from the randomization process** | 1.1 Was the allocation sequence random? | PY | Centrally randomised using website.  Concealment inferred. |
|  | 1.2 Was the allocation sequence concealed until participants were recruited and assigned to interventions? | PY |  |
|  | 1.3 Were there baseline imbalances that suggest a problem with the randomization process? | NI |  |
|  | 1.4 Is a roughly equal proportion of participants allocated to each of the two groups? | PY | Equal split assumed. |
|  | 1.5 If N/PN to 1.4: Are period effects included in the analysis? | NA |  |
|  | **Risk of bias judgement** | Low |  |
|  | Optional: What is the predicted direction of bias arising from the randomization process? | Blank |  |
| **Bias due to deviations from intended interventions** | 2.1. Were participants aware of their assigned intervention during each period of the trial? | PN | Sham for both stimulation and breathing, but not evaluated.  Single-blinded. |
|  | 2.2. Were carers and trial personnel aware of participants' assigned intervention during each period of the trial? | Y |  |
|  | 2.3. If Y/PY/NI to 2.1 or 2.2: Were important co-interventions balanced across the two interventions? | NI | Difference in use of rescue medicine for pain flare-ups not reported. |
|  | 2.4. Was the intervention implemented successfully? | Y | Not reported otherwise. Stimulation amplitude continuously adjusted to counter habituation. |
|  | 2.5. Did study participants adhere to the assigned intervention regimen? | Y | Short term intervention |
|  | 2.6. If N/PN/NI to 2.3, 2.4 or 2.5: Was an appropriate analysis used to estimate the effect of starting and adhering to the intervention? | NI | Standard ITT analysis used. |
|  | 2.7 Was there sufficient time for any carry-over effects to have disappeared before outcome assessment in the second period? | PY | Cites acupuncture study for 1 week washout period justification. |
|  | **Risk of bias judgement** | Some concerns | Sham group has some significant differences from baseline. |
|  | Optional: What is the predicted direction of bias due to deviations from intended interventions? | Blank |  |
| **Bias due to missing outcome data** | 3.1 Were outcome data available for all, or nearly all, participants randomized? | PY | Missing for 6 of 20 participant’s ultrasound recordings. Does this mean 6 bad recordings or 12 (crossover)? CSB results not reported. |
|  | 3.2 If N/PN/NI to 3.1: Are the proportions of missing outcome data and reasons for missing outcome data similar across interventions? | NA |  |
|  | 3.3 If N/PN/NI to 3.1: Is there evidence that results were robust to the presence of missing outcome data? | NA |  |
|  | **Risk of bias judgement** | Low |  |
|  | Optional: What is the predicted direction of bias due to missing outcome data? | Blank |  |
| **Bias in measurement of the outcome** | 4.1 Were outcome assessors aware of the intervention received by study participants? | Y | Single-blinded |
|  | 4.2 If Y/PY/NI to 4.1: Was the assessment of the outcome likely to be influenced by knowledge of intervention received? | PN | Objective measures, although investigator could cue participants. |
|  | **Risk of bias judgement** | Low |  |
|  | Optional: What is the predicted direction of bias due to measurement of the outcome? | Blank |  |
| **Bias in selection of the reported result** | Are the reported outcome data likely to have been selected, on the basis of the results, from... |  | Only two Vagal tone and QST measurments made for before/after. |
|  | 5.1. ... multiple outcome measurements (e.g. scales, definitions, time points) within the outcome domain? | PN |  |
|  | 5.2 ... multiple analyses of the data? | NI | Both within group and between group reported. Several outcome measured with none defined as primary. No multiple comparisons between these different variables. Any of which would have been a positive trial outcome. CSB results not reported. |
|  | 5.3 … the outcome of a statistical test for carry-over? | PN | No carry-over analysis done. |
|  | **Risk of bias judgement** | Some concerns |  |
|  | Optional: What is the predicted direction of bias due to selection of the reported result? | Blank |  |
| **Overall bias** | **Risk of bias judgement** | Some concerns |  |
|  | Optional:  What is the overall predicted direction of bias for this outcome? | Blank |  |
