## Supplementary Material 9 - Individual RoB Rubrics for "Auricular Vagus Neuromodulation – A Systematic Review on Quality of Evidence and Clinical Effects": Kovacic (2017) Parallel (Individually Randomised) - Per Protocol Analysis.docx

### The RoB 2.0 tool (individually randomized, parallel group trials)

| **Assessor name/initials** | NV |
| --- | --- |
| **Study ID and/or reference(s)** | Kovacic (2017) |

**Study design**

| 🗹 | Randomized parallel group trial |
| --- | --- |
| □ | Cluster-randomized trial |
| □ | Randomized cross-over or other matched design |

| **Specify which outcome is being assessed for risk of bias** | Change in worst pain intensity and composite of Painr-Frequency-Severity-Duration (PFSD) scale from baseline to end of week 3. |
| --- | --- |

| **Specify the numerical result being assessed.** In case of multiple alternative analyses being presented, specify the numeric result (e.g. RR = 1.52 (95% CI 0.83 to 2.77) and/or a reference (e.g. to a table, figure or paragraph) that uniquely defines the result being assessed. | worst pain: sham has significantly higher worst pain at end of 3 weeks (median 7·0, IQR 5·0–9·0) compared to active (5·0, 4·0–7·0; p=0·003), both groups were comparable at baseline with active at (8·0, IQR 7·0–9·0) and sham at (7·5, 6·0–9·0; p=0·092). composite score: after 3 weeks, patients in the active group reported a significantly lower median PFSD composite score (8·4, IQR 3·2–16·2) than those in the sham group (15·2, 4·4–36·8) p = 0.003, scores were similar at baseline in active (24·5, IQR 16·8–33·3) and in sham (22·8, 8·4–38·2). |
| --- | --- |

**Is your aim for this study…?**

| □ | to assess the effect of *assignment to intervention* |
| --- | --- |
| 🗹 | to assess the effect of *starting and adhering to intervention* |

**Which of the following sources have you obtained to help inform your risk of bias judgements (tick as many as apply)?**

🗹	Journal article(s) with results of the trial

□	Trial protocol

□	Statistical analysis plan (SAP)

□	Non-commercial trial registry record (e.g. ClinicalTrials.gov record)

□	Company-owned trial registry record (e.g. GSK Clinical Study Register record)

□	“Grey literature” (e.g. unpublished thesis)

□	Conference abstract(s) about the trial

□	Regulatory document (e.g. Clinical Study Report, Drug Approval Package)

□	Research ethics application

□	Grant database summary (e.g. NIH RePORTER, Research Councils UK Gateway to Research)

□	Personal communication with trialist

□	Personal communication with the sponsor

#### Risk of bias assessment for a parallel group trial with interest in the effect of starting and adhering to intervention

| **Domain** | **Signalling questions** | **Response options** | **Description/Support for judgement** |
| --- | --- | --- | --- |
| **Bias arising from the randomization process** | 1.1 Was the allocation sequence random? | Y | Well reported randomization and concealment. |
|  | 1.2 Was the allocation sequence concealed until participants were recruited and assigned to interventions? | Y |  |
|  | 1.3 Were there baseline imbalances that suggest a problem with the randomization process? | PN | Baseline was generally well balanced. One medication (serotonin inhibitor) was imbalanced. |
|  | **Risk of bias judgement** | Low |  |
|  | Optional: What is the predicted direction of bias arising from the randomization process? | Blank |  |
| **Bias due to deviations from intended interventions** | 2.1. Were participants aware of their assigned intervention during the trial? | PY | Blinding evaluated by questionnaire at end of treatment, no analysis performed on results. Active group had greater fraction of subjects who believe they were in active group as compare to sham. Since questionnaire was at end of treatment, it is unclear if results are indicative of unblinding or perceived treatment effect. Questionnaire closer to start of treatment may have avoided this ambiguity. |
|  | 2.2. Were carers and trial personnel aware of participants' assigned intervention during the trial? | PN |  |
|  | 2.3. If Y/PY/NI to 2.1 or 2.2: Were important co-interventions balanced across intervention groups? | PY | Although no stimulation used as control, active was subsensory and claim was made that active and sham device had same sensation. |
|  | 2.4. Was the intervention implemented successfully? | Y |  |
|  | 2.5. Did study participants adhere to the assigned intervention regimen? | Y |  |
|  | 2.6. If N/PN/NI to 2.3, 2.4 or 2.5: Was an appropriate analysis used to estimate the effect of starting and adhering to the intervention? | NA |  |
|  | **Risk of bias judgement** | Some concerns | Due to potential unblinding indicated by questionnaire. |
|  | Optional: What is the predicted direction of bias due to deviations from intended interventions? | Favours experimental |  |
| **Bias due to missing outcome data** | 3.1 Were outcome data available for all, or nearly all, participants randomized? | Y |  |
|  | 3.2 If N/PN/NI to 3.1: Are the proportions of missing outcome data and reasons for missing outcome data similar across intervention groups? | NA |  |
|  | 3.3 If N/PN/NI to 3.1: Is there evidence that results were robust to the presence of missing outcome data? | NA |  |
|  | **Risk of bias judgement** | Low |  |
|  | Optional: What is the predicted direction of bias due to missing outcome data? | Blank |  |
| **Bias in measurement of the outcome** | 4.1 Were outcome assessors aware of the intervention received by study participants? | PN | Double blind, not evaluated. |
|  | 4.2 If Y/PY/NI to 4.1: Was the assessment of the outcome likely to be influenced by knowledge of intervention received? | NA |  |
|  | **Risk of bias judgement** | Low |  |
|  | Optional: What is the predicted direction of bias due to measurement of the outcome? | Blank |  |
| **Bias in selection of the reported result** | Are the reported outcome data likely to have been selected, on the basis of the results, from... |  |  |
|  | 5.1. ... multiple outcome measurements (e.g. scales, definitions, time points) within the outcome domain? | PN | Primary endpoint (1 of 2) pre-registered along with time point |
|  | 5.2 ... multiple analyses of the data? | PN | Several secondary endpoints pre-registered |
|  | **Risk of bias judgement** | Low |  |
|  | Optional: What is the predicted direction of bias due to selection of the reported result? | Blank |  |
| **Overall bias** | **Risk of bias judgement** | Some concerns | Mainly due to potential concerns with unblinding. Same sensation of active and sham device claimed by investigators may be true. Overall, high quality study. |
|  | Optional:  What is the overall predicted direction of bias for this outcome? | Favours experimental |  |
