## Supplementary Material 9 - Individual RoB Rubrics for "Auricular Vagus Neuromodulation – A Systematic Review on Quality of Evidence and Clinical Effects": Laqua 2014 - Parallel (Individually Randomised) - Per Protocol Analysis.docx

### The RoB 2.0 tool (individually randomized, parallel group trials)

| **Assessor name/initials** | MK |
| --- | --- |
| **Study ID and/or reference(s)** | Laqua 2014 |

**Study design**

| 🗹 | Randomized parallel group trial |
| --- | --- |
| □ | Cluster-randomized trial |
| □ | Randomized cross-over or other matched design |

| **Specify which outcome is being assessed for risk of bias** | Pain Threshold |
| --- | --- |

| **Specify the numerical result being assessed.** In case of multiple alternative analyses being presented, specify the numeric result (e.g. RR = 1.52 (95% CI 0.83 to 2.77) and/or a reference (e.g. to a table, figure or paragraph) that uniquely defines the result being assessed. | No significant difference between tVNS and placebo. 15 out of 21 volunteers responded with an increase in PT during TVNS (p < 0.01 vs baseline) but not during the placebo session.  The other 6 participants responded with a decrease in PT during and after TVNS ( p < 0.05 vs baseline), but not during the placebo session |
| --- | --- |

**Is your aim for this study…?**

| □ | to assess the effect of *assignment to intervention* |
| --- | --- |
| 🗹 | to assess the effect of *starting and adhering to intervention* |

**Which of the following sources have you obtained to help inform your risk of bias judgements (tick as many as apply)?**

□	Journal article(s) with results of the trial

□	Trial protocol

□	Statistical analysis plan (SAP)

□	Non-commercial trial registry record (e.g. ClinicalTrials.gov record)

□	Company-owned trial registry record (e.g. GSK Clinical Study Register record)

□	“Grey literature” (e.g. unpublished thesis)

□	Conference abstract(s) about the trial

□	Regulatory document (e.g. Clinical Study Report, Drug Approval Package)

□	Research ethics application

□	Grant database summary (e.g. NIH RePORTER, Research Councils UK Gateway to Research)

□	Personal communication with trialist

□	Personal communication with the sponsor

#### Risk of bias assessment for a parallel group trial with interest in the effect of starting and adhering to intervention

| **Domain** | **Signalling questions** | **Response options** | **Description/Support for judgement** |
| --- | --- | --- | --- |
| **Bias arising from the randomization process** | 1.1 Was the allocation sequence random? | PY | The study claims to be randomized |
|  | 1.2 Was the allocation sequence concealed until participants were recruited and assigned to interventions? | PY | No reason to suspect otherwise |
|  | 1.3 Were there baseline imbalances that suggest a problem with the randomization process? | NI | Healthy patients with no demographic data listed |
|  | **Risk of bias judgement** | Low |  |
|  | Optional: What is the predicted direction of bias arising from the randomization process? | NA |  |
| **Bias due to deviations from intended interventions** | 2.1. Were participants aware of their assigned intervention during the trial? | PY | The participants did not know the purpose, but knew that one stimulation was supposed to be “low” and the other one was “high” |
|  | 2.2. Were carers and trial personnel aware of participants' assigned intervention during the trial? | Y | The carers and trial personnel were not blinded |
|  | 2.3. If Y/PY/NI to 2.1 or 2.2: Were important co-interventions balanced across intervention groups? | NI | There is no information on the baseline amplitude of subject in the participants |
|  | 2.4. Was the intervention implemented successfully? | PY | The painful stimuli seemed to be a reliable intervention. |
|  | 2.5. Did study participants adhere to the assigned intervention regimen? | PY | The study took place in a controlled environment. |
|  | 2.6. If N/PN/NI to 2.3, 2.4 or 2.5: Was an appropriate analysis used to estimate the effect of starting and adhering to the intervention? | PN | Not mentionned |
|  | **Risk of bias judgement** | Some concerns |  |
|  | Optional: What is the predicted direction of bias due to deviations from intended interventions? |  |  |
| **Bias due to missing outcome data** | 3.1 Were outcome data available for all, or nearly all, participants randomized? | PY | 21/22 participants outcome data was available and analyzed |
|  | 3.2 If N/PN/NI to 3.1: Are the proportions of missing outcome data and reasons for missing outcome data similar across intervention groups? | NA |  |
|  | 3.3 If N/PN/NI to 3.1: Is there evidence that results were robust to the presence of missing outcome data? | NA |  |
|  | **Risk of bias judgement** | Low |  |
|  | Optional: What is the predicted direction of bias due to missing outcome data? | NA |  |
| **Bias in measurement of the outcome** | 4.1 Were outcome assessors aware of the intervention received by study participants? | PY | Yes, the outcomes assessors read the “pain threshold” and there is no reason to believe that they were not aware of the intervention |
|  | 4.2 If Y/PY/NI to 4.1: Was the assessment of the outcome likely to be influenced by knowledge of intervention received? | PN | No, the participants determined the pain threshold and were blinded from the purpose of the intervention |
|  | **Risk of bias judgement** | Low |  |
|  | Optional: What is the predicted direction of bias due to measurement of the outcome? | NA |  |
| **Bias in selection of the reported result** | Are the reported outcome data likely to have been selected, on the basis of the results, from... |  |  |
|  | 5.1. ... multiple outcome measurements (e.g. scales, definitions, time points) within the outcome domain? | PN |  |
|  | 5.2 ... multiple analyses of the data? | PY | The study separated the participants into those that had an anti-nociceptive and pro-nociceptive effects in order to find significance. |
|  | **Risk of bias judgement** | High | Overridden from algorithm – see above comment |
|  | Optional: What is the predicted direction of bias due to selection of the reported result? | NA |  |
| **Overall bias** | **Risk of bias judgement** | High |  |
|  | Optional:  What is the overall predicted direction of bias for this outcome? | NA |  |
