## Supplementary Material 9 - Individual RoB Rubrics for "Auricular Vagus Neuromodulation – A Systematic Review on Quality of Evidence and Clinical Effects": Maharjan (2018) Cross-over (Individually Randomised) - Per Protocol Analysis.docx

### The RoB 2.0 tool (individually randomized, cross-over trials)

| **Assessor name/initials** | RC |
| --- | --- |
| **Study ID and/or reference(s)** | Maharjan (2018) |

**Study design**

| □ | Randomized parallel group trial |
| --- | --- |
| □ | Cluster-randomized trial |
| 🗹 | Randomized cross-over or other matched design |

| **Specify which outcome is being assessed for risk of bias** | odor threshold test (OTT), supra-threshould test (STT) |
| --- | --- |

| **Specify the numerical result being assessed.** In case of multiple alternative analyses being presented, specify the numeric result (e.g. RR = 1.52 (95% CI 0.83 to 2.77) and/or a reference (e.g. to a table, figure or paragraph) that uniquely defines the result being assessed. | supra-thresold test (STT) score in figure 4. There were significant differences in the STT scores after high frequencyVNS [p=0.021 (Wilcoxon signed-rank test), correlationcoefficient (r): 0.39] but not under the low frequency VNS[ p=0.439 (Wilcoxon signed-ranked test) or placebo stimulation(p=0.083 (paired samplet-test). |
| --- | --- |

**Is your aim for this study…?**

| □ | to assess the effect of *assignment to intervention* |
| --- | --- |
| 🗹 | to assess the effect of *starting and adhering to intervention* |

**Which of the following sources have you obtained to help inform your risk of bias judgements (tick as many as apply)?**

🗹	Journal article(s) with results of the trial

🗹	Trial protocol

□	Statistical analysis plan (SAP)

□	Non-commercial trial registry record (e.g. ClinicalTrials.gov record)

□	Company-owned trial registry record (e.g. GSK Clinical Study Register record)

□	“Grey literature” (e.g. unpublished thesis)

□	Conference abstract(s) about the trial

□	Regulatory document (e.g. Clinical Study Report, Drug Approval Package)

□	Research ethics application

□	Grant database summary (e.g. NIH RePORTER, Research Councils UK Gateway to Research)

□	Personal communication with trialist

□	Personal communication with the sponsor

#### Risk of bias assessment for a cross-over trial with interest in the effect of starting and adhering to intervention

| **Domain** | **Signalling questions** | **Response options** | **Description/Support for judgement** |
| --- | --- | --- | --- |
| **Bias arising from the randomization process** | 1.1 Was the allocation sequence random? | Y | Individually randomized  No reason to suspect pre-emptive sequence revealing |
|  | 1.2 Was the allocation sequence concealed until participants were recruited and assigned to interventions? | Y |  |
|  | 1.3 Were there baseline imbalances that suggest a problem with the randomization process? | NI | N, they did not test whether there is a baseline significance with respect to the order of experiments for each participant. However, they did baseline comparison for each cross-over arm and found no significant difference. |
|  | 1.4 Is a roughly equal proportion of participants allocated to each of the two groups? | Y | Individual randomized. Simple randomisation using a randomisation table created by computer software Orders of experimental conditions were counterbalanced across the participants. |
|  | 1.5 If N/PN to 1.4: Are period effects included in the analysis? | NA |  |
|  | **Risk of bias judgement** | Low | From algorithm |
|  | Optional: What is the predicted direction of bias arising from the randomization process? | Blank |  |
| **Bias due to deviations from intended interventions** | 2.1. Were participants aware of their assigned intervention during each period of the trial? | PY | The controls were a no current placebo; For placebo difference in perception could alter results.  Experimenters were careful to maintain double-blind |
|  | 2.2. Were carers and trial personnel aware of participants' assigned intervention during each period of the trial? | PN |  |
|  | 2.3. If Y/PY/NI to 2.1 or 2.2: Were important co-interventions balanced across the two interventions? | PY | Sham replicate sensations that those in active group would receive. |
|  | 2.4. Was the intervention implemented successfully? | PY | No reported issues with intervention implementation |
|  | 2.5. Did study participants adhere to the assigned intervention regimen? | PY | No reported issues with participant adherence. |
|  | 2.6. If N/PN/NI to 2.3, 2.4 or 2.5: Was an appropriate analysis used to estimate the effect of starting and adhering to the intervention? | NA |  |
|  | 2.7 Was there sufficient time for any carry-over effects to have disappeared before outcome assessment in the second period? | PY | 24 hours is likely to be sufficient |
|  | **Risk of bias judgement** | Low | Based on the algrithm |
|  | Optional: What is the predicted direction of bias due to deviations from intended interventions? | Blank |  |
| **Bias due to missing outcome data** | 3.1 Were outcome data available for all, or nearly all, participants randomized? | Y | All outcome data were available. |
|  | 3.2 If N/PN/NI to 3.1: Are the proportions of missing outcome data and reasons for missing outcome data similar across interventions? | NA |  |
|  | 3.3 If N/PN/NI to 3.1: Is there evidence that results were robust to the presence of missing outcome data? | NA |  |
|  | **Risk of bias judgement** | Low | From algorithm |
|  | Optional: What is the predicted direction of bias due to missing outcome data? | Blank |  |
| **Bias in measurement of the outcome** | 4.1 Were outcome assessors aware of the intervention received by study participants? | PN | Investigators were careful to maintain double blind |
|  | 4.2 If Y/PY/NI to 4.1: Was the assessment of the outcome likely to be influenced by knowledge of intervention received? | NA |  |
|  | **Risk of bias judgement** | Low | From algorithm |
|  | Optional: What is the predicted direction of bias due to measurement of the outcome? | Blank |  |
| **Bias in selection of the reported result** | Are the reported outcome data likely to have been selected, on the basis of the results, from... |  |  |
|  | 5.1. ... multiple outcome measurements (e.g. scales, definitions, time points) within the outcome domain? | N | Reported results were appropriately recorded corresponding to intended outcome measurements in the registered protocol. |
|  | 5.2 ... multiple analyses of the data? | N | The analysis followed the registered protocol. |
|  | 5.3 … the outcome of a statistical test for carry-over? | PN | No reason to suspect reporting of results from only one period of the cross-over in any case |
|  | **Risk of bias judgement** | Low | From algorithm |
|  | Optional: What is the predicted direction of bias due to selection of the reported result? | Blank |  |
| **Overall bias** | **Risk of bias judgement** | Low | Well-constructed and executed study |
|  | Optional:  What is the overall predicted direction of bias for this outcome? | Favours experimental |  |
