## Supplementary Material 9 - Individual RoB Rubrics for "Auricular Vagus Neuromodulation – A Systematic Review on Quality of Evidence and Clinical Effects": Napadow (2012) Cross-over (Individually Randomised) - Per Protocol Analysis.docx

### The RoB 2.0 tool (individually randomized, cross-over trials)

| **Assessor name/initials** | JM |
| --- | --- |
| **Study ID and/or reference(s)** | Napadow (2012) |

**Study design**

| □ | Randomized parallel group trial |
| --- | --- |
| □ | Cluster-randomized trial |
| 🗹 | Randomized cross-over or other matched design |

| **Specify which outcome is being assessed for risk of bias** | pain intensity rating (deep pain), and temporal pain summation; both measured after applying pain stimuli and asking subjects to estimate the severity of their pain from 0-100, with 0 being no pain and 100 being extreme pain |
| --- | --- |

| **Specify the numerical result being assessed.** In case of multiple alternative analyses being presented, specify the numeric result (e.g. RR = 1.52 (95% CI 0.83 to 2.77) and/or a reference (e.g. to a table, figure or paragraph) that uniquely defines the result being assessed. | Reduction in deep pain intensity rating showed a significant difference in stimulation effect in repeated measures ANOVA (approximated from graph): active (17 SD 11 during stim, 19 SD 11 immediately after, 22 SD 11 15 min after), control(10 SD 11, 10 SD 11, 14 SD 11), p=0.049. Significant within-group decreases for both (p<0.05) but active tended to be larger. Reduction in temporal summation showed significant repeated measures ANOVA stim*time point interaction(approximated from graph): active (13 SD 23, 11 SD 20, 9 SD 20), control (1 SD 11.6, 8 SD 16.7, 5.5 SD 16.7), p=0.04. Significant within-group decrease only in active (p=0.05) |
| --- | --- |

**Is your aim for this study…?**

| □ | to assess the effect of *assignment to intervention* |
| --- | --- |
| 🗹 | to assess the effect of *starting and adhering to intervention* |

**Which of the following sources have you obtained to help inform your risk of bias judgements (tick as many as apply)?**

🗹	Journal article(s) with results of the trial

□	Trial protocol

□	Statistical analysis plan (SAP)

□	Non-commercial trial registry record (e.g. ClinicalTrials.gov record)

□	Company-owned trial registry record (e.g. GSK Clinical Study Register record)

□	“Grey literature” (e.g. unpublished thesis)

□	Conference abstract(s) about the trial

□	Regulatory document (e.g. Clinical Study Report, Drug Approval Package)

□	Research ethics application

□	Grant database summary (e.g. NIH RePORTER, Research Councils UK Gateway to Research)

□	Personal communication with trialist

□	Personal communication with the sponsor

#### Risk of bias assessment for a cross-over trial with interest in the effect of starting and adhering to intervention

| **Domain** | **Signalling questions** | **Response options** | **Description/Support for judgement** |
| --- | --- | --- | --- |
| **Bias arising from the randomization process** | 1.1 Was the allocation sequence random? | Y | Individually randomized.  No reason to suspect allocation sequence was pre-emptively revealed. |
|  | 1.2 Was the allocation sequence concealed until participants were recruited and assigned to interventions? | PY |  |
|  | 1.3 Were there baseline imbalances that suggest a problem with the randomization process? | PY | Baseline pain ratings differed between active and control; control was higher (p = 0.02) |
|  | 1.4 Is a roughly equal proportion of participants allocated to each of the two groups? | PY | Not specified which group numbers there were to start out, but cross-over design makes equal group size reasonable |
|  | 1.5 If N/PN to 1.4: Are period effects included in the analysis? | NA |  |
|  | **Risk of bias judgement** | Some concerns | Significant baseline difference, not really addressed by authors. Repeated measures ANOVA revealed no time or session significant effect? |
|  | Optional: What is the predicted direction of bias arising from the randomization process? | Unpredictable |  |
| **Bias due to deviations from intended interventions** | 2.1. Were participants aware of their assigned intervention during each period of the trial? | PN | Well-designed sham  Not indicated, likely that they were not blinded since they had to manage stimulation intervention, however. |
|  | 2.2. Were carers and trial personnel aware of participants' assigned intervention during each period of the trial? | NI |  |
|  | 2.3. If Y/PY/NI to 2.1 or 2.2: Were important co-interventions balanced across the two interventions? | Y | All interventions appear balanced |
|  | 2.4. Was the intervention implemented successfully? | PN | No reported issues with stimulation interventions. Some cuff algometry issues, but data was not included. |
|  | 2.5. Did study participants adhere to the assigned intervention regimen? | PN | 3 subjects dropped the trial after their first session. Data not included. |
|  | 2.6. If N/PN/NI to 2.3, 2.4 or 2.5: Was an appropriate analysis used to estimate the effect of starting and adhering to the intervention? | PN | No compensatory statistics appear to have been performed to account for removed data. |
|  | 2.7 Was there sufficient time for any carry-over effects to have disappeared before outcome assessment in the second period? | Y | 1 week between sessions should be sufficient. |
|  | **Risk of bias judgement** | High | Sham was well-executed, but too many unknowns concerning group interventions and missing data with lacking correct analysis to compensate. |
|  | Optional: What is the predicted direction of bias due to deviations from intended interventions? | Favours experimental |  |
| **Bias due to missing outcome data** | 3.1 Were outcome data available for all, or nearly all, participants randomized? | PN | Outcome data missing for the 3 that dropped out, plus a one subject whose data was dropped due to inadvertent within-session cuff pressure alterations. |
|  | 3.2 If N/PN/NI to 3.1: Are the proportions of missing outcome data and reasons for missing outcome data similar across interventions? | NI | Unclear what sessions the 3 had started in before they dropped. Individual group size not reported. |
|  | 3.3 If N/PN/NI to 3.1: Is there evidence that results were robust to the presence of missing outcome data? | NI | No evidence stating robustness of results. |
|  | **Risk of bias judgement** | Some concerns | Some missing data, proportions and robustness unclear. |
|  | Optional: What is the predicted direction of bias due to missing outcome data? | Unpredictable |  |
| **Bias in measurement of the outcome** | 4.1 Were outcome assessors aware of the intervention received by study participants? | NI | Not stated, but likely had to be in order to deliver stimulation interventions. |
|  | 4.2 If Y/PY/NI to 4.1: Was the assessment of the outcome likely to be influenced by knowledge of intervention received? | PN | Scores were acquired verbally from patients, so unlikely that assessors would affect this with bias. However, investigators could feasibly influence patient response by asking in a more/less hopeful tone of voice, priming them to expect one result or another. |
|  | **Risk of bias judgement** | Low | Outcome measurement methods seem sufficient |
|  | Optional: What is the predicted direction of bias due to measurement of the outcome? | Blank |  |
| **Bias in selection of the reported result** | Are the reported outcome data likely to have been selected, on the basis of the results, from... |  | Outcomes acquired from single 0-100 scales in the case of pain and anxiety. Multiple measurements not used. |
|  | 5.1. ... multiple outcome measurements (e.g. scales, definitions, time points) within the outcome domain? | PN |  |
|  | 5.2 ... multiple analyses of the data? | PY | Multiple statistics used; it seems like they try to claim ANOVA showing no interactions between stim and time justifies significant difference between baseline values. |
|  | 5.3 … the outcome of a statistical test for carry-over? | PN | No evidence to suggest this. |
|  | **Risk of bias judgement** | High | Serious concerns with use of different statistics to justify significant differences at baseline |
|  | Optional: What is the predicted direction of bias due to selection of the reported result? | Favours experimental |  |
| **Overall bias** | **Risk of bias judgement** | High | Initially seemed well designed, but this study’s analyses and justifications seemed inaccurate given the missing data. Significant risk of bias. |
|  | Optional:  What is the overall predicted direction of bias for this outcome? | Favours experimental |  |
