## Supplementary Material 9 - Individual RoB Rubrics for "Auricular Vagus Neuromodulation – A Systematic Review on Quality of Evidence and Clinical Effects": Stowell 2019 Cross-over (Individually Randomised) - Per Protocol Analysis.docx

### The RoB 2.0 tool (individually randomized, cross-over trials)

| **Assessor name/initials** | MK |
| --- | --- |
| **Study ID and/or reference(s)** | Stowell et al. 2019 |

**Study design**

| □ | Randomized parallel group trial |
| --- | --- |
| □ | Cluster-randomized trial |
| 🗹 | Randomized cross-over or other matched design |

| **Specify which outcome is being assessed for risk of bias** | Arterial blood pressure (Systolic blood pressure, Diastolic blood pressure, Mean arterial pressure) using an inflatable finger cuff in conjunction with a Finometer device |
| --- | --- |

| **Specify the numerical result being assessed.** In case of multiple alternative analyses being presented, specify the numeric result (e.g. RR = 1.52 (95% CI 0.83 to 2.77) and/or a reference (e.g. to a table, figure or paragraph) that uniquely defines the result being assessed. | Decrease in arterial blood pressure (systolic) with 100Hz stimulation. Subjects presented an average reduction of -5.06±10.55 mmHg during the 100 Hz session. |
| --- | --- |

**Is your aim for this study…?**

| □ | to assess the effect of *assignment to intervention* |
| --- | --- |
| 🗹 | to assess the effect of *starting and adhering to intervention* |

**Which of the following sources have you obtained to help inform your risk of bias judgements (tick as many as apply)?**

🗹	Journal article(s) with results of the trial

□	Trial protocol

□	Statistical analysis plan (SAP)

□	Non-commercial trial registry record (e.g. ClinicalTrials.gov record)

□	Company-owned trial registry record (e.g. GSK Clinical Study Register record)

□	“Grey literature” (e.g. unpublished thesis)

□	Conference abstract(s) about the trial

□	Regulatory document (e.g. Clinical Study Report, Drug Approval Package)

□	Research ethics application

□	Grant database summary (e.g. NIH RePORTER, Research Councils UK Gateway to Research)

□	Personal communication with trialist

□	Personal communication with the sponsor

#### Risk of bias assessment for a cross-over trial with interest in the effect of starting and adhering to intervention

| **Domain** | **Signalling questions** | **Response options** | **Description/Support for judgement** |
| --- | --- | --- | --- |
| **Bias arising from the randomization process** | 1.1 Was the allocation sequence random? | PY | “The order of stimulation frequencies was randomized across subjects” |
|  | 1.2 Was the allocation sequence concealed until participants were recruited and assigned to interventions? | PY | It was not mentioned specifically but it is likely. Especially because the study commented that during the placebo-stimulation. “For Sham stimulation, subjects’ sensory thresholds were first identified, then subjects were told that they may or may not feel pulsing in the ear and that we wanted to ensure that the stimulus was not painful.” |
|  | 1.3 Were there baseline imbalances that suggest a problem with the randomization process? | NI |  |
|  | 1.4 Is a roughly equal proportion of participants allocated to each of the two groups? | Y | It’s a cross over trial  “Subjects attended five stimulation sessions in which they received RAVANS at 2, 10, 25, 100Hz or Sham stimulation during a given visit.” |
|  | 1.5 If N/PN to 1.4: Are period effects included in the analysis? | NA |  |
|  | **Risk of bias judgement** | Low |  |
|  | Optional: What is the predicted direction of bias arising from the randomization process? | NA |  |
| **Bias due to deviations from intended interventions** | 2.1. Were participants aware of their assigned intervention during each period of the trial? | PN | “The intensity of stimulation was adjusted to achieve a moderate, non-painful sensation”  “Sham stimulation, subjects’ sensory thresholds were first identified, then subjects were told that they may or may not feel pulsing in the ear and that we wanted to ensure that the stimulus was not painful”  It seems like the subjects would not have known but 5 interventions is a lot so it’s not definitive |
|  | 2.2. Were carers and trial personnel aware of participants' assigned intervention during each period of the trial? | Y | Double blind is not mentioned |
|  | 2.3. If Y/PY/NI to 2.1 or 2.2: Were important co-interventions balanced across the two interventions? | NI | A potential confound is higher frequency also means higher charge delivered since they are keeping pulse width constant. They did not report mA values for different frequencies. |
|  | 2.4. Was the intervention implemented successfully? | PN | Stimulation amplitude was a poorly controlled variable and could be leading to the effects rather than stimulation frequency |
|  | 2.5. Did study participants adhere to the assigned intervention regimen? | PY | It is likely the subjects adhered to the assigned intervention. |
|  | 2.6. If N/PN/NI to 2.3, 2.4 or 2.5: Was an appropriate analysis used to estimate the effect of starting and adhering to the intervention? | N | Stimulation amplitude was not controlled for |
|  | 2.7 Was there sufficient time for any carry-over effects to have disappeared before outcome assessment in the second period? | Y | A minimum of 24-hours was set between stimulation sessions to account for potential carry over effects. |
|  | **Risk of bias judgement** | Some concerns | The placebo (no stim) increased arterial blood pressure |
|  | Optional: What is the predicted direction of bias due to deviations from intended interventions? | NA |  |
| **Bias due to missing outcome data** | 3.1 Were outcome data available for all, or nearly all, participants randomized? | PY |  |
|  | 3.2 If N/PN/NI to 3.1: Are the proportions of missing outcome data and reasons for missing outcome data similar across interventions? | NA |  |
|  | 3.3 If N/PN/NI to 3.1: Is there evidence that results were robust to the presence of missing outcome data? | NA |  |
|  | **Risk of bias judgement** | Low | There is some high standard deviations. Since there are only 12 subjects, it would have been interesting/informational to look at individualized data. |
|  | Optional: What is the predicted direction of bias due to missing outcome data? | NA |  |
| **Bias in measurement of the outcome** | 4.1 Were outcome assessors aware of the intervention received by study participants? | PY | No mention of double blind |
|  | 4.2 If Y/PY/NI to 4.1: Was the assessment of the outcome likely to be influenced by knowledge of intervention received? | PN | “Continuous blood pressure signals were semi- automatically annotated – systolic peaks were automatically detected and then manually inspected and corrected if needed- with the LabChart Pro Blood Pressure module (ADInstruments, CO, USA). Systolic (SBP), diastolic (DBP), and mean arterial blood pressure (MAP) values were extracted from the annotated data”  The blood pressure was semi automatically measured. The frequency of “correction” was not mentioned. |
|  | **Risk of bias judgement** | Low | It is unlikely that the instruments’ measurements were greatly affected by the outcome assessors |
|  | Optional: What is the predicted direction of bias due to measurement of the outcome? | NA |  |
| **Bias in selection of the reported result** | Are the reported outcome data likely to have been selected, on the basis of the results, from... |  |  |
|  | 5.1. ... multiple outcome measurements (e.g. scales, definitions, time points) within the outcome domain? | PN | Since no significance was found for stimulations at 2Hz, 10 Hz, and 25Hz, the paper chose to omit any numerical data relating to this experiment. The data is only visible in figures. |
|  | 5.2 ... multiple analyses of the data? | NI | Baseline measurements were compared to measurements during the stimulation and in “recovery”. Data could also have been compared recovery to stimulation rather than baseline to stimulation. |
|  | 5.3 … the outcome of a statistical test for carry-over? | NI |  |
|  | **Risk of bias judgement** | Some concerns | Some concerns because numerical data was not report for non-significant results |
|  | Optional: What is the predicted direction of bias due to selection of the reported result? | NA |  |
| **Overall bias** | **Risk of bias judgement** | High |  |
|  | Optional:  What is the overall predicted direction of bias for this outcome? | NA |  |
