## Supplementary Material 9 - Individual RoB Rubrics for "Auricular Vagus Neuromodulation – A Systematic Review on Quality of Evidence and Clinical Effects": Tobaldini (2019) Cross-over (Individually Randomised) - Per Protocol Analysis.docx

### The RoB 2.0 tool (individually randomized, cross-over trials)

| **Assessor name/initials** | JM |
| --- | --- |
| **Study ID and/or reference(s)** | Tobaldini (2019) |

**Study design**

| □ | Randomized parallel group trial |
| --- | --- |
| □ | Cluster-randomized trial |
| 🗹 | Randomized cross-over or other matched design |

| **Specify which outcome is being assessed for risk of bias** | HR, LF/HF, systolic arterial BP variance, R-R interval pattern, and respiratory rate at rest and under orthostatic stress |
| --- | --- |

| **Specify the numerical result being assessed.** In case of multiple alternative analyses being presented, specify the numeric result (e.g. RR = 1.52 (95% CI 0.83 to 2.77) and/or a reference (e.g. to a table, figure or paragraph) that uniquely defines the result being assessed. | At rest: median (25th-75th) significant HR decrease [63(60-66) active vs 66(61-68) control, p<0.01], SAP variance decrease [13(6-19) active vs 19(8-27) control, p<0.01], 0 variability interval pattern % [17(5-20) active vs 18(8-27) control, p<0.01]. During 75 degree tilt table procedure: some R-R interval percentage decrease compared to at rest condition [-23(-29--17) active vs -21(-29--13) control, p=0.07] and respiratory rate increase [6(1-12) active vs -5(-14-9) control, p=0.08]. |
| --- | --- |

**Is your aim for this study…?**

| □ | to assess the effect of *assignment to intervention* |
| --- | --- |
| 🗹 | to assess the effect of *starting and adhering to intervention* |

**Which of the following sources have you obtained to help inform your risk of bias judgements (tick as many as apply)?**

🗹	Journal article(s) with results of the trial

□	Trial protocol

□	Statistical analysis plan (SAP)

□	Non-commercial trial registry record (e.g. ClinicalTrials.gov record)

□	Company-owned trial registry record (e.g. GSK Clinical Study Register record)

□	“Grey literature” (e.g. unpublished thesis)

□	Conference abstract(s) about the trial

□	Regulatory document (e.g. Clinical Study Report, Drug Approval Package)

□	Research ethics application

□	Grant database summary (e.g. NIH RePORTER, Research Councils UK Gateway to Research)

□	Personal communication with trialist

□	Personal communication with the sponsor

#### Risk of bias assessment for a cross-over trial with interest in the effect of starting and adhering to intervention

| **Domain** | **Signalling questions** | **Response options** | **Description/Support for judgement** |
| --- | --- | --- | --- |
| **Bias arising from the randomization process** | 1.1 Was the allocation sequence random? | Y | Specified as random  No reason to suspect pre-emptive revealing of the allocation sequence |
|  | 1.2 Was the allocation sequence concealed until participants were recruited and assigned to interventions? | PY |  |
|  | 1.3 Were there baseline imbalances that suggest a problem with the randomization process? | PN | Reported baseline values fall within IQR |
|  | 1.4 Is a roughly equal proportion of participants allocated to each of the two groups? | NI | Results of group sizes after randomization not reported |
|  | 1.5 If N/PN to 1.4: Are period effects included in the analysis? | NA |  |
|  | **Risk of bias judgement** | Some concerns | From algorithm |
|  | Optional: What is the predicted direction of bias arising from the randomization process? | Unpredictable |  |
| **Bias due to deviations from intended interventions** | 2.1. Were participants aware of their assigned intervention during each period of the trial? | Y | No blind |
|  | 2.2. Were carers and trial personnel aware of participants' assigned intervention during each period of the trial? | Y |  |
|  | 2.3. If Y/PY/NI to 2.1 or 2.2: Were important co-interventions balanced across the two interventions? | N | Active with stim vs control with no stimulator present at all makes for pretty serious imbalance. Additional issues with time difference between active and control “at rest” condition (20 min active vs 10 min control) which could affect recorded resting HR due to time spent relaxed in supine position |
|  | 2.4. Was the intervention implemented successfully? | PY | No reported instances of failed interventions |
|  | 2.5. Did study participants adhere to the assigned intervention regimen? | PY | No reported noncompliance |
|  | 2.6. If N/PN/NI to 2.3, 2.4 or 2.5: Was an appropriate analysis used to estimate the effect of starting and adhering to the intervention? | PN | No corrective statistics performed |
|  | 2.7 Was there sufficient time for any carry-over effects to have disappeared before outcome assessment in the second period? | PY | 24 h seems like enough compared to most tVNS studies |
|  | **Risk of bias judgement** | High | From algorithm |
|  | Optional: What is the predicted direction of bias due to deviations from intended interventions? | Favours experimental |  |
| **Bias due to missing outcome data** | 3.1 Were outcome data available for all, or nearly all, participants randomized? | Y | No subjects reported missing |
|  | 3.2 If N/PN/NI to 3.1: Are the proportions of missing outcome data and reasons for missing outcome data similar across interventions? | NA |  |
|  | 3.3 If N/PN/NI to 3.1: Is there evidence that results were robust to the presence of missing outcome data? | NA |  |
|  | **Risk of bias judgement** | Low | From algorithm |
|  | Optional: What is the predicted direction of bias due to missing outcome data? | Blank |  |
| **Bias in measurement of the outcome** | 4.1 Were outcome assessors aware of the intervention received by study participants? | Y | No blind |
|  | 4.2 If Y/PY/NI to 4.1: Was the assessment of the outcome likely to be influenced by knowledge of intervention received? | PN | Outcomes were physiological measurements |
|  | **Risk of bias judgement** | Low | From algorithm |
|  | Optional: What is the predicted direction of bias due to measurement of the outcome? | Blank |  |
| **Bias in selection of the reported result** | Are the reported outcome data likely to have been selected, on the basis of the results, from... |  | Many different subsets of measurements taken (e.g. LF and HF subsets of LF/HF) with no intentions specified |
|  | 5.1. ... multiple outcome measurements (e.g. scales, definitions, time points) within the outcome domain? | NI |  |
|  | 5.2 ... multiple analyses of the data? | PN | Between-group comparisons, with t-test or Wilcoxon signed rank as necessary |
|  | 5.3 … the outcome of a statistical test for carry-over? | PN | No carry-over tests performed |
|  | **Risk of bias judgement** | Some concerns | From algorithm |
|  | Optional: What is the predicted direction of bias due to selection of the reported result? | Unpredictable |  |
| **Overall bias** | **Risk of bias judgement** | High |  |
|  | Optional:  What is the overall predicted direction of bias for this outcome? | Unpredictable |  |
