## Supplementary Material 9 - Individual RoB Rubrics for "Auricular Vagus Neuromodulation – A Systematic Review on Quality of Evidence and Clinical Effects": Tutar (2020) Parallel (Individually Randomised) - Per Protocol Analysis.docx

### The RoB 2.0 tool (individually randomized, parallel group trials)

| **Assessor name/initials** | JM |
| --- | --- |
| **Study ID and/or reference(s)** | Tutar (2020) |

**Study design**

| 🗹 | Randomized parallel group trial |
| --- | --- |
| □ | Cluster-randomized trial |
| □ | Randomized cross-over or other matched design |

| **Specify which outcome is being assessed for risk of bias** | Tinnitus Handicap Inventory (THI) and Depression Anxiety Stress Scales (DASS) |
| --- | --- |

| **Specify the numerical result being assessed.** In case of multiple alternative analyses being presented, specify the numeric result (e.g. RR = 1.52 (95% CI 0.83 to 2.77) and/or a reference (e.g. to a table, figure or paragraph) that uniquely defines the result being assessed. | ITT between: THI difference in active (unilateral -27.6 ± 18.8, bilateral -26.45 ± 19.36) vs control (-9.30 ± 8.67), p=0.001. DASS depression active (unilateral -7.95 ± 5.77, bilateral -8.75 ± 7.11) vs control (-3.65 ± 3.44), p=0.013. DASS anxiety active (unilateral -7.8 ± 5.95, bilateral -9.10 ± 6.11) vs control (-3.20 ± 2.98), p=0.002. DASS stress active (unilateral -7.05 ± 5.85, bilateral -9.50 ± 5.48) vs control (-3.40 ± 3.58), p=0.001. ITT within: All THI and DASS decrease p=0.001. |
| --- | --- |
