## Supplementary Material 9 - Individual RoB Rubrics for "Auricular Vagus Neuromodulation – A Systematic Review on Quality of Evidence and Clinical Effects": Yu (2017) Parallel (Individually Randomised) Per Protocol Analysis.docx

### The RoB 2.0 tool (individually randomized, parallel group trials)

| **Assessor name/initials** | JM |
| --- | --- |
| **Study ID and/or reference(s)** | Yu (2017) |

**Study design**

| 🗹 | Randomized parallel group trial |
| --- | --- |
| □ | Cluster-randomized trial |
| □ | Randomized cross-over or other matched design |

| **Specify which outcome is being assessed for risk of bias** | Effect of LLTS on incidence of VPBs |
| --- | --- |

| **Specify the numerical result being assessed.** In case of multiple alternative analyses being presented, specify the numeric result (e.g. RR = 1.52 (95% CI 0.83 to 2.77) and/or a reference (e.g. to a table, figure or paragraph) that uniquely defines the result being assessed. | VPB incidence over 7 days was 154+-115 vs 551+-214 control |
| --- | --- |

**Is your aim for this study…?**

| □ | to assess the effect of *assignment to intervention* |
| --- | --- |
| 🗹 | to assess the effect of *starting and adhering to intervention* |

**Which of the following sources have you obtained to help inform your risk of bias judgements (tick as many as apply)?**

🗹	Journal article(s) with results of the trial

□	Trial protocol

□	Statistical analysis plan (SAP)

□	Non-commercial trial registry record (e.g. ClinicalTrials.gov record)

□	Company-owned trial registry record (e.g. GSK Clinical Study Register record)

□	“Grey literature” (e.g. unpublished thesis)

□	Conference abstract(s) about the trial

□	Regulatory document (e.g. Clinical Study Report, Drug Approval Package)

□	Research ethics application

□	Grant database summary (e.g. NIH RePORTER, Research Councils UK Gateway to Research)

□	Personal communication with trialist

□	Personal communication with the sponsor

#### Risk of bias assessment for a parallel group trial with interest in the effect of starting and adhering to intervention

| **Domain** | **Signalling questions** | **Response options** | **Description/Support for judgement** |
| --- | --- | --- | --- |
| **Bias arising from the randomization process** | 1.1 Was the allocation sequence random? | PY | Individuals randomised to experiment or control  No reason to suspect that investigator or participant knew of allocation. |
|  | 1.2 Was the allocation sequence concealed until participants were recruited and assigned to interventions? | PY |  |
|  | 1.3 Were there baseline imbalances that suggest a problem with the randomization process? | N | Baseline values have no significant differences |
|  | **Risk of bias judgement** | Low | Randomization procedure was adequate for this study |
|  | Optional: What is the predicted direction of bias arising from the randomization process? | Blank |  |
| **Bias due to deviations from intended interventions** | 2.1. Were participants aware of their assigned intervention during the trial? | PY | Control was placebo (no stim) so participants could figure out if they were in experimental or control  Only echocardiographers stated as blinded. |
|  | 2.2. Were carers and trial personnel aware of participants' assigned intervention during the trial? | PY |  |
|  | 2.3. If Y/PY/NI to 2.1 or 2.2: Were important co-interventions balanced across intervention groups? | PN | Possible differences in drugs administered by physicians in charge of each subject |
|  | 2.4. Was the intervention implemented successfully? | Y | Y. No confounds reported |
|  | 2.5. Did study participants adhere to the assigned intervention regimen? | Y | Y. No divergences from protocol reported |
|  | 2.6. If N/PN/NI to 2.3, 2.4 or 2.5: Was an appropriate analysis used to estimate the effect of starting and adhering to the intervention? | NI | No compensatory statistics appear to have been used |
|  | **Risk of bias judgement** | High | From algorithm |
|  | Optional: What is the predicted direction of bias due to deviations from intended interventions? | Favours experimental |  |
| **Bias due to missing outcome data** | 3.1 Were outcome data available for all, or nearly all, participants randomized? | PY | Most data available, a few excluded due to developing left main or multiple coronary artery disease |
|  | 3.2 If N/PN/NI to 3.1: Are the proportions of missing outcome data and reasons for missing outcome data similar across intervention groups? | NA |  |
|  | 3.3 If N/PN/NI to 3.1: Is there evidence that results were robust to the presence of missing outcome data? | NA |  |
|  | **Risk of bias judgement** | Low | Relevant data included |
|  | Optional: What is the predicted direction of bias due to missing outcome data? | Blank |  |
| **Bias in measurement of the outcome** | 4.1 Were outcome assessors aware of the intervention received by study participants? | PY | Only echocardiographers were specified to have been blinded. |
|  | 4.2 If Y/PY/NI to 4.1: Was the assessment of the outcome likely to be influenced by knowledge of intervention received? | N | VBP incidence and biomarker levels are unlikely to change because of assessor bias. |
|  | **Risk of bias judgement** | Low | Difficult for even biased investigator change measurement outcome |
|  | Optional: What is the predicted direction of bias due to measurement of the outcome? | Blank |  |
| **Bias in selection of the reported result** | Are the reported outcome data likely to have been selected, on the basis of the results, from... |  | Multiple time points in some outcome measurements |
|  | 5.1. ... multiple outcome measurements (e.g. scales, definitions, time points) within the outcome domain? | NI |  |
|  | 5.2 ... multiple analyses of the data? | N | Only between-group comparisons |
|  | **Risk of bias judgement** | Some concerns | From algorithm |
|  | Optional: What is the predicted direction of bias due to selection of the reported result? | Unpredictable |  |
| **Overall bias** | **Risk of bias judgement** | High | Placebo brings up issues with accidental participant unblinding, in addition to differences between drug treatments at physician discretion |
|  | Optional:  What is the overall predicted direction of bias for this outcome? | Unpredictable |  |
