## Supplementary Material 9 - Individual RoB Rubrics for "Auricular Vagus Neuromodulation – A Systematic Review on Quality of Evidence and Clinical Effects": Zamotrinsky (2001) Parallel (Individually Randomised) - Per Protocol Analysis.docx

### The RoB 2.0 tool (individually randomized, parallel group trials)

| **Assessor name/initials** | NV |
| --- | --- |
| **Study ID and/or reference(s)** | Zamotrinsky (2001) |

**Study design**

| 🗹 | Randomized parallel group trial |
| --- | --- |
| □ | Cluster-randomized trial |
| □ | Randomized cross-over or other matched design |

| **Specify which outcome is being assessed for risk of bias** | HR, BP, LV diastolic function, LV filling (within group comparison in VNS group) and transient HF in 3-4 coronoary artery bypass post-op period, histology of right heart auricle (above atrium) resected during surgery for sympathetic nerve and vascular density (between group comparison VNS and untreated) |
| --- | --- |

| **Specify the numerical result being assessed.** In case of multiple alternative analyses being presented, specify the numeric result (e.g. RR = 1.52 (95% CI 0.83 to 2.77) and/or a reference (e.g. to a table, figure or paragraph) that uniquely defines the result being assessed. | VNS increased the density of noradrenergic plexuses (method measures how much NE is stored in nerves (i.e. not released meaning less sympathetic activity)): 1.3 SD 0.6% (n=8) VNS treated vs. 0.5 SD 0.3% (n=10) non-treated patients (p < 0.05) and vascular density 3.3 SD 0.8% VNS and 2.0 SD 1.3% untreated (p < 0.05). Early in the postoperative period 1-7 days after surgery, heart failure developed in 1/8 VNS-treated patients and in 9/10 of non-treated patients (p<0.05). For first few session, HR dropped during VNS and SBP increased when VNS stopped, for the later sessions this HR and BP effect didn't occur and anti-angina effects took its place. Improvement in ECG waveforms. Echostudies revealed an average increase of left ventricular contractility of 16% (p < 0.05) within group comparison. |
| --- | --- |

**Is your aim for this study…?**

| □ | to assess the effect of *assignment to intervention* |
| --- | --- |
| 🗹 | to assess the effect of *starting and adhering to intervention* |

**Which of the following sources have you obtained to help inform your risk of bias judgements (tick as many as apply)?**

🗹	Journal article(s) with results of the trial

□	Trial protocol

(deleted rest of list – no selection)

#### Risk of bias assessment for a parallel group trial with interest in the effect of starting and adhering to intervention

| **Domain** | **Signalling questions** | **Response options** | **Description/Support for judgement** |
| --- | --- | --- | --- |
| **Bias arising from the randomization process** | 1.1 Was the allocation sequence random? | Y | Specified 18 subjects randomly assigned to treatment or control group. No details. |
|  | 1.2 Was the allocation sequence concealed until participants were recruited and assigned to interventions? | NI |  |
|  | 1.3 Were there baseline imbalances that suggest a problem with the randomization process? | PN | N=8 treatment, n=10 control |
|  | **Risk of bias judgement** | Low |  |
|  | Optional: What is the predicted direction of bias arising from the randomization process? | Blank |  |
| **Bias due to deviations from intended interventions** | 2.1. Were participants aware of their assigned intervention during the trial? | Y | No intervention as control  Investigator blind for echocardiogram tech and postop care team. |
|  | 2.2. Were carers and trial personnel aware of participants' assigned intervention during the trial? | PN |  |
|  | 2.3. If Y/PY/NI to 2.1 or 2.2: Were important co-interventions balanced across intervention groups? | PN | Large placebo effect from novel treatment and increased attention during pre-op care. |
|  | 2.4. Was the intervention implemented successfully? | PY |  |
|  | 2.5. Did study participants adhere to the assigned intervention regimen? | PY |  |
|  | 2.6. If N/PN/NI to 2.3, 2.4 or 2.5: Was an appropriate analysis used to estimate the effect of starting and adhering to the intervention? | N | Co-intervention applies to entire group. |
|  | **Risk of bias judgement** | Some concerns | Investigator blind maintained but no patient blind. Large HR drop during 15 mins of treatment may be due to relaxation in the supine position. Control group did not have these measurements. |
|  | Optional: What is the predicted direction of bias due to deviations from intended interventions? | Favours experimental | Placebo effect from novel medical device. |
| **Bias due to missing outcome data** | 3.1 Were outcome data available for all, or nearly all, participants randomized? | Y |  |
|  | 3.2 If N/PN/NI to 3.1: Are the proportions of missing outcome data and reasons for missing outcome data similar across intervention groups? | NA |  |
|  | 3.3 If N/PN/NI to 3.1: Is there evidence that results were robust to the presence of missing outcome data? | NA |  |
|  | **Risk of bias judgement** | Low |  |
|  | Optional: What is the predicted direction of bias due to missing outcome data? | Blank |  |
| **Bias in measurement of the outcome** | 4.1 Were outcome assessors aware of the intervention received by study participants? | PN | Investigator blind for echocardiogram tech and postop care team. |
|  | 4.2 If Y/PY/NI to 4.1: Was the assessment of the outcome likely to be influenced by knowledge of intervention received? | NA |  |
|  | **Risk of bias judgement** | Some concerns | However, team delivering aVNS and surgery team may not be blinded. No intervention control. |
|  | Optional: What is the predicted direction of bias due to measurement of the outcome? | Favours experimental |  |
| **Bias in selection of the reported result** | Are the reported outcome data likely to have been selected, on the basis of the results, from... |  |  |
|  | 5.1. ... multiple outcome measurements (e.g. scales, definitions, time points) within the outcome domain? | NI | No pre-registration. Many outcome measures. Relevance of outcome measures well justified. |
|  | 5.2 ... multiple analyses of the data? | NI | Between and within group comparison used. |
|  | **Risk of bias judgement** | Some concerns |  |
|  | Optional: What is the predicted direction of bias due to selection of the reported result? | Blank |  |
| **Overall bias** | **Risk of bias judgement** | High | Largely due to poor control (no intervention) and confounds such as additional care for treatment group and HR/blood pressure effects which may be due to relaxation in supine position during aVNS delivery. |
|  | Optional:  What is the overall predicted direction of bias for this outcome? | Favours experimental |  |
